# Development and content validity of the Patient Safety in Radiation Oncology questionnaire (PaSaRO): A multi-method study

**DOI:** 10.64898/2026.01.09.26343762

**Authors:** Maximilian Grohmann, Eva Christalle, Felicitas Schwenzer, Maria Jäckel, Nina Michalowski, Isabelle Scholl, Andrea Baehr

## Abstract

**Background:** Currently, no comprehensive tool exists to systematically assess patient safety in radiation oncology (RO). To address this gap, we developed the Patient Safety in Radiation Oncology questionnaire (PaSaRO), a German instrument enabling RO professionals to evaluate patient safety within their departments.

**Methods:** Building on a literature review identifying 145 patient safety indicators (PSIs), we determined further PSIs via two focus groups with RO professionals, patient interviews, and expert consultations. RO professionals were recruited through professional networks and societies, while patients were recruited as a convenience sample at a university hospital centre. Content validity was ensured by assessing relevance and comprehensiveness through a Delphi study and comprehensibility through cognitive interviews with RO professionals.

**Findings:** Two focus groups generated 48 new PSIs, while nine patient interviews contributed 15 PSIs, and three experts suggested 12 more. In combination with the PSI from the review, a total of 213 PSIs were compiled and subsequently rated in the Delphi study by 84 professionals in the first round and 72 in the second. During this process, seven additional PSIs were suggested, and 158 were ultimately deemed relevant. In cognitive interviews, 43 PSIs were linguistically refined to improve clarity.

**Interpretation:** This study produced a pilot version of the PaSaRO, comprising 158 consensus-based PSIs—the first comprehensive questionnaire specifically developed for RO. The rigorous multi-method development process ensures strong content validity. Future research will conduct psychometric evaluation, after which PaSaRO may serve as a standardized tool for assessing, monitoring, and improving patient safety in RO.

## INTRODUCTION

Patient safety is a critical aspect of healthcare delivery, particularly in complex and high-risk disciplines such as radiation oncology (RO). RO requires precise coordination among radiation oncologists, medical physicists, radiation therapists, oncology nurses, medical receptionists and other healthcare professionals.^1^ Given the intricacies of treatment planning, dose calculation, and equipment operation, RO is inherently prone to errors arising from human factors, system-level deficiencies, or a combination of both. Although novel treatment protocols and technological advancements – such as ultra-hypofractionated treatments and online adaptive therapy – have expanded therapeutic possibilities, they have also introduced additional layers of complexity. This increases the need for robust quality assurance mechanisms and continuous safety monitoring to mitigate potential risks.^2^

Patient Safety Indicators (PSIs) have been proposed as key metrics to evaluate and enhance the safety of healthcare delivery.^3,4^ Distinct from quality indicators, which evaluate the efficiency and effectiveness of healthcare processes, PSIs specifically aim to reduce preventable harm. They do this by examining specific healthcare processes, structures, or outcomes, serving as tools to monitor, assess, and enhance the safety of care.^3^ The challenge, however, lies in tailoring existing PSI sets for application within RO. Most existing PSIs were developed with a broad healthcare context in mind – often emphasizing inpatient care scenarios such as fall prevention or pressure ulcer management^4^ – and may not sufficiently address the predominantly outpatient setting of RO practice. Moreover, the unique attributes of RO – its technical complexity, the precision required in treatment delivery, and the distinct spectrum of potential adverse effects – call for a specialized set of PSIs designed to capture and mitigate these specific risks. While internationally a few PSI sets have been proposed specifically for RO,^5–8^ none of them comprehensively covers all relevant aspects including supportive care, risk management and safety culture.^9^ In a preliminary work, we generated an initial set of RO-specific PSIs through a systematic literature review, resulting in 145 PSIs organized in 4 categories and 27 subcategories.^10^ These PSIs aim to be used by healthcare professionals within RO to rate the level of patient safety in their own department. Based on this, the departments can derive specific measures to improve patient safety and monitor it over time.^11^

The aim of this study was to expand this initial set of PSIs and to develop a pilot version of the Patient Safety in Radiation Oncology questionnaire (PaSaRO) - a German-language instrument designed to assess patient safety specifically within the context of RO from the perspective of healthcare professionals. As a second aim, we evaluated the content validity of the PaSaRO, examining whether its PSIs are relevant, comprehensive and comprehensible to RO professionals for the purpose of measuring patient safety in this field.^12^

## METHODS

### Study Design

The development of the PaSaRO was part of a 3-year, mixed methods study called Patient Safety in German Radiation Oncology (PaSaGeRO).^9^ As described in the study protocol^9^, it comprises three phases: (1) development, (2) psychometric testing and (3) routine implementation of the PaSaRO. As a first step of the development process, we previously conducted a systematic literature review to identify RO-specific PSIs, which were divided into four main and subcategories.^10^ Here we describe how these PSIs were expanded and subsequently tested. To do so, we structured the study in three steps, following a best-practice example for questionnaire development:^13^ (1) preparation, (2) PSI generation and (3) PSI selection and testing of content validity. An overview is given in Figure 1. The study was conducted by an interdisciplinary team including experts in RO, medical physics, psychology and statistics. No additional methodological consultation took place. We followed the DELPHISTAR reporting guideline^14^ to describe the Delphi study (Appendix 1), and the COSMIN reporting guideline^15^ (Appendix 2) to report all aspects related to the measurement properties of the PaSaRO.

**Figure 1:**
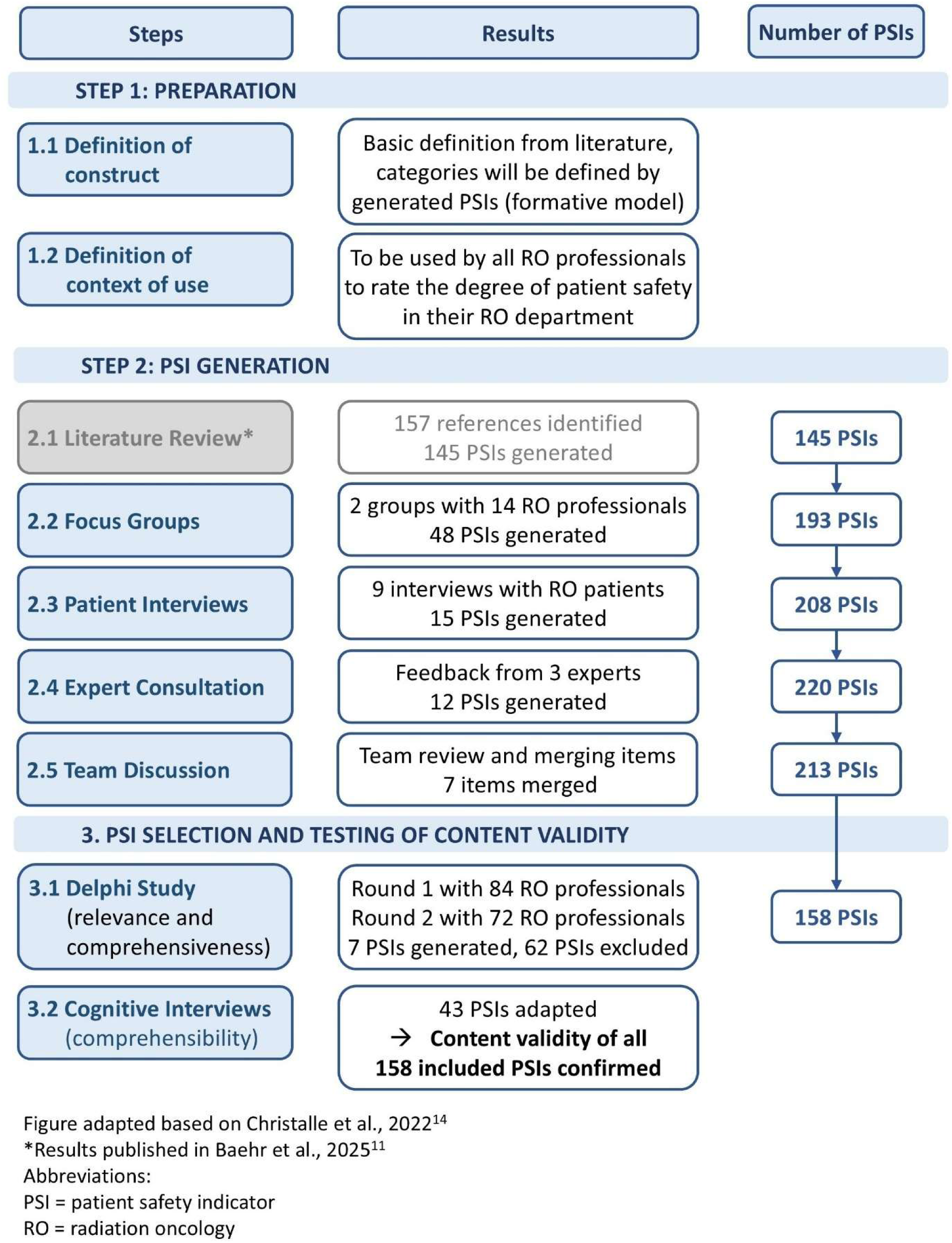
Study overview.

### Sample

For focus groups, the Delphi study and cognitive interviews, RO professionals from five professional groups (physicians, medical physicists, nurses, administrative staff and radiation therapists) were recruited between December 2023 and November 2024. Recruitment was conducted via invitation emails sent through the member mails lists of the German Society for Radiation Oncology (DEGRO), its working group on patient safety, and the risk management working group of the German Society for Medical Physics (DGMP). Furthermore, recruitment emails were sent via personal contacts of the study team to clinics and practices in Germany. In addition, a call for recruitment was posted on social media. All clinical staff members who either currently or previously worked in a German RO department were eligible for participation. During recruitment, we made a particular effort to ensure balanced representation across all professional groups by specifically reaching out to those that were underrepresented. Patient recruitment for interviews was conducted at the RO department of the University Medical Center Hamburg-Eppendorf (UKE, Hamburg, Germany). Patients were asked to participate during their discharge medical consultation on their last day of radiation therapy. All patients with tumour disease were eligible for participation if they were German-speaking, older than 18 years and capable of speaking during a phone-based interview. Patients were selected to ensure representation of different age, sex, types of tumours and treatment aim (curative vs. palliative). Participants for expert consultations were selected based on their professional background and recruited by personal contact, including one member of the study team (MG). All participants gave informed consent. Interview participants received 25€ monetary payment as an expense allowance. Participants in focus groups received 50€.

### Step 1: Preparation

#### Step 1.1: Definition of construct

During the PSI generation process, we followed the definition of patient safety provided by the Aktionsbündnis Patientensicherheit White Paper on Patient Safety,^11^ a foundational report by the German Coalition for Patient Safety highlighting key strategies and policy recommendations to improve patient safety in healthcare. In summary, patient safety is the extent to which healthcare professionals create conditions where adverse events are rare, risks are well-managed, and safety behaviours are promoted. It also reflects their commitment to continuous improvement and innovation to ensure safe care. Hence, all PSIs generated were related to aspects of this definition. The PaSaRO used a formative measurement model, meaning that we assumed the PSIs generated in the different steps collectively define patient safety. Therefore, we did not predefine different dimensions for patient safety, but instead created categories and subcategories based on the PSIs generated in the previous review and in this study.

#### Step 1.2: Definition of context of use

The PaSaRO can be used by RO professionals to systematically assess the level of patient safety in their RO departments. Moreover, it can be applied to evaluate different aspects of a department’s workflow, develop targeted interventions to improve safety, and monitor changes in patient safety over time^9^.

### Step 2: PSI generation

#### Step 2.1: Literature review

In a previous systematic literature review,^10^ we identified 157 publications on patient safety and quality indicators in RO. Two study members (AB and MG) analysed those publications and generated 145 PSIs. The wording of these indicators was refined during team discussions and subsequently grouped into four categories (Human Resources, Institutional Culture, Quality and Risk Management, and Patient-specific Processes) and 27 subcategories. The result formed the basis that was complemented in the following PSI generation steps.

#### Step 2.2: Focus groups

In February 2024, we conducted two focus groups with RO professionals from various German RO departments. Focus groups lasted for five hours and were moderated by team members (AB, EC). Using a semi-structured interview guide developed for this study (see Appendix 3), the moderators steered the conversation with key questions. At first, the study, its aims and the four categories of identified PSIs were introduced to the participants. Then the group discussion was structured using the four categories. Here, the moderator presented example PSIs from the literature review and then invited participants to share their experiences with this topic and what actions or processes either promote or hinder patient safety.

Focus groups were audio-recorded and then transcribed using the self-developed Python (v.3.11.7) program WhisperSpeaker (v.1.1) - a Python script combining open-source libraries, including OpenAI’s Whisper (using model large-v1) for speech recognition and pyannote.audio for speaker diarisation. A team member (MG, AB or FS) subsequently verified the accuracy of the transcriptions. Personal data was anonymized during transcription. We conducted qualitative data analysis similar to that used in a previous questionnaire development study.^13^ First, a predefined coding scheme was developed based on the categories and subcategories identified in the previous literature review^10^ (deductive approach). All relevant text segments were then assigned to an existing subcategory where applicable (deductive approach) using QDA Miner lite (Version 6.0.11, Provalis Research, Montreal, Canada). If no suitable subcategory was available, a new subcategory was created (inductive approach). While doing this, all coded text segments were either assigned to pre-existing PSIs or used to create new PSIs. AB coded all focus groups and interviews, while EC reviewed the coding and suggested refinements. They then met to establish a consensus version. Finally, the research team (AB, MG, FS, and EC) convened to review and discuss all newly created PSIs along with the corresponding citations.

#### Step 2.3: Patient interviews

Semi-structured patient interviews were conducted between April and May 2024. All interviews were conducted remotely via telephone and were led by a team member with a background of educational science with several years of experience in healthcare research (FS). She used an interview guide co-developed with the study’s patient advisory board (see Appendix 4) to ensure relevance and understandable wording. The guide focused on exploring participants’ perceptions of safety during radiation therapy, their conceptual understanding of patient safety, and experiences with safety-related communication in clinical settings. All interviews were audio recorded, transcribed and analysed in the same way as the focus groups.

#### Step 2.4: Expert consultation

As many aspects of radiation treatment planning were not discussed in each of the focus groups and literature search did not reveal sufficient recommendations which extend national law specifications, we asked two medical physic experts (MG, MJ) for important recommendations concerning that specific topic. Both experts developed additional PSIs during a face-to-face meeting. Furthermore, we asked a hospital pharmacist (NM) to review the PSIs and corresponding literature citations, assessing their applicability and clarity. She added further relevant recommendations to complement the PSIs, regarding issues with antitumoral medication and other medication which are especially relevant in the RO routine. Based on her recommendations, AB and NM worked together to create PSIs in a process of written feedback and adaptation until they reached consensus. All PSIs created by experts were then reviewed by all study team members.

#### Step 2.5: Team discussion

Finally, all items were reviewed in a team discussion to ensure that items are consistently worded and to merge items with similar content.

### Step 3: PSI selection and testing of content validity

Content validity is defined as the degree to which the items (here PSIs) of a questionnaire are relevant, comprehensive and comprehensible regarding the construct, target population and context of use.^12^ Below, we outline the two steps undertaken to ensure the content validity of the PaSaRO.

#### Step 3.1: Checking relevance and comprehensiveness in a Delphi study

To test relevance and comprehensiveness, we conducted a Delphi study between July and September 2024. This method is appropriate for indicator selection, as it integrates the opinions of experts with diverse backgrounds in a consensus process, while its anonymity prevents dominance by individuals due to hierarchy or perceived expertise.^16^ The aim of the Delphi study was to reach consensus on which PSIs are relevant. Experts were defined as all individuals currently or previously working in one of the RO professional groups, regardless of years of experience. Decisions on design and cut-off values were based on literature on Delphi studies with the aim to select healthcare quality indicators.^16,17^ We used a modified online Delphi study with a predefined length of two rounds. Participants received personalized survey links via LimeSurvey (LimeSurvey GmbH, Hamburg, Germany), with each round remaining open for four weeks. To maximize response rates, automated reminders were sent once per week.

In round 1, participants rated the relevance of each PSI for patient safety in RO using a 9-point Likert scale (1 = to a minor extent, 9 = to a very high degree). Consensus was predefined as ≥90% of respondents scoring a PSI as ≥6/9 (defining rating of ≥6/9 as “agreement”).^17^ PSIs endorsed by 75–89% of participants advanced to round 2 for re-evaluation. PSIs with an endorsement rate below 75% were excluded after round 1. Since different professional groups have different expertise, we expected that not every person would be able to accurately rate the relevance of all PSIs. Therefore, respondents could select “I cannot rate this item” for PSIs outside their expertise. PSIs were presented within their respective subcategories, and the order of subcategories was randomized. Furthermore, at the end of each subcategory free-text comment fields allowed participants to suggest revisions to PSI wording or propose new indicators. These were analysed by two study members (AB and MG), who drafted new PSIs or proposed adaptations accordingly. These were then discussed by the full team to reach consensus on the final wording.

Round 2 included all PSIs meeting the 75% relevance threshold from round 1, alongside newly proposed or revised PSIs. Participants received feedback on their own rating for relevance from round 1 and the median rating from all participants for all PSIs that were re-rated.

In addition, all PSI included in round 2 were evaluated regarding two characteristics: (1) Implementation feasibility, assessed through the question: “How easy is it to implement the measure/circumstance described here in a radiotherapy center (i.e. what are the costs and effort involved)?”. Responses were recorded on a four-point Likert scale: 1= very difficult to implement; 2= rather difficult to implement; 3= rather easy to implement; 4 = very easy to implement, with the option “I cannot rate” for cases where participants felt unable to provide an informed judgement. (2) Verifiability, assessed through the question: “How easily can the implementation of the described measure/circumstance be verified (e.g. how well is the information available)?” This was rated also on a four-point Likert scale: 1 = very difficult to check; 2 = rather difficult to check; 3 = rather easy to check; 4 = very easy to check, again including the option “I cannot rate ”. We analysed again the agreement rates regarding relevance of each PSI as well as the mean of the feasibility and verifiability rating. Inclusion of PSI in the pilot version of the PaSaRO was based solely on the relevance rating (> 90% in round 1 or round 2) and not influenced by other ratings or free text comments. All quantitative data was analysed combined for all participants without weighting or subgroup analyses.

#### Step 3.2: Checking comprehensibility in cognitive interviews

Between October and December 2024, cognitive interviews were conducted with professionals from RO facilities. All interviews were held via digital conference call and led by one team member (FS) using a semi-structured interview guide specifically developed for this purpose (see Appendix 5). Participants saw a presentation with one PSI including the response scale per slide. While all five professions described above were eligible, presented PSI were selected based on professions to fit their expertise. The response scale was designed as a six-point Likert scale ranging from 1 = “completely disagree” to 6 = “completely agree”, a response scale which was successfully used for several other questionnaires (for example^13,18^). Additionally, respondents were able to select “I cannot judge ” or “Not relevant for my department ” for any item. FS read out loud the PSI and participants were asked to rephrase those in their own words to evaluate the comprehensibility of the wording. Three interviews were based on the think-aloud-technique. Here participants were asked to assess the PSI for their own RO facility and explain how they derive their rating. This was done to check that participants not only understand the content of the PSI but also can apply the response scale. Two team members (AB and EC) independently reviewed all interview recordings and evaluated how well each PSI was understood and whether any suggestions for improvement were made. A PSI was considered final if it was rated as understandable in three consecutive interviews.^13^ If modifications were made to the wording based on misunderstandings or improvement suggestions, an additional three interviews confirming full understanding were required to finalize the PSI.

## RESULTS

### Sample

An overview of the distribution of professional groups in all study phases is shown in Table 1. All professional groups were represented, with the majority of participants being radiation therapists, medical physicists and physicians. Patient participants were 66% male and 44% female. They were treated for a range of tumour entities including breast cancer, prostate cancer and skin cancer. For more details on all samples, refer to Appendix 6.

**Table 1:**
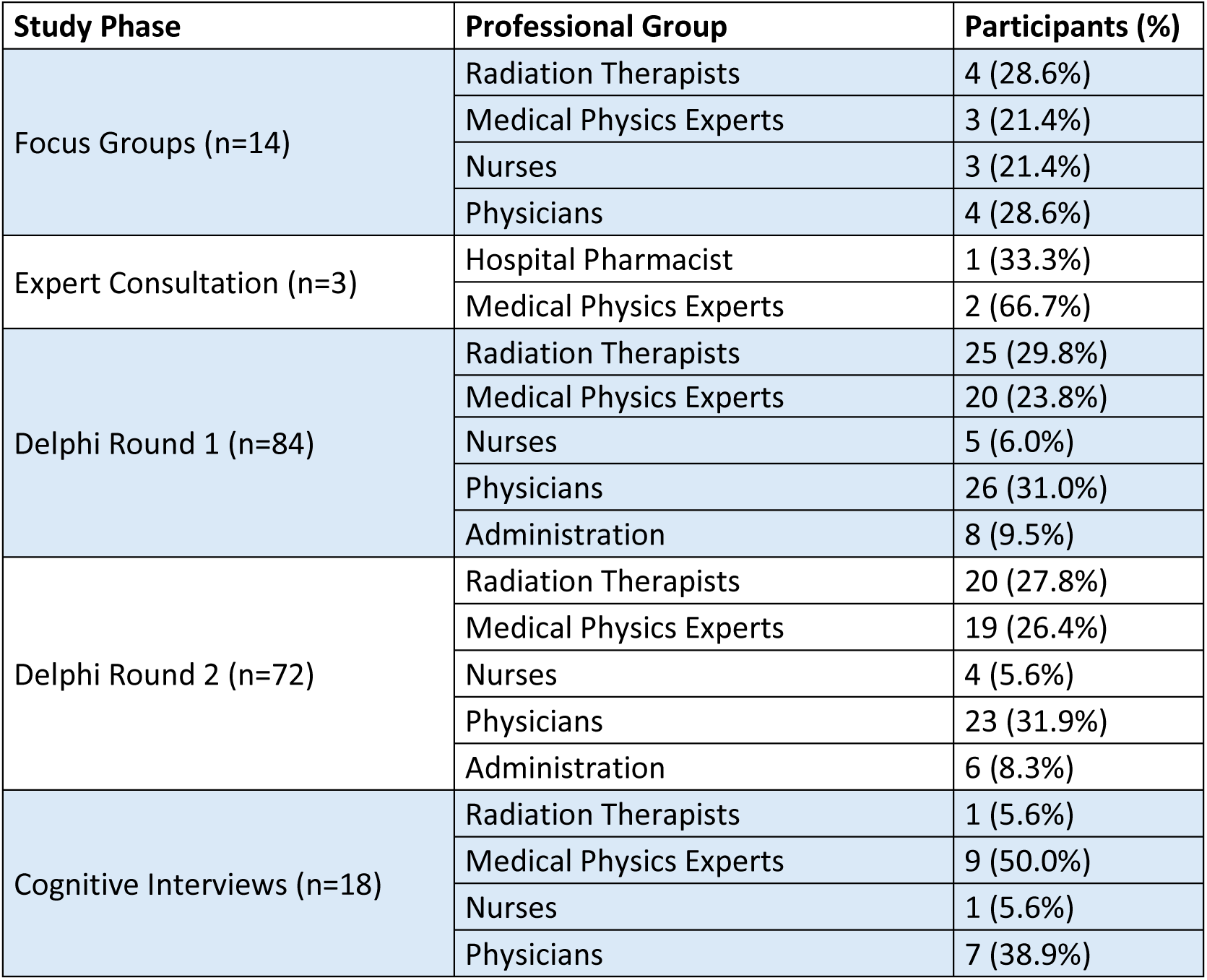
Overview of professional groups across all development steps.

| Study Phase | Professional Group | Participants (%) |
| --- | --- | --- |
| Focus Groups (n=14) | Radiation Therapists | 4 (28.6%) |
|  | Medical Physics Experts | 3 (21.4%) |
|  | Nurses | 3 (21.4%) |
|  | Physicians | 4 (28.6%) |
| Expert Consultation (n=3) | Hospital Pharmacist | 1 (33.3%) |
|  | Medical Physics Experts | 2 (66.7%) |
| Delphi Round 1 (n=84) | Radiation Therapists | 25 (29.8%) |
|  | Medical Physics Experts | 20 (23.8%) |
|  | Nurses | 5 (6.0%) |
|  | Physicians | 26 (31.0%) |
|  | Administration | 8 (9.5%) |
| Delphi Round 2 (n=72) | Radiation Therapists | 20 (27.8%) |
|  | Medical Physics Experts | 19 (26.4%) |
|  | Nurses | 4 (5.6%) |
|  | Physicians | 23 (31.9%) |
|  | Administration | 6 (8.3%) |
| Cognitive Interviews (n=18) | Radiation Therapists | 1 (5.6%) |
|  | Medical Physics Experts | 9 (50.0%) |
|  | Nurses | 1 (5.6%) |
|  | Physicians | 7 (38.9%) |

### Step 2: PSI generation

After focus groups 48 PSIs were added, 15 PSIs based on patient interviews and 12 PSIs based on expert consultation. In team discussion, seven items were merged with other items. The majority were added for the category Patient-specific Processes. For the categories Human Resources and Quality And Risk Management PSIs were only added based on focus groups. An overview of all generated items per category and method is displayed in Table 2.

**Table 2:**
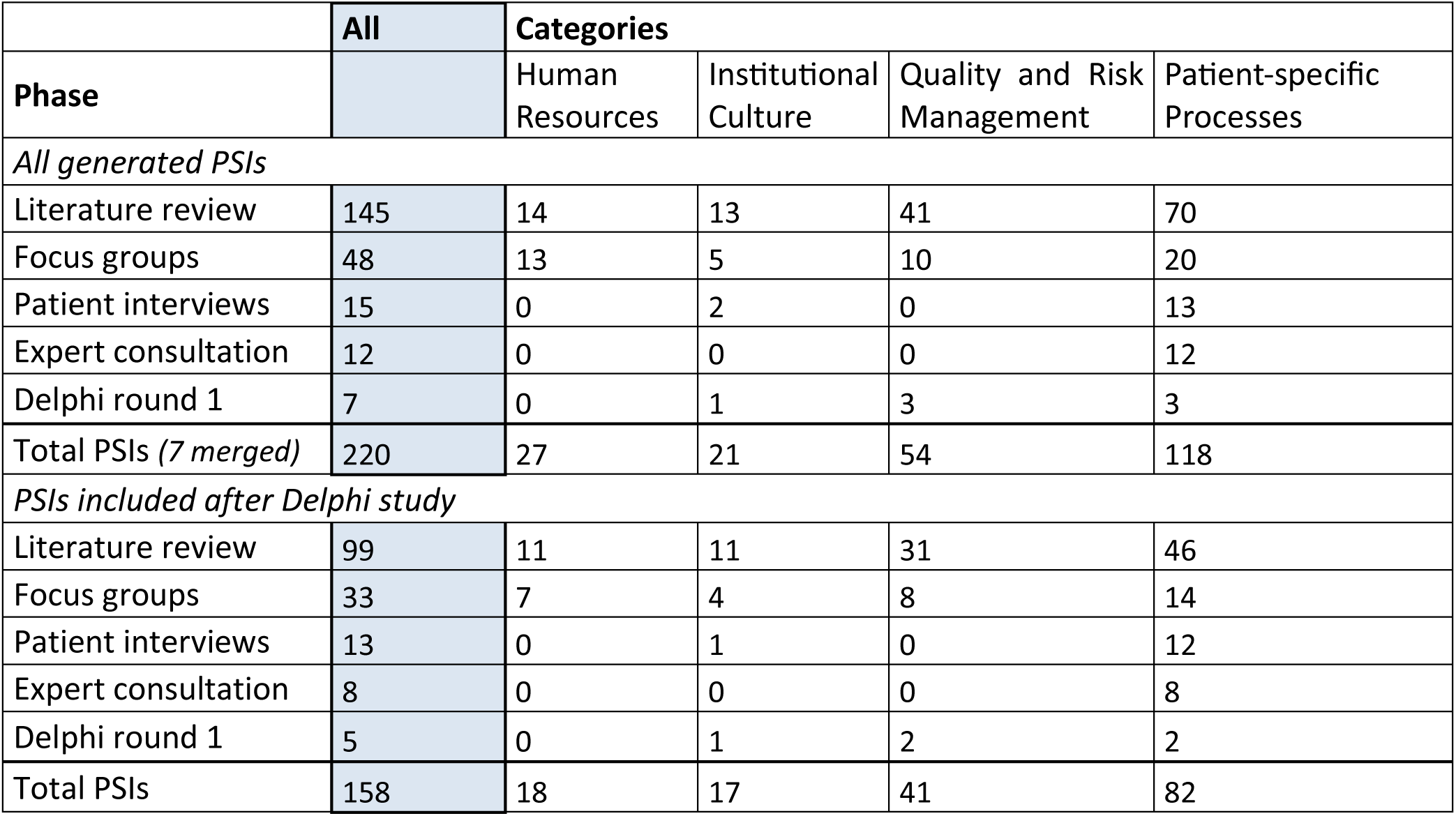
Distribution of patient safety indicators (PSIs) over categories and phase of generation.

|  | All | Categories |  |  |  |
| --- | --- | --- | --- | --- | --- |
| Phase |  | Human Resources | Institutional Culture | Quality and Risk Management | Patient-specific Processes |
| *All generated PSIs* |  |  |  |  |  |
| Literature review | 145 | 14 | 13 | 41 | 70 |
| Focus groups | 48 | 13 | 5 | 10 | 20 |
| Patient interviews | 15 | 0 | 2 | 0 | 13 |
| Expert consultation | 12 | 0 | 0 | 0 | 12 |
| Delphi round 1 | 7 | 0 | 1 | 3 | 3 |
| Total PSIs (7 merged) | 220 | 27 | 21 | 54 | 118 |
| *PSIs included after Delphi study* |  |  |  |  |  |
| Literature review | 99 | 11 | 11 | 31 | 46 |
| Focus groups | 33 | 7 | 4 | 8 | 14 |
| Patient interviews | 13 | 0 | 1 | 0 | 12 |
| Expert consultation | 8 | 0 | 0 | 0 | 8 |
| Delphi round 1 | 5 | 0 | 1 | 2 | 2 |
| Total PSIs | 158 | 18 | 17 | 41 | 82 |

### Step 3: PSI selection and testing of content validity

#### Step 3.1: Checking relevance and comprehensiveness in Delphi study

The two-round Delphi process for assessing PSI relevance started with an initial pool of 213 indicators (Figure 2). Detailed results for each PSI are shown in Appendix 7. In round 1, 76 PSIs (35.7%) achieved consensus. Another 75 PSIs (35.2%) received agreement ratings between 75-90% and were re-evaluated in round 2, while 25 PSIs were rephrased based on expert free-text feedback. Additionally, through free-text comments, the expert panel suggested seven new PSIs. The remaining 37 PSIs (17.4%) did not achieve the minimum consensus threshold of 75% and were excluded. Of those, 37 PSIs were generated based on the literature review, 7 based on focus groups, 3 based on expert consultation.

**Figure 2:**
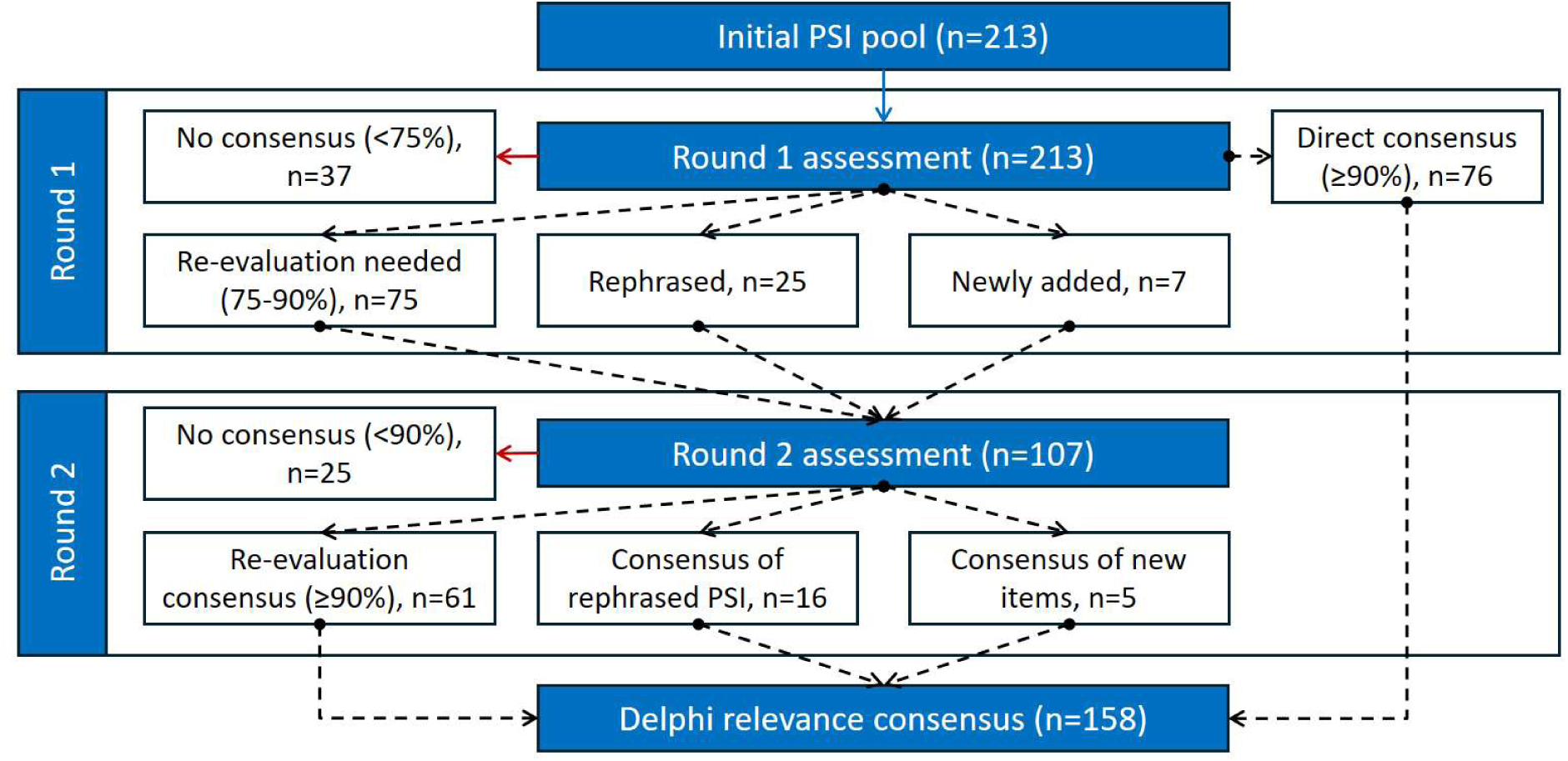
Flow diagram of the two-round Delphi process.

In round 2, 107 PSIs were evaluated for relevance. Of the 75 re-evaluated indicators, 61 PSIs achieved consensus. Additionally, 16 of the rephrased indicators and 5 of the newly phrased indicators were accepted. Twenty-five indicators failed to reach the 90% threshold and were excluded. The final pilot version after the relevance assessment consisted of 158 PSIs that achieved consensus.

The mean agreement rate (percentage of ratings of at least 6 out of 9) in round 1 was 84.92% (SD=12.83%, range: 37.50-100%). In round 2, with 107 evaluated PSIs, the mean agreement increased to 92.66% (SD=4.67%, range: 77.40-100%). For the 75 re-evaluated PSIs, the mean increase in agreement between rounds was 8.24% (SD=4.79%, range: -5.60-19.30%).

The results regarding implementation feasibility and verifiability are summarized in Table 3, showing category-specific differences in feasibility and ease of data collection ratings. The category Institutional Culture received the highest mean feasibility score (3.03 ± 0.26), while Human Resources had the lowest mean score (2.76 ± 0.30). In terms of ease of data collection, Patient-specific processes had the highest mean ratings (3.11 ± 0.22), whereas Institutional Culture received the lowest mean rating (2.83 ± 0.30).

**Table 3:**
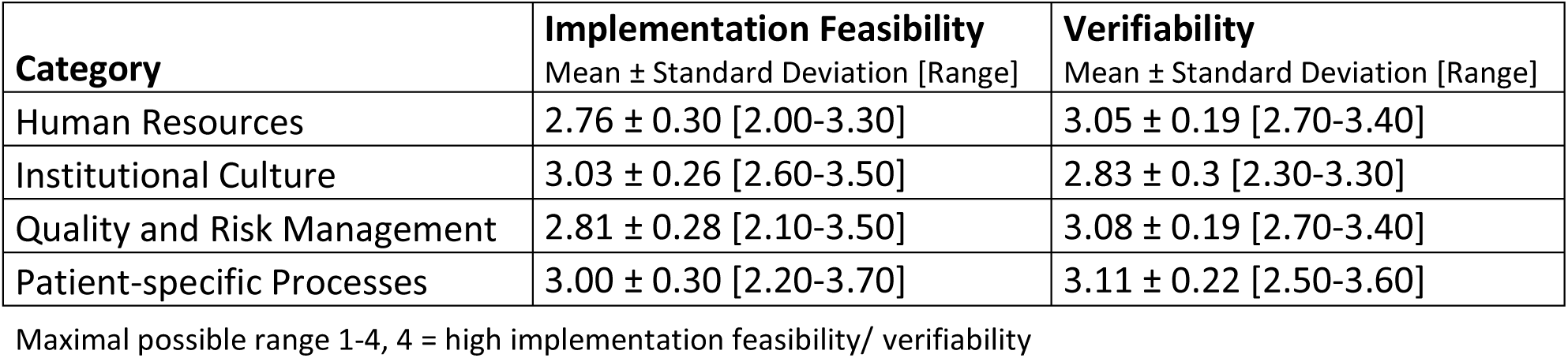
Implementation feasibility and verifiability (mean ± standard deviation (SD); range) across four categories.

| Category | Implementation Feasibility<br>Mean ± Standard Deviation [Range] | Verifiability<br>Mean ± Standard Deviation [Range] |
| --- | --- | --- |
| Human Resources | $2.76 \pm 0.30$ [2.00-3.30] | $3.05 \pm 0.19$ [2.70-3.40] |
| Institutional Culture | $3.03 \pm 0.26$ [2.60-3.50] | $2.83 \pm 0.3$ [2.30-3.30] |
| Quality and Risk Management | $2.81 \pm 0.28$ [2.10-3.50] | $3.08 \pm 0.19$ [2.70-3.40] |
| Patient-specific Processes | $3.00 \pm 0.30$ [2.20-3.70] | $3.11 \pm 0.22$ [2.50-3.60] |

Maximal possible range 1-4, 4 = high implementation feasibility/ verifiability

#### Step 3.2: Checking comprehensibility in cognitive interviews

Three rounds with a total of 18 cognitive interviews were required until all PSIs were well understood by participating RO professionals. 43 PSIs were linguistically optimized. For 12, only small adaptions were made whereas the formulation was changed in 31 PSIs. In addition, an instruction was added to explain the additional response categories “I cannot judge ” or “Not relevant for my department ” to improve comprehensibility.

### Excerpt from the pilot version of the PaSaRO

The pilot version of the PaSaRO includes a wide range of indicators covering various categories. Table 4 offers an overview about all categories and sub-categories deduced from PSIs in this study.

**Table 4:** Categorization of the 158 included patient safety indicators in the PaSaRO.

|  | Category | Count |
| --- | --- | --- |
| **1** | **Human Resources** | **18** |
| 1.1 | Promotion of continuous education and competency review | 5 |
| 1.2 | Staffing | 8 |
| 1.3 | Training: Courses for new technologies and procedures | 5 |
| **2** | **Institutional Culture** | **17** |
| 2.1 | Promotion of a safety culture | 8 |
| 2.2 | Internal feedback loops | 5 |
| 2.3 | Patient involvement | 4 |
| **3** | **Quality and Risk Management** | **41** |
| 3.1 | Existence and effectiveness of a system for error reporting and analysis | 8 |
| 3.2 | External peer review procedures | 2 |
| 3.3 | Internal peer review procedures | 10 |
| 3.4 | Control points and checklists | 9 |
| 3.5 | Failure of therapy-critical processes | 2 |
| 3.6 | Standard Operating Procedures (SOPs) | 6 |
| 3.7 | Technology assessment and risk analyses for new procedures | 4 |
| **4** | **Patient-specific Processes** | **82** |
| 4.1 | Selection and administration of tumor-effective systemic therapies | 7 |
| 4.2 | Treatment of patients with pacemakers and defibrillators | 3 |
| 4.3 | Radiation planning and plan evaluation | 9 |
| 4.4 | Contouring | 2 |
| 4.5 | Initial setup and subsequent irradiations | 5 |
| 4.6 | Patient presentation, informed consent, indication and prescription | 9 |
| 4.7 | Patient-specific quality assurance | 1 |
| 4.8 | Planning imaging and positioning | 6 |
| 4.9 | Prevention of adverse effects | 5 |
| 4.10 | Spatial and temporal organization of patient pathways | 7 |
| 4.11 | Interfaces between care providers | 7 |
| 4.12 | Symptom management | 14 |
| 4.13 | Therapy completion and follow-up care | 5 |
| 4.14 | Monitoring and adjustment of non-tumor-targeting medication therapies | 2 |
| **All** |  | **158** |

Table 5 provides a selection of examples translated into English, showcasing different categories, their acceptance rates in the Delphi process, and ratings for implementation feasibility and ease of data collection. These examples were selected to represent the range of rating results. A complete list of all PSIs, along with additional details, is provided in the Appendix 7.

**Table 5:** Exemplary patient safety indicators (PSIs) and ratings from the Delphi consensus process.

| Category - Subcategory | Patient Safety Indicator | Adapted after round 1 | Agreement round 1 | Agreement round 2 | Implementation feasibility | Verifiability |
| --- | --- | --- | --- | --- | --- | --- |
| Human resources - Staffing | The current staffing ratio appropriately reflects the actual departmental workload. Additional tasks of the team such as administration, research or teaching are taken into account appropriately. | no | 87.8 % | 90.3 % | M = 2.0 | M = 2.9 |
| Institutional culture - Involving the patients | Patients are encouraged to actively participate in the daily identification before therapy (e.g. make sure they give their name correctly). | Newly deduced from free texts | - | 93.4 % | M = 3.4 | M = 2.8 |
| Quality and risk management - Emergency concepts | There are written emergency concepts documented for the failure of therapy-critical processes. | no | 92.3 % | Consensus achieved in round 1 | M = 2.9 | M = 3.2 |
| Patient-specific processes - Planning and plan evaluation | The irradiation instruction/prescription is stored digitally in the irradiation planning system and is automatically adopted when the plan is created (e.g. fractionation and dose). | no | 100% | Consensus achieved in round 1 | M = 3.2 | M = 3.5 |
| Patient-specific processes - Patient presentation, information, indication and prescription | The patient's psychosocial situation and current psychological stress is asked about in the medical history | no | 77.8 % | 92.5 % | M = 3.0 | M = 2.9 |
| Patient-specific processes - Prophylaxis of adverse effects | There is a standard for prophylactic and therapeutic skin care during therapy. | no | 83.1 % | 94.1 % | M = 3.3 | M = 3.3 |
| Patient-specific processes - Management of symptoms | Every patient with <u>malignant</u> disease receives a clinical examination at least once a week. | yes (addition underlined) | 75.1 % | 95.0 % | M = 3.0 | M = 3.3 |

## DISCUSSION

The PaSaRO was developed in a rigorous multi-method study. Building on a systematic literature review, 82 additional PSIs were generated through focus groups with RO professionals and interviews with RO patients. In a Delphi study, 158 PSIs were identified as relevant and comprehensive. Their comprehensibility was then tested in cognitive interviews, ensuring that the pilot version of PaSaRO possesses strong content validity for measuring patient safety in RO departments from the professionals’ perspective. The PaSaRO reflects the multifaceted nature of patient safety in RO, and the Delphi ratings further demonstrated that most indicators are considered highly feasible to implement and verify.

We found no previously published indicator catalogues with a similarly broad methodological and stakeholder inclusion. For instance, while the American Association of Physicists in Medicine’s safety profile assessment (SPA) tool^5^ originated from multiple existing recommendations and was refined through a focus group, it involved only 24 participants and did not include the full spectrum of RO professionals.^19^ Similarly, the work of Vaandering et al.,^20^ Gabriele et al.^21^ and Lopez Torrecilla et al.^19^ on the development of national quality indicators, though valuable, relied primarily on literature reviews and small working groups. The Spanish study by Lopez Torrecilla et al.^19^ included a well-executed Delphi process among radiation oncologists, and Chargari et al.’s^22^ work on cervical cancer quality indicators involved 99 physicians in a Delphi approach, but both maintained a primarily physician-centric perspective. On the contrary, we included not only physicians, but five professional groups in our Delphi survey. Especially for our comprehensive approach with inclusion of all aspects of care during radiation treatment courses (including e.g., medication safety), it seems unreasonable to exclude perspective of nurses or administrative staff or even radiation therapists or physicists when rating relevance, feasibility or ease of data collection. Several interview or focus group studies with radiation therapists concerning safety in RO or nurses concerning safety in oncology in general can be found in literature, underlining a broad insight in safety aspects and willingness of these professional groups to talk about these topics.^21–23^ Besides, radiation therapists, nurses and administrative staff engage with RO patients longer during their pathway through radiation therapy than physicians and therefore experience daily examples for especially safe or unsafe processes during treatment delivery every day and with different perspective than physicians or medical physicists.^24,25^

Our comprehensive approach included the patients’ perspective on PS, which has not yet been taken into account in the development of any published PSI or quality indicator catalogue. The important role of patients as experts for their own healthcare safety was recently underlines by WHO and OECD in general healthcare^26,27^ Even though patients were described as very willing to participate in safety initiatives, there is often a lack in the structure of scientific studies to engage patients in a sufficient way ^28–30^ and initiatives which include patients are rare in general.^23^ Similar to other healthcare sectors,^31^ patients in RO have been described to monitor their own treatment as well as to contribute to safer treatment by reporting irregularities.^32^ The patient interviews proved to be an important source for the formulation of PSIs. Of 15 initially through the patient interviews created PSI, 13 gained consensus in the Delphi rounds which contain – as to expect – only aspects in the main category patient specific processes.

Concerning feasibility, our indicators in the human resources section were ranked lowest, suggesting that increasing staffing levels or improving training might face greater barriers or demands greater organizational commitment when compared to e.g., patient specific processes. Overall, we saw a high rating for verifiability in all our categories but lowest ratings for institutional culture, what might reflect the need for more nuanced or qualitative methods for evaluation of e.g., team dynamics. Nevertheless, the overall results reflect that these indicators are both feasible to adopt and straightforward to track.

### Strengths and limitations

A key strength of this development study is the inclusion of a wide range of stakeholders—not only five groups of RO professionals but also RO patients. This comprehensive approach ensures broad coverage of aspects relevant to patient safety in RO. Additionally, the use of multiple methodological steps based on a best practice example for questionnaire development ^13^ and the COSMIN reporting guideline^15^ ensures strong scientific rigor and transparent reporting of the development process of the PaSaRO.

Despite these strengths, several limitations must be noted. The relative underrepresentation of nurses and administrative staff might limit the PaSaRO’s ability to fully capture their perspectives on safety. Moreover, voluntary participation in the Delphi survey could also introduce bias, as those considering patient safety particularly important may have been more likely to participate and thus agreement rates might be increased.

### Implications

The 158 PSIs presented here form a pilot version of the PaSaRO. At this stage, only content validity has been established. The next steps include psychometric evaluation across all five professional groups. This evaluation will assess interrater reliability (consistency of ratings across professionals within the same department), test–retest reliability, and item-level characteristics (such as item difficulty and inter-item correlations). Based on these analyses, refinements may be made before finalizing the PaSaRO.

Once a psychometrically validated version of PaSaRO is available, implementation research will be essential. Key questions to address include the appropriate format for administering the tool should be applied, who should complete it, how often it should be used, and how the results should be communicated to staff. In addition, strategies will also be needed to translate findings into concrete safety improvements - whether through whole-team discussions or smaller task forces - and to monitor changes over time.

The final, validated PaSaRO has immediate potential for clinical implementation as a standardized tool to measure, monitor, and improve patient safety in RO. However, successful adoption will require adequate institutional support, as introducing numerous measures simultaneously could strain facilities with limited resources.

Beyond implementation, several research gaps remain. Linking PSI adherence to error rates (e.g., through incident learning systems such as ROSIS or SAFRON [38,39]) could provide evidence of predictive validity. Longitudinal studies are needed to determine whether PSI adoption leads to sustained reductions in adverse events. Additionally, international harmonization will be important to adapt PaSaRO to diverse healthcare systems, particularly in low-resource settings, where contextual feasibility may differ significantly.

## Conclusion

In this study, the PaSaRO was developed, the first comprehensive RO-specific questionnaire to assess patient safety in RO departments. The included 158 RO–specific PSIs were derived through a multi-methods approach—complementing indicators of a previous systematic literature review with focus groups, patient interviews, expert consultations, and a two-round Delphi survey with all major RO professional groups. The scientifically rigorous development process based on relevant guidelines and best practice examples ensured high content validity of the PaSaRO. The PSIs are organized into 4 main domains – Human Resources, Institutional Culture, Quality and Risk Management, and Patient-specific Processes– and 27 subcategories, ensuring thorough coverage of RO safety. Many indicators received high ratings in implementation feasibility and ease of data collection, indicating strong potential for enhancing patient safety when integrated into routine clinical practice.

### Ethics and registration

Ethical approval for this study was granted by the Hamburg Ethics Committee (Approval Number: 2023-101018-BO-ff). The trial is registered with the ARO (Arbeitsgemeinschaft Radioonkologie, the working group for radiation oncology of the German Cancer Society) under protocol number 2023-03 and is listed in the German Register for Clinical Trials (DRKS00034690).

## Declaration of generative AI and AI-assisted technologies in the manuscript preparation process

During the preparation of this work, the first authors used ChatGPT (GPT-5, OpenAI) to assist with English language proofreading and minor grammar improvements. The tool was used solely to enhance linguistic clarity; all scientific content, analyses, and interpretations were developed, verified, and validated by the author. After using this tool, the authors thoroughly reviewed and edited the text and take full responsibility for the content of the published article.

## Supporting information

Appendix 1 DELPHISTAR Reporting Guideline

Appendix 2 COSMIN Reporting Guideline

Appendix 3 Focus group interview guide

Appendix 4 Patient interview guide

Appendix 5 Cognitive interview guide

Appendix 6 Sample Characteristics

Appendix 7 Full list of PSIs

## Data Availability

All data produced in the present study are available upon reasonable request to the authors

## Acknowledgements

We would like to thank our patient advisory board, including Ines Moegling, Katrin Wemheuer, and Günther Carl, for their ongoing support and valuable feedback throughout the study.

