## Appendix 1 DELPHISTAR Reporting Guideline for "Development and content validity of the Patient Safety in Radiation Oncology questionnaire (PaSaRO): A multi-method study"

**Appendix 1: DELPHISTAR Reporting Guideline**
Article: Development and content validity of the Patient Safety in Radiation Oncology questionnaire (PaSaRO): A multi-method study
Authors: Maximilian Grohmann, Eva Christalle, Felicitas Schwenzer, Maria Jäckel, Nina Michalowski, Isabelle Scholl, Andrea Baehr

DELPHISTAR
Delphi studies in social and health sciences – recommendations for a standardized reporting

DeWiss Delphi techniques in health and social sciences

PH Schwäbisch Gmünd University of Education

### Delphi studies in social and health sciences – recommendations for an interdisciplinary standardized reporting (DELPHISTAR).

From: Niederberger, M., Schifano, J., Deckert, S., Hirt, J., Homberg, A., Köberich, S., Kuhn, R., Rommel, A., Sonnberger, M. & the DEWISS network (2024). Delphi studies in social and health sciences—Recommendations for an interdisciplinary standardized reporting (DELPHISTAR). Results of a Delphi study. *PLoS ONE 19(8):* e0304651. https://doi.org/10.1371/journal.pone.0304651

More information under OSF (https://osf.io/gc4jk) and DEWISS (https://delphi.ph-gmuend.de/)

**What is the aim of DELPHISTAR?**

- Improve, harmonize, and make the reporting in publications on Delphi studies comparable
- Facilitate the evaluation of Delphi studies including during peer review processes
- Reduce, and ideally prevent, inconsistencies and unclear descriptions in publications on Delphi studies
- Raise awareness of the diversity among the Delphi variants and of their specific potentials and challenges

**DELPHISTAR is a Delphi reporting guideline that is:**

- valid for all Delphi variants (e.g., classic Delphi, real-time Delphi, group Delphi, policy Delphi, argumentative Delphi, café Delphi)
- applicable to different purposes (e.g., Delphi studies to establish consensus, to gather expert judgments or to forecast)
- given equal consideration in the health and social sciences

**This reporting guideline is meant for studies using Delphi techniques in the health and social sciences.** These also include all Delphi variants and modifications that meet the following criteria:

1. Survey of several people (=experts) with specialized knowledge (e.g., operational knowledge, experiential knowledge, functional knowledge, contextual knowledge)
2. Structured communication process
3. Carrying out at least two survey rounds or the option to respond at least two times
4. Feedback: the (interim) results are presented to the experts starting from the second round
5. Basis is a quantitative questionnaire with the possibility to contribute or supplement arguments for the respective position
6. Quantitative and qualitative answer are systematically analyzed (quantitative: e.g., descriptive statistics, qualitative: e.g., thematic analysis)

This reporting guideline is available in English and German at https://delphi.ph-gmuend.de/activities/delphistar (last update October 2024).

**Contact**
Prof. Dr. Marlen Niederberger

Department of Research Methods in Health Promotion and Prevention
Institute for Health Sciences, University of Education Schwäbisch Gmünd,
Oberbettringer Straße 200, 73525 Schwäbisch Gmünd, Germany

### Appendix 1: DELPHISTAR Reporting Guideline

Article: Development and content validity of the Patient Safety in Radiation Oncology questionnaire (PaSaRO): A multi-method study
Authors: Maximilian Grohmann, Eva Christalle, Felicitas Schwenzer, Maria Jäckel, Nina Michalowski, Isabelle Scholl, Andrea Baehr

| Topic | Section | Item | Checklist Item | Location where item is reported | Exemplary wording |
|---|---|---|---|---|---|
| I Title and Abstract | | 1 | Identification as a Delphi study in the title | Not in the title, since it is just one of several methods | What is a public health intervention? Results of a Delphi study. |
| | | 2 | Identification as a Delphi study in the abstract | Abstract - Methods | A Delphi study was selected to answer the research question. |
| | | 3 | Structured abstract | Abstract | e.g., background, method, results and discussion |
| II Context | Formal | 4 | Information about the sources of funding | Funding | The Delphi study was funded by [SOURCE]. |
| | | 5 | Information about the team of authors and/or researchers (e.g., discipline, institution) | Methods - Study Design | The Delphi study was conducted by an interdisciplinary team with representatives from medicine, public health, and health promotion. |
| | | 6 | Information about method consulting | Methods - Study Design | The study group was advised by experts from [INSTITUTION] regarding statistics. Or: No consulting in regard to method took place. |
| | | 7 | Information about the project background | Methods - Study Design | The Delphi study was part of a mixed-methods study on [AIM]. |
| | | 8 | Information about the study protocol | Methods - Study Design | The study protocol is available at [LINK]. |
| | Content | 9 | Justification of the chosen method (Delphi) to answer the research question | Methods - Step 3.1: Checking relevance and comprehensiveness in Delphi study | The Delphi method is suitable for answering the research question because it systematically gathers the judgments of different expert groups and can identity agreement and disagreement. |
| | | 10 | Aim of the Delphi study (e.g., consensus, forecasting) | Methods - Step 3.1: Checking relevance and comprehensiveness in Delphi study | The aim of the Delphi study is to find consensus on criteria to define a public health intervention. |
| III Method | Body & Integration of knowledge | 11 | Identification and elucidation of relevant expertise, spheres of experience, and perspectives (e.g., theory, practice, affected groups, disciplines) | Methods - sample | The experts represent the sciences and clinical practice because [REASON]. |
| | | 12 | Handling of knowledge, expertise and perspectives which are missing or have been deliberately not integrated | All relevant professional groups were included. | If it is not possible to recruit experts specialized in [AREA], this will be openly communicated to the other experts during the Delphi study. |
| | | 13 | Basic definition of expert[1] | Methods - Step 3.1: Checking relevance and comprehensiveness in Delphi study | A person who has been active in the area for at least [NUMBER] years is considered to be an expert. |
| | Delphi variant and modifi-cations | 14 | Identification of the type of Delphi variant and potential modifications (e.g., classic Delphi, real-time Delphi, group Delphi) | Methods - Step 3.1: Checking relevance and comprehensiveness in Delphi study | A classic Delphi study was used [LITERATURE REFERENCE]. |
| | | 15 | Justification of the Delphi variant and modifications, | Not applicable | If the willingness to participate clearly decreases between the first and second round, a third round will not be held. |

**Appendix 1: DELPHISTAR Reporting Guideline**
Article: Development and content validity of the Patient Safety in Radiation Oncology questionnaire (PaSaRO): A multi-method study
Authors: Maximilian Grohmann, Eva Christalle, Felicitas Schwenzer, Maria Jäckel, Nina Michalowski, Isabelle Scholl, Andrea Baehr

| Topic | Section | Item | Checklist Item | Location where item is reported | Exemplary wording |
|---|---|---|---|---|---|
| | | | including during the Delphi study, if applicable | | |
| | **Sample of experts** | 16 | Selection criteria for the experts (per round, per expert group if applicable) | Methods - Sample | All of the experts who met the definition were invited to the first round. All of the experts who completed the previous round were invited to participate in the subsequent round. |
| | | 17 | Identification of the experts | Methods - Sample | The experts were identified based on publications in [DATABASE]. |
| | | 18 | Information about recruiting and any subsequent recruiting of experts | Methods - Sample | The experts were informed about the Delphi study and invited to participate. |
| | **Survey** | 19 | Elucidation of the content development for the questionnaire[2] | Methods - Section Step 2: PSI generation | The questionnaire was developed based on the results of systematic reviews [LITERATURE REFERENCE]. |
| | | 20 | Description of the questionnaire (content and structure) | Methods - Step 3.1: Checking relevance and comprehensiveness in Delphi study | The questionnaire was divided into three segments on [TOPICS]. The statements made in the questionnaire were evaluated using standardized items, with the option to comment in free-text boxes. |
| | **Delphi rounds** | 21 | Number of Delphi rounds | Methods - Step 3.1: Checking relevance and comprehensiveness in Delphi study | Three Delphi rounds were held. |
| | | 22 | Information about the aims of the individual Delphi rounds | Methods - Step 3.1: Checking relevance and comprehensiveness in Delphi study | The first Delphi round focused on exploring relevant aspects. These aspects were then presented to the experts in the second Delphi round for standardized evaluation. |
| | | 23 | Disclosure and justification of the criterion for discontinuation | Not applicable | The number of rounds was defined in advance to be a maximum of three rounds. |
| | **Feedback** | 24 | Information about what data was reported back per round | Methods - Step 3.1: Checking relevance and comprehensiveness in Delphi study | In terms of feedback, we shared the statistical results plus the summary of the open responses. |
| | | 25 | Information on how the results of the previous Delphi round were fed back to the experts surveyed (e.g., via frequencies, mean values, measures of dispersion, listing of comments) | Methods - Step 3.1: Checking relevance and comprehensiveness in Delphi study | Mean values, standard deviations and percentage frequency distributions were reported. |
| | | 26 | Information on whether feedback was differentiated by specific groups (e.g., by field of expertise, institutional affiliation) | Methods - Step 3.1: Checking relevance and comprehensiveness in Delphi study | The feedback was aggregated across all expert groups. |
| | | 27 | Information about how dissent and unclear results were handled | Methods - Step 3.1: Checking relevance and comprehensiveness in Delphi study | The results showing dissent were presented again for evaluation in the next Delphi round. |

**Appendix 1: DELPHISTAR Reporting Guideline**

Article: Development and content validity of the Patient Safety in Radiation Oncology questionnaire (PaSaRO): A multi-method study

Authors: Maximilian Grohmann, Eva Christalle, Felicitas Schwenzer, Maria Jäckel, Nina Michalowski, Isabelle Scholl, Andrea Baehr

| Topic | Section | Item | Checklist Item | Location where item is reported | Exemplary wording |
|---|---|---|---|---|---|
| | **Data analysis** | 28 | Disclosure of the quantitative and qualitative analytical strategy | Methods - Step 3.1: Checking relevance and comprehensiveness in Delphi study | The quantitative items were descriptively analyzed. The open-ended items were analyzed using thematic analysis [LITERATURE REFERENCE]. |
| | | 29 | Definition and measurement of consensus | Methods - Step 3.1: Checking relevance and comprehensiveness in Delphi study | Consensus was defined as percentage agreement, meaning that agreement was assumed if at least 80% of the respondents agreed on an item. |
| | | 30 | Information on group-specific analysis or weighting of experts (e.g., theory vs. practice, discipline-specific analysis) | Methods - Step 3.1: Checking relevance and comprehensiveness in Delphi study | In the analysis, the mean values for percent agreement are weighted for each expert group in terms of the number of group members. |
| IV Results | **Delphi process** | 31 | Illustration of the Delphi study (e.g., in a flow chart) | Figure 2 | A summary of the Delphi study is illustrated in a flow chart (Figure 1). |
| | | 32 | Information about special aspects during the Delphi study (e.g., deviations from the intended approach with justification) | Not applicable | During the Delphi study the political discussion mentioned climate change and the effects on health. It is possible that this influenced the experts' responses. |
| | | 33 | Number of experts per round (both invited and participating) | Results – Sample and Table 1 | The number of experts participating in the first Delphi round was [NUMBER], and the number of experts in the second round was [NUMBER]. This corresponds to a response rate of [NUMBER]% in the first round and [NUMBER]% in the second round. |
| | **Results** | 34 | Presentation of the results for each Delphi round and the final results | Results - Step 3.1: Checking relevance and comprehensiveness in Delphi study | In the first Delphi round [NUMBER]% of the experts agreed, in the second [NUMBER]%, and in the third [NUMBER]%. |
| V Discussion | **Quality of findings** | 35 | Highlighting the findings from the Delphi study | Discussion first paragraph | The central findings can be summarized as follows: [STATE FINDINGS]. |
| | | 36 | Validity of the results (e.g., transferability of the findings) | Not discussed since the Delphi study is just one part of a broader study | The results are not transferable to other countries due to different legal regulations. |
| | | 37 | Reliability of the results (e.g., split half, inter-rater reliability) | Not discussed since the Delphi study is just one part of a broader study | The responses in the free-text comments were analyzed by two independent reviewers [SPECIFY]. |
| | | 38 | Reflection on potential limitations (e.g., number of experts, response bias) | Discussion – Strengths and limitations | The results are to be viewed critically with regard to the composition of the panel because [REASONS]. |

[1] "Experts" are the participants; these can be people from academia, practice, or representatives of lived experience (e.g., patients, family members).

[2] The term "questionnaire" stands for the survey instrument regardless of whether quantitative or qualitative items are integrated or weighted.
