## Appendix 2 COSMIN Reporting Guideline for "Development and content validity of the Patient Safety in Radiation Oncology questionnaire (PaSaRO): A multi-method study"

**Appendix 2: COSMIN Reporting Guideline**
Article: Development and content validity of the Patient Safety in Radiation Oncology questionnaire (PaSaRO): A multi-method study
Authors: Maximilian Grohmann, Eva Christalle, Felicitas Schwenzer, Maria Jäckel, Nina Michalowski, Isabelle Scholl, Andrea Baehr

### The COSMIN reporting guideline for studies on measurement properties of PROMs - Version 2.0

Please refer to this document as:

JJ Gagnier, et al. COSMIN reporting guideline for studies on measurement properties of patient-reported outcome measures: version 2.0. Quality of Life research. 2025. https://doi.org/10.1007/s11136-025-03950-x

Information about how to apply the guideline is provided:

Arruda GT, et al. Explanation & Elaboration document of the COSMIN Reporting Guideline 2.0 for studies on measurement properties of patient-reported outcome measures. Quality of Life Research, 2025. Doi: 10.1007/s11136-025-03949-4

| **Item number – item name** | **Item description** | |
|---|---|---|
| **Report section: Title** | | |
| T1 – Title | Identify the report as a study of one or more measurement properties of a specific PROM to measure a specified construct in a specified population. | Title includes “Development and content validity of the Patient Safety in Radiation Oncology questionnaire (PaSaRO)” |
| Report section: Abstract | | |
| A1 – Objectives | Provide the specific objective(s) of the research, specifying (1) the name (and version, if relevant), and construct(s) of the PROM, (2) the measurement properties being evaluated, and (3) relevant study characteristics. | See Abstract - section *Background* and *Methods* |
| A2 – Design | Specify (details of the) study design used to evaluate the measurement properties. | See Abstract - section *Methods* |
| A3 – Methods | Specify the methods for evaluating each measurement property. | See Abstract - section *Methods* |
| A4 – Results | Provide the main results for all measurement properties evaluated. | See Abstract - section *Findings* |
| A5 – Discussion/Conclusions | Provide a brief statement of the implications of the findings in the context of existing evidence on the PROM. | See Abstract - section *Interpretation* |
| **Report section: Introduction** | | |
| I1 – PROM | Specify the name and, if relevant, the version, and construct(s) of the PROM. | See Introduction, last paragraph for the name and second to last for a definition |
| I2 – Target population | Specify the target population and context of use that the PROM was designed for. | See Introduction, last paragraph |

**Appendix 2: COSMIN Reporting Guideline**

Article: Development and content validity of the Patient Safety in Radiation Oncology questionnaire (PaSaRO): A multi-method study

Authors: Maximilian Grohmann, Eva Christalle, Felicitas Schwenzer, Maria Jäckel, Nina Michalowski, Isabelle Scholl, Andrea Baehr

| | | |
|---|---|---|
| I3 – State of knowledge & Rationale | Provide a description of the current scientific knowledge (what is known and not known) regarding the measurement properties of the PROM. Explain why the new study is necessary. Provide citations for the original development paper(s). | See Introduction, second paragraph |
| I4 – Objectives | Provide the specific objective(s) of the research, specifying (1) the name (and version, if relevant) of the PROM, (2) the measurement properties being evaluated, and (3) relevant study sample characteristics. | See Introduction, last paragraph |
| **Report section: General Methods** | | |
| GM1 – Study design | Specify (details of the) study design used to evaluate the measurement properties. | See Methods, section *study design* |
| GM2 – Participants | Specify how the study participants were selected. Specify the inclusion and exclusion criteria | See Methods, section *sample* |
| GM3 – PROM details | Provide details about the original version of the PROM as well as of the PROM version being studied, specify the conceptual framework (reflective/formative model), details on the structure (the number of items and subscales), the language, response scale, recall period, direction of scoring, and scoring algorithm of the PROM. Specify how the PROM was administered (e.g., in what setting, mode of administration (e.g. paper, electronic) what instructions were given), including the country in which it is administered | This paper describes the development of the original version. Subscales are based on previous work described in Methods - section *Step 2.1: Literature review*. The number of items are a result of this study and therefore described in Results. Further details like the conceptual model are described in Methods - section *Step 1: Preparation.* |
| GM4 – Additional data collection | Describe why and how other data was collected (e.g., construct and measurement properties of the comparator instruments, characteristics of groups being compared, and rationale for choosing groups), including mode of administration (e.g., paper, electronic). | See Methods, where for each step data collection is described in detail |
| GM5 – Time points procedures | Provide all time points of all measurements. | See Methods, where for each step data collection is described in detail |
| GM6 – Justification for sample size | Provide a rationale for the sample size for all measurement properties analyses (including subgroups). | The only MP tested here is content validity. For Delphi study … For cognitive interviews the rationale for the sample size is described in Methods - *Step 3.2: Checking comprehensibility in cognitive interviews.* |
| GM7 – Statistical analyses | Describe the statistical analyses corresponding to all objectives (see measurement properties specific boxes). Describe the criteria for good | See methods – section *Step 3.1: Checking relevance and comprehensiveness in Delphi study* and section |

| | | |
|---|---|---|
| | measurement properties. Name the statistical package used and the version. | *Step 3.2: Checking comprehensibility in cognitive interviews* |
| GM8 – Missing data | Describe approaches for dealing with missing data. | Not applicable. |
| GM9 – Unplanned analysis | Specify analyses that were unplanned and their rationale. | Not applicable. |
| **Report section: General results** | | |
| GR1 – Participant characteristics | Provide study participants' characteristics, specified per subgroup if applicable. | See Results – section *Sample* and Appendix X |
| GR2 – Sample size | Provide the total number of participants included in the study and the sample size for each analysis. | See Results – Table 1 |
| GR3 – Missing data | Provide amount of (proportion or count) and reasons for missing data for each analysis for the PROM, and for any analyses of other outcome measurement instruments. | Not applicable |
| GR4 – Results | Describe the results corresponding to all objectives (see measurement properties specific boxes). | See Results in the corresponding sections |
| **Report section: Discussion/conclusions** | | |
| DC1 – Measurement property evidence | Provide the main findings and if each measurement property is sufficient or insufficient and why. | See Discussion – first paragraph |
| DC2 – Practical relevance | Discuss the practical relevance of the findings in terms of recommendations for (not) using the PROM. | See Discussion – section *Implications* |
| DC3 – Strengths and limitations | Discuss strengths and limitations of each study. For example, discuss if there were any potential biases in the study that could have impacted the results. | See Discussion – section *Strengths and limiations* |
| DC4 – Generalizability | Discuss generalizability of the results. For example, discuss whether the results could be generalized to other populations given the sample studied. | See Discussion – section *Strengths and limiations* |
| DC5 – Instrument changes | Discuss what modifications are needed to the existing PROM. | No modifications yet required, but psychometric validation is necessary. See Discussion – section *Implications* |
| DC6 – Future research | Describe new research questions or hypotheses generated from these findings, and provide/describe the research needed to answer those questions. | See Discussion – section *Implications* |
| DC7 – Conclusions | Provide the overall conclusions for the use of the PROM. | See Discussion – section *Conclusions* |
| **Report section: Other information** | | |

**Appendix 2: COSMIN Reporting Guideline**

Article: Development and content validity of the Patient Safety in Radiation Oncology questionnaire (PaSaRO): A multi-method study

Authors: Maximilian Grohmann, Eva Christalle, Felicitas Schwenzer, Maria Jäckel, Nina Michalowski, Isabelle Scholl, Andrea Baehr

| | | |
|---|---|---|
| O1 – Conflict of interest | State any conflict of interest you may have related to the PROM. This may include any involvement in the development of the PROM or any commercial funding or profit. | See Conflict of interest |

**Appendix 2: COSMIN Reporting Guideline**

Article: Development and content validity of the Patient Safety in Radiation Oncology questionnaire (PaSaRO): A multi-method study

Authors: Maximilian Grohmann, Eva Christalle, Felicitas Schwenzer, Maria Jäckel, Nina Michalowski, Isabelle Scholl, Andrea Baehr

| Specific Reporting recommendations for studies on Content Validity | | |
|---|---|---|
| **Item number – item name** | **Item description – Second version** | |
| **Content validity: Methods** | | |
| CV1 – Relevance | Specify if and how patients and/or professionals were asked whether the instructions, each of the items, response options, and the recall period were relevant for the construct, population, and context of use. | See Methods - section *Step 3.1: Checking relevance and comprehensiveness in Delphi study* |
| CV2 – Comprehensiveness | Specify whether and how patients and/or professionals were asked whether all key concepts are included in the PROM. | See Methods - section *Step 3.1: Checking relevance and comprehensiveness in Delphi study* |
| CV3 – Comprehensibility | Specify whether and how the comprehensibility of the PROM instructions, items, response options, and recall period was evaluated by patients and/or professionals. | See Methods - *Step 3.2: Checking comprehensibility in cognitive interviews* |
| **Content validity: Results** | | |
| CV4 – Relevance | Specify if the instructions, all items, response options, and recall period were considered relevant, by patients and/or professionals, to the construct, population, and context of use. | See results - *Step 3.1: Checking relevance and comprehensiveness in Delphi study* |
| CV5 – Comprehensiveness | Specify whether patients and/or professionals considered all key concepts to be included in the PROM. | See results - *Step 3.1: Checking relevance and comprehensiveness in Delphi study* |
| CV6 – Comprehensibility | Specify whether patients understood the PROM instructions, items, response options, and recall period as intended and/or whether professionals considered the instructions, items, response options, were appropriately worded. | See results - *Step 3.2: Checking comprehensibility in cognitive interviews* |
