## Appendix 3 Focus group interview guide for "Development and content validity of the Patient Safety in Radiation Oncology questionnaire (PaSaRO): A multi-method study"

**Appendix 3: Focus group interview guide**
Article: Development and content validity of the Patient Safety in Radiation Oncology questionnaire (PaSaRO): A multi-method study
Authors: M. Grohmann, E. Christalle, F. Schwenzer, M. Jäckel, N. Michalowski, I. Scholl, A. Baehr

### Guideline focus groups

**Purpose**

The aim of the focus groups is to derive new patient safety indicators (PSIs) or further develop existing ones during a moderated group discussion among experts in the field of radiation oncology.

The two groups consist of 6 to 9 experts in the field of radiation oncology (physicians, medical physicists, nurses, administrative staff and radiation therapists) who meet for a 6-hour workshop that includes discussions on patient safety in radiation oncology. The discussion is moderated and supported by PowerPoint slides with examples of PSIs deduced from a literature review in four categories. For all categories, specific topics were identified by the study team that were not sufficiently evaluated regarding patient safety in the literature review. The moderator will introduce those topics to the group with the aim of deriving relevant PSIs (topic-specific parts).

The discussion is recorded and additional notes are written down by a student research assistant (FS). The recording will be transcribed afterwards. All personal data (e.g., information on certain hospitals or names) will be deleted to create an anonymous transcript. The transcript will be analysed and discussed by two members of the study team to deduce PSIs from the content or to adapt existing ones.

**Task of moderation**

- Moderation by a psychologist (EC) who is not familiar with the participants to ensure neutrality
- Additional moderation by a radiation oncologist (AB) to ensure that relevant topics are covered in depth
- Facilitate discussion between participants
- Lead discussion based on the 4 categories (human resources, institutional culture, quality and risk management, and patient-specific processes)
- Ensure focus on purpose of focus group
- Create a pleasant, respectful atmosphere
- Keep your own opinions out, be curious but neutral

**Examples for moderation**

- Ask for reactions in the group on a topic: What are your views on this? What are your experiences? What do you think about it? Does anyone have any other experiences/ideas?
- Encourage all professional groups to participate in the discussion: How do you see this as a (professional function)?
- Steer back if the discussion strays too far from PS: That's an exciting topic. What role does it play in patient safety / How does it relate to patient safety? That's also an interesting topic, but I have the impression that we've strayed away thematically from PS. Can we leave the topic as it is and focus on patient safety / sub-topic XY again?

- In-depth questions, e.g. to understand something better: I haven't quite understood that yet. What do you mean by ...? or descriptive questions: Do I understand correctly that you mean ...?
- Always lead to specific formulations for indicators: Can you describe in more detail what exactly needs to happen to ensure patient safety at this point? / What specific measures are needed at this point to make it safe for patients?
- Ask about your own experiences: How is this implemented in your clinic?

**PROCEDURE**

**1. Welcome and round of introductions (AB and EC):**

- Team introduces itself
- All participants in turn: name, professional function, workplace (medical centre and city)
- Information about audio recordings (data are analysed anonymously, the audio recording is transcribed, personal data are deleted)
- Additional bullet point protocol written down by student research assistant (FS)
- Do you consent? --> Start recording

**2. Introduction to the topic (AB):**

- Introducing the purpose of the focus group
  - Definition of patient safety
  - Definition of patient safety indicators
  - Brief description of the PaSaGeRO study, including next steps
- Present the role of the moderator
  - ➢ We discuss different areas for PSIs. The moderator leads through the topics and sometimes asks specific questions. In general, we would like you to have an open discussion.
  - ➢ The moderator only steers the discussion, e.g. to discuss topics in more depth and ensure the focus is on our research questions.
  - ➢ The important thing is that you don't have to agree. We are just as interested in agreement and consensus as we are in differing opinions.
  - ➢ There is no 'right' or 'wrong', it's about your own perspectives from your different areas of expertise.
- Instruction to improve recording quality
  - ➢ Try to speak one after the other. This is sometimes difficult, but if several people speak at the same time, it is difficult to understand on the recording.

**Appendix 3: Focus group interview guide**
Article: Development and content validity of the Patient Safety in Radiation Oncology questionnaire (PaSaRO): A multi-method study
Authors: M. Grohmann, E. Christalle, F. Schwenzer, M. Jäckel, N. Michalowski, I. Scholl, A. Baehr

**Introductory question (EC)**
Objective: Establish a connection to the topic
Procedure: Participants receive index cards and pencils. The moderator gives an instruction and all participants note their thoughts and suggestions. A maximum of two cards are discussed with the aim of jointly formulating an indicator so that it can be easily evaluated.

- ➢ Instruction: Spontaneously write down one aspect that particularly jeopardizes patient safety in your professional field and one aspect that particularly ensures patient safety.
- ➢ Who would like to present a card?
- ➢ What could a PSI look like that you can easily rate?

**3. Discussion category human resources (EC)**

**General part:**

- One slide per topic area with definition and 1-2 examples PSIs derived from the literature review
- ➢ What other indicators can you think of in this area?
- ➢ What specific measures do you take in this area in your department?

**Category-specific part (if there is time):**

- ➢ What role does training for staff play in patient safety?
- ➢ Which training courses for employees do you think should be mandatory?
- ➢ For which techniques must continuous training and/or competence checks take place? (Examples: IGRT, stereotaxy)
- ➢ What experience do you have with key users, i.e. people who become experts in a technique and train others?
- ➢ What other ways are there to implement training content in a department?

**4. Discussion category institutional culture (EC)**

**General part:**

- One slide per topic area with definition and 1-2 examples PSIs derived from the literature review
  - ➢ What other indicators can you think of in this area?
  - ➢ What specific measures do you take in this area in your department?

**Category-specific part (if there is time):**

- ➢ What needs to be ensured to establish a positive error culture?
- ➢ What are the obstacles for employees to speak openly about safety concerns and errors?
- ➢ What are beneficial measures for increasing a constructive safety culture?
- ➢ What role does leadership play in establishing a safety culture?

5. Discussion on quality and risk management (EC)

**General part:**

- One slide per topic area with definition and 1-2 examples PSIs derived from the literature review
  - What other indicators can you think of in this area?
  - What specific measures do you take in this area in your department?

**Category-specific part (if there is time):**

- How do error reporting and learning systems need to be structured and established to be as effective and efficient as possible? (What features do they need?)
- What are your experiences with systems that apply to the whole medical centre compared to intra-departmental systems? Where do you see advantages and disadvantages?
- What measures are important when introducing new therapies or procedures?
- What role do certifications play for the patient safety of therapies?
- What can be certified?
- What should be certified?
- Do radiation therapy-specific certifications bring benefits?

**6. Discussion of patient-specific processes (EC)**

**General part:**

- One slide per topic area with definition and 1-2 examples PSIs derived from the literature review
  - What other indicators can you think of in this area?
  - What specific measures do you take in this area in your department?

**Category-specific part (if there is time):**

- Are there any special queries and/or contraindications for brachytherapy?
- How should we proceed to ensure that prior radiation therapy is reliably queried and taken into account? (*We do not yet have a PSI for this. Discuss in detail and specifically so that we can create PSIs.*)
- There are many recommendations for specific tumours regarding how long it should take to start treatment. What options are there to ensure that this is adhered to? What guidelines do you follow? Who ensures this? Where are the pitfalls?
- In your opinion, in which cases would it be necessary to refer patients to other clinics with different treatment options and expertise? Do you have any such cases? Do you have specific criteria for this?
- How is therapy determined for older patients? What standards do you have? For example, do you use scoring tools before chemotherapy or radiotherapy? (*We do not yet have a PSI for this. Discuss in detail and specifically so that we can create PSIs.*)
- How are patients screened and treated for malnutrition, pain, and nausea? What role do scoring tools play in your work in this context? What advantages and disadvantages do you see in specifying specific scores in the PSI compared to the general wording "It is systematically recorded..."?

- What are the risks involved in radiation planning? How are these effectively mitigated? (*So far, we have few PSIs on radiation planning, apart from peer reviews. Not everything can be identified in a peer review. However, much is also regulated in DIN standards.*)
- What role do national or international training courses and/or competitions play for employees? Is training at the European level preferable to national training? Do competitions, e.g., for radiation plans, add value?
- What are the special considerations for patients undergoing combined radioimmunotherapy? E.g., during therapy or follow-up care?
- There are many recommendations on how to safely treat patients with cardiac devices (e.g., switching off the device, performing an ECG during or after radiation therapy, etc.). What specific measures do you take? Which measures do you consider particularly essential? Who is responsible for risk stratification? How is stratification and monitoring carried out?

**7. Conclusion of the discussion on PSIs (EC)**
- What important topics have we not yet discussed? / Have we forgotten anything important?

**Final round (AB)**
- Flashlight: What are your take-home messages of this day? What surprised you? What else would you like to share with us?
- Acknowledgements

**Debriefing of the focus group**
- We discuss and write down our impressions - What went well? What could have gone better? Were there any noticeable group dynamics?
