## Appendix 4 Patient interview guide for "Development and content validity of the Patient Safety in Radiation Oncology questionnaire (PaSaRO): A multi-method study"

**Appendix 4: Patient interview guide**
Article: Development and content validity of the Patient Safety in Radiation Oncology questionnaire (PaSaRO): A multi-method study
Authors: M. Grohmann, E. Christalle, F. Schwenzer, M. Jäckel, N. Michalowski, I. Scholl, A. Baehr

### Interview guide for semi-structured telephone interviews with patients

#### Purpose

The aim of the interviews is to shed light on patient safety in radiotherapy from the patient's perspective. The interview guide consists of main questions with optional follow-up questions (semi-structured interviews). The interviewer (FS) ensures that all topics are addressed but adapts the order of the main questions and follow-up questions to the course of the interview if necessary. The interviewer's attitude is interested, open, neutral and appreciative.
The interviews are conducted by telephone by a student research assistant with a background in educational science and medical psychology. The interviews are scheduled for a maximum of one hour (longer only if the participants explicitly request it).

All participants received written study information and gave informed consent prior to the interview.

**Introduction (5 min)**

- ➢ As you have already read, the interview will be recorded. After the interview, we will transcribe the recordings, i.e. we will write down what was said. In doing so, we will delete all information that would allow conclusions to be drawn about your person. For example, we delete all names of people or hospitals and practices.. In this way, the transcript is anonymized at the end. After transcription, we delete the recordings and only work with the anonymized transcript for data analysis.
- ➢ Do you agree to this? [Wait for consent] Then I will start the recording now.
- ➢ We are conducting this interview as part of the PaSaGeRO study. Our aim is to promote patient safety in radiotherapy. To this end, we are developing a questionnaire for healthcare professionals to provide clinics and practices. with feedback on the current status of patient safety and possible improvement measures. I would therefore like to shed some light on your perspective on the topic of patient safety. I will ask you some questions about your experiences in radiotherapy. In doing so, I will refer to all sub-steps. I will therefore ask you questions on various topics one by one. These are the preparation, the implementation of the radiotherapy, the support with possible side effects, the visits and the completion of the treatment as well as the aftercare. As already mentioned, we are particularly interested in the topic of patient safety, i.e. any experiences that made you feel safe or unsafe and, above all, what you specifically needed to feel safe. For the interview, it is important that you have the confidence to describe your opinion and experiences openly and honestly. There is no right or wrong. Feel free to share both critical and positive impressions. We are interested in anything that you think is relevant to patient safety. Of course, you do not have to answer a question if you feel uncomfortable with it and you can stop the interview at any time without any disadvantages for you.

**Introductory questions (5 min)**

- ➢ For the context and for my own understanding, I would like to ask a few brief questions about your treatment. As a reminder, we will later anonymize the information about your personal treatment so that no conclusions can be drawn about you personally.
- ➢ What kind of treatment did you receive? (e.g. external radiation, brachytherapy or a combination)
- ➢ Did the treatment take place on an outpatient or inpatient basis?
- ➢ Which area was irradiated?
- ➢ Was surgery or chemotherapy or other therapy with medication for the same illness carried out before or after?
- ➢ Did you also receive radiotherapy outside this clinic? If yes, where?

**Appendix 4: Patient interview guide**
Article: Development and content validity of the Patient Safety in Radiation Oncology questionnaire (PaSaRO): A multi-method study
Authors: M. Grohmann, E. Christalle, F. Schwenzer, M. Jäckel, N. Michalowski, I. Scholl, A. Baehr

- Approximately how long did the entire radiotherapy last?
- How long ago did you receive the treatment?

**Introduction of the topic of patient safety (5 min)**

- I would like to move straight into the topic of patient safety.
- What does safety mean to you as a patient in the context of radiotherapy? What do you understand by the term?
- I will now go through various stages of radiotherapy with you step by step. As I said, I would like to focus on the topic of patient safety. That's why there are three key questions that I will explicitly ask again and again and that you are welcome to keep in mind as a thematic framework.
- When you think about your own treatment, what were the experiences that made you feel safe?
- What were experiences that made you feel unsafe?
- What should the healthcare professionals ideally have done to make you feel safe?

**Pretreatment consultation (10 min)**

- Think back to your own treatment, starting with your very first appointment at the radiotherapy clinic/practice. Please think back to your first consultation at the clinic/radiotherapy practice.
- What was this conversation about?
- Do you remember who you spoke to?
- Were you alone in the conversation or did you have someone with you?
- We return to the topic of patient safety. What topics did you find particularly relevant to your own safety during the first appointment?
- Was there anything in the conversation that caused you to have concerns about your safety? If so, what exactly was it? Why did it make you feel unsafe?
- How should the conversation ideally have gone for you to feel particularly safe?

- What questions were you asked?
  - Which questions did you feel were particularly relevant to your safety?
  - Which questions should ideally have been asked to make you feel safe?

- What information was given to you?
  - How was the information given to you? (e.g. verbally, in writing)
  - What is important for you so that you can understand the information well?
  - What is important for you to be able to remember the information well?
  - Was there any information that you were missing at this point?

- What information did you receive about side effects?
  - What information would you have liked to receive about side effects?
  - What information did you receive about late effects?
  - What information would you have liked to receive on late effects?

**Appendix 4: Patient interview guide**
Article: Development and content validity of the Patient Safety in Radiation Oncology questionnaire (PaSaRO): A multi-method study
Authors: M. Grohmann, E. Christalle, F. Schwenzer, M. Jäckel, N. Michalowski, I. Scholl, A. Baehr

- Topic: Further preparations:
  - ➢ Apart from this consultation, what else was done before your first radiotherapy treatment?
  - ➢ What needs to happen before the first radiotherapy so that you feel that the treatment is safe?
  - ➢ Have you had any markings placed?
    - What information did you receive about the markings?
    - To what extent was it explained to you when and how they would be applied?
    - What was explained to you about the care of the markings?
    - To what extent was it explained to you how the completeness and condition of the markings are checked?
    - What information would you have liked to receive on the subject of markings?
  - ➢ Were you given radiation protection plasters?
    - What was explained to you about the cases in which they are applied?
    - To what extent was it explained to you when they are applied and how they are changed?

**Execution of the radiotherapy (10 min)**

- ➢ Now think back to the radiotherapy itself. So now it's all about what happened on the days you came for radiotherapy.
- ➢ Were there any questions that you were asked every day? [If yes: Which ones? (if necessary, add: this may have been done informally, for example "How are you?")
- ➢ Which ones do you think should have been asked to make you feel safe?
- ➢ What did the healthcare professionals do that made you feel safe?
- ➢ Were there any experiences that made you feel unsafe or worried?

- Topic: "speak up"
  - ➢ Suppose you had noticed something unusual, e.g. something during the preparation that was done differently than usual: How could you have reacted?
    Who could you have contacted?
  - ➢ Was it explained to you who you could turn to if something seemed strange or if you felt unsafe?
  - ➢ Have you experienced such a situation in which you noticed something strange or felt unsafe?
    - What exactly happened?
    - Did you tell the healthcare professionals?
    - How did they react?
    - How did you feel about this reaction?
    - How should the healthcare professionals have reacted to something like this?

- Topic: Preconditions for radiotherapy
  - ➢ Did you have any instructions on how you had to appear for the radiotherapy? (e.g. bladder filling) If yes:
    - How was this checked?
    - How easy was it for you to comply?
    - What was explained to you about why the instructions were given?
    - How were you supported in meeting the requirements?
    - What would you have needed to be able to fulfill the requirements well?

**Appendix 4: Patient interview guide**

Article: Development and content validity of the Patient Safety in Radiation Oncology questionnaire (PaSaRO): A multi-method study

Authors: M. Grohmann, E. Christalle, F. Schwenzer, M. Jäckel, N. Michalowski, I. Scholl, A. Baehr

- Topic: Interfaces
- ➢ Different, often changing healthcare professionals are involved in radiation treatment. How did you
  experience the exchange of information between the healthcare professionals?
  - To what extent did you have the impression that all the healthcare professionals knew a lot about you and your illness?
  - To what extent did you experience situations in which you yourself were responsible for passing on important information?

**Visits/ doctor's consultations (10 min)**

- ➢ Do you remember the visits/doctor's consultations?
  - How often were there doctor's consultations?
  - Can you describe how a typical doctor's consultations went?
- ➢ To what extent were you informed that you could have a visit or a doctor's appointment at any time?
  - Did you actively ask for a consultation? If so, what was the reason for this?
- ➢ What should ideally have happened during the visits or doctor's consultations to make you feel safe?
- ➢ What do you think was particularly relevant for your safety during the visits?

**Supportive therapy (10 min)**

(Treatment of symptoms caused by tumor or radiation, e.g. nausea)

- ➢ Side effects can occur during radiotherapy, such as damage to the skin or teeth, nausea or tiredness. Have you experienced any of these?
  - To what extent have such side effects already been treated as a precaution?
  - How did the healthcare professionals react to this?
  - How did the healthcare professionals determine/investigate/ask about such side effects?
  - In your opinion, what should the healthcare professionals do to ensure that such side effects are
  - recognized?
  - How were the side effects treated?
  - What support would you have liked in dealing with side effects?
- ➢ To what extent were you emotionally burdened by the radiotherapy?
  - How did the healthcare professionals deal with it?
  - To what extent did the healthcare professionals pay attention to/ask about such stress?
  - What support was offered to you?
  - What reaction would you have liked from the healthcare professionals?
  - What support would you have liked to receive?

**Completion of radiotherapy/follow-up (10 min)**

- ➢ Here I am interested in what information was given to you about follow-up.
  - How was the information given to you? (e.g. verbally, in writing)
  - What is important for you so that you can understand the information well?
  - What is important for you to be able to remember the information well?
  - Was there any information that you were missing at this point?
- ➢ What documents were given to you?
  - What documents do you think would have been important for your safety?
- ➢ What support would you have liked to have had after completing treatment?
  - What information did you receive about social and legal support, e.g. for rehabilitation
  - applications, care services, etc.?
  - How important was this for your safety?

**Appendix 4: Patient interview guide**

Article: Development and content validity of the Patient Safety in Radiation Oncology questionnaire (PaSaRO): A multi-method study

Authors: M. Grohmann, E. Christalle, F. Schwenzer, M. Jäckel, N. Michalowski, I. Scholl, A. Baehr

- How well did you know what to do after completing treatment?
  - How well were you informed about further appointments, e.g. when and where your next appointments would take place?
  - To what extent were you supported in arranging follow-up appointments?

**Conclusion of the interview**

- Are there any other topics that are important to you for patient safety that I did not ask about?
- Do you want to add something?

- Thank you.
