## Appendix 5 Cognitive interview guide for "Development and content validity of the Patient Safety in Radiation Oncology questionnaire (PaSaRO): A multi-method study"

Article: Development and content validity of the Patient Safety in Radiation Oncology questionnaire (PaSaRO): A multi-method study

Authors: M. Grohmann, E. Christalle, F. Schwenzer, M. Jäckel, N. Michalowski, I. Scholl, A. Baehr

Interview guide for cognitive interviews with experts

**Purpose:**

The aim of the cognitive interviews is to test the comprehensibility of the patient safety indicators (PSI) developed for the Patient Safety in Radiation Oncology questionnaire (PaSaRO).

The interviews include all patient safety indicators that were selected in the relevance rating of a two-stage Delphi process. Participants will see different PSIs in their interview. On the one hand, some items are only presented to certain professional groups who can evaluate the PSI appropriately in terms of content. Secondly, the duration of the cognitive interviews is limited to one hour. Participants are radiation oncology experts from five professional groups: physicians, medical physicists, nurses, administrative staff and radiation therapists. The PSIs are asked in a fixed order until the maximum duration (1 hour) is reached.

The interviews will be conducted via the platform Zoom and audio-recorded. The interviewer's attitude is interested, open, neutral and appreciative. The PSIs are read out to the participants by the interviewer. In addition, they can read the items themselves in a PowerPoint presentation shared in Zoom.

**Introduction (5 min)**

- As you have already read, the interview will be recorded. I have an audio recorder here [showing device] which I will use to record the interview. After the interview, we will anonymize the audio recording by overlaying all names, location details, etc. with a noise. The anonymized audio files will be stored on a server at the University Medical Center Hamburg-Eppendorf and will only be accessible to our project staff.
- Do you agree to this? [wait for consent] Then I'll start the recording now.
- We are conducting this interview as part of the PaSaGeRO study. Our aim is to promote patient safety in radiotherapy. To this end, we are developing a questionnaire for healthcare professionals to provide clinics and practices with feedback on the current status of patient safety and possible improvement measures. The questionnaire will ask for various statements on patient safety. Topics are, for example,, existing measures, protocols, limit values and technical equipment in the context of radiotherapy. These statements were developed on the basis of scientific literature and expert knowledge. In the end, this questionnaire will be used by a team of several people from different professional groups from one and the same department, who will jointly assess the level of patient safety in their own department. With your help, we would now like to check whether the statements in the questionnaire are easy to understand. We do not want to talk to you about how these topics are handled in your institution, but are only interested in how you understand the statements.
- To do this, I would like to ask you to go through the statements in this questionnaire with me piece by piece and think out loud. Tell me everything you notice, how you understand the statement and anything else that occurs to you.
- I will help you to think aloud by asking a few questions such as “What do you understand by this statement?”, “If you were evaluating this statement for your department, what would you think about?” or “What reasons can you think of why you would have answered the statement this way?”.

- At some points, I will also ask you to rephrase individual words or I will ask you questions about them. I will often ask you to describe in your own words what you have just read. This is very unusual at first, but it is important for me to be able to understand how the statements are understood by you. You are welcome to say everything that is on your mind out loud. There is no right or wrong, only your own thoughts.
- If you wish, you can stop the interview at any time without any disadvantages.
  Do you have any questions?

**Interview (up to max. 1 hour total duration)**

**Evaluation of the choices "I cannot assess" and "not relevant for my department"**

- Here you can see 2 sample questions from the questionnaire. As you can see, the statements can be rated on a scale. In this scale there is also the option to select "I cannot assess" and "not relevant for my department".
- Could you give me examples or scenarios where someone who ends up using this questionnaire would select one of the latter choices?

Example items for this slide (plus response scale):

- "For paediatric tumours, radiation treatment plans are reviewed by a referral centre."
- "For the administration of systemic tumour therapies, standards for handling extravasates are defined and appropriate antidotes are available."

**Evaluation of the PSIs**
This is now the first statement of the questionnaire. We will now go through the questionnaire together, statement by statement.

The interviewer shows slides with all available PSIs one after the other according to their main and subcategories. She/he reads out the PSI and supports the participant with one of the following prompts:

**Prompts for the interviews using rephrasing:**

- What do you understand by the statement?
- How do you understand the statement?
- What does the word XXX mean to you?
- Please explain in your own words what you understand by XXX.
- When you try to evaluate this statement for your department, what do you think about?
- What would need to be done for you to check "Completely true"?
- What are examples of how this statement can be implemented?
- For the keyword "standard" - Feel free to ask: "What exactly could such a standard look like?" or "What exactly should be regulated in it?"

**Prompts for the interviews using think aloud:**

- Please think out loud about how you would evaluate this statement for your department?
- Why did you choose this answer?
- What would need to happen in your department for you to be able to select "Fully applies" here?
