## Appendix 6 Sample Characteristics for "Development and content validity of the Patient Safety in Radiation Oncology questionnaire (PaSaRO): A multi-method study"

**Appendix 6: Sample Characteristics**
Article: Development and content validity of the Patient Safety in Radiation Oncology questionnaire (PaSaRO): A multi-method study
Authors: M. Grohmann, E. Christalle, F. Schwenzer, M. Jäckel, N. Michalowski, I. Scholl, A. Baehr

### Sample Characteristics Focus Groups (n=14)

| *Variable* | | *Count* | *Percent* |
|---|---|---|---|
| *Gender* | Male | 10 | 71% |
| | Female | 4 | 29% |
| *Age* | 18–29 years | 1 | 7% |
| | 30–39 years | 8 | 57% |
| | 40–49 years | 2 | 14% |
| | 50–59 years | 2 | 14% |
| | 60–69 years | 1 | 7% |
| *Profession* | Physicians | 4 | 29% |
| | Radiation Therapists | 4 | 29% |
| | Medical Physics Experts | 3 | 21% |
| | Nurses | 3 | 21% |
| | Administration | 0 | 0% |
| *Professional Experi-ence* | < 5 years | 1 | 7% |
| | 5–10 years | 4 | 29% |
| | 11–20 years | 7 | 50% |
| | > 20 years | 2 | 14% |
| *Current Employer* | University hospital or maximum care hospital | 13 | 93% |
| | Primary/standard care hospital | 0 | 0% |
| | Practice or MVZ (Medical Care Center) | 1 | 7% |
| *Previous Employer* | University hospital or maximum care hospital | 6 | 43% |
| | Primary/standard care hospital | 1 | 7% |
| | Practice or MVZ (Medical Care Center) | 5 | 36% |
| | No response | 4 | 29% |
| *Techniques - Currently working with* | Teletherapy | 11 | 79% |
| | Brachytherapy | 9 | 64% |
| | Adaptive | 1 | 7% |
| | SRS (stereotactic radiosurgery) | 11 | 79% |
| | SBRT (stereotactic body radiotherapy) | 11 | 79% |
| | IMRT (intensity modulated radiotherapy) | 11 | 79% |
| | SGRT (surface-guided radiotherapy) | 2 | 14% |
| | Systemic antitumor substances (e.g. chemotherapy) | 5 | 36% |
| *Techniques – Previ-ous Experience* | Teletherapy | 10 | 71% |
| | Brachytherapy | 9 | 64% |
| | Adaptive | 1 | 7% |
| | SRS (stereotactic radiosurgery) | 10 | 71% |
| | SBRT (stereotactic body radiotherapy) | 10 | 71% |
| | IMRT (intensity modulated radiotherapy) | 10 | 71% |
| | SGRT (surface-guided radiotherapy) | 3 | 21% |
| | Systemic antitumor substances (e.g. chemotherapy) | 5 | 36% |

**Appendix 6: Sample Characteristics**

Article: Development and content validity of the Patient Safety in Radiation Oncology questionnaire (PaSaRO): A multi-method study

Authors: M. Grohmann, E. Christalle, F. Schwenzer, M. Jäckel, N. Michalowski, I. Scholl, A. Baehr

### Sample Characteristics Patient Interviews (n=10)

| *Variable* | | *Count* | *Percent* |
|---|---|---|---|
| *Gender* | Male | 5 | 50% |
| | Female | 5 | 50% |
| *Age* | 18–29 years | 0 | 0% |
| | 30–39 years | 1 | 10% |
| | 40–49 years | 2 | 20% |
| | 50–59 years | 1 | 10% |
| | 60–69 years | 4 | 40% |
| | ≥70 years | 2 | 20% |
| *Tumour entitiy* | Breast | 2 | 20% |
| | Prostate | 1 | 10% |
| | Sarcoma | 1 | 10% |
| | Colorectal tumor | 1 | 10% |
| | Lymphoma | 1 | 10% |
| | Breast with brain metastases | 2 | 20% |
| | Skin tumor with brain metastases | 1 | 10% |
| | Head and neck tumor | 1 | 10% |
| *Metastatic Disease* | Yes | 3 | 30% |
| | No | 7 | 70% |
| *Surgery* | Yes | 7 | 70% |
| | No | 3 | 30% |
| *Chemotherapy* | Yes | 4 | 40% |
| | No | 6 | 60% |
| *Other Medical Therapy* | Yes | 4 | 40% |
| | No | 6 | 60% |

**Appendix 6: Sample Characteristics**
Article: Development and content validity of the Patient Safety in Radiation Oncology questionnaire (PaSaRO): A multi-method study
Authors: M. Grohmann, E. Christalle, F. Schwenzer, M. Jäckel, N. Michalowski, I. Scholl, A. Baehr

Sample Characteristics Cognitive Interviews (n=16)

| *Variable* | | *Count* | *Percent* |
|---|---|---|---|
| *Gender* | Male | 8 | 50% |
| | Female | 8 | 50% |
| *Age* | 18–29 years | 3 | 19% |
| | 30–39 years | 7 | 44% |
| | 40–49 years | 1 | 6% |
| | 50–59 years | 3 | 19% |
| | 60–69 years | 1 | 6% |
| | No response | 1 | 6% |
| *Profession* | Physicians | 7 | 44% |
| | Radiation Therapists | 1 | 6% |
| | Medical Physics Experts | 6 | 38% |
| | Nurses | 1 | 6% |
| | Administration | 0 | 0% |
| | No response | 1 | 6% |
| *Professional Experience* | < 5 years | 7 | 44% |
| | 5–10 years | 2 | 13% |
| | 11–20 years | 3 | 19% |
| | > 20 years | 3 | 19% |
| | No response | 1 | 6% |
| *Current Employer* | University hospital or maximum care hospital | 14 | 88% |
| | Primary/standard care hospital | 0 | 0% |
| | Practice or MVZ (Medical Care Center) | 0 | 0% |
| | Other | 1 | 6% |
| | No response | 1 | 6% |
| *Previous Employer* | University hospital or maximum care hospital | 3 | 19% |
| | Primary/standard care hospital | 0 | 0% |
| | Practice or MVZ (Medical Care Center) | 8 | 50% |
| | No response | 6 | 38% |
| *Techniques - Currently working with* | Teletherapy | 13 | 81% |
| | Brachytherapy | 11 | 69% |
| | Adaptive | 1 | 6% |
| | SRS (stereotactic radiosurgery) | 13 | 81% |
| | SBRT (stereotactic body radiotherapy) | 13 | 81% |
| | IMRT (intensity modulated radiotherapy) | 13 | 81% |
| | SGRT (surface-guided radiotherapy) | 6 | 38% |
| | Systemic antitumor substances (e.g. chemotherapy) | 9 | 56% |
| | No response | 1 | 6% |
| *Techniques - Previous Experience* | Teletherapy | 12 | 75% |
| | Brachytherapy | 10 | 63% |
| | Adaptive | 1 | 6% |
| | SRS (stereotactic radiosurgery) | 11 | 69% |
| | SBRT (stereotactic body radiotherapy) | 11 | 69% |
| | IMRT (intensity modulated radiotherapy) | 11 | 69% |
| | SGRT (surface-guided radiotherapy) | 6 | 38% |
| | Systemic antitumor substances (e.g. chemotherapy) | 7 | 44% |
| | No response | 1 | 6% |

**Appendix 6: Sample Characteristics**
Article: Development and content validity of the Patient Safety in Radiation Oncology questionnaire (PaSaRO): A multi-method study
Authors: M. Grohmann, E. Christalle, F. Schwenzer, M. Jäckel, N. Michalowski, I. Scholl, A. Baehr

### Sample Characteristics Delphi Study (Round 1)

| *Variable* | | *Count* | *Percent* |
|---|---|---|---|
| *Gender* | Male | 27 | 38% |
| | Female | 43 | 61% |
| *Age* | 18–29 years | 4 | 6% |
| | 30–39 years | 33 | 48% |
| | 40–49 years | 16 | 23% |
| | 50–59 years | 10 | 15% |
| | 60–69 years | 6 | 9% |
| *Profession* | Physicians | 26 | 31% |
| | Radiation Therapists | 25 | 30% |
| | Medical Physics Experts | 20 | 24% |
| | Nurses | 5 | 6% |
| | Administration | 10 | 12% |
| *Professional Experience* | < 5 years | 4 | 6% |
| | 5–10 years | 23 | 32% |
| | 11–20 years | 23 | 32% |
| | > 20 years | 21 | 30% |
| *Current Employer* | University hospital or maximum care hospital | 46 | 69% |
| | Primary/standard care hospital | 4 | 6% |
| | Practice or MVZ (Medical Care Center) | 17 | 25% |
| *Previous Employer* | University hospital or maximum care hospital | 43 | 51% |
| | Primary/standard care hospital | 14 | 17% |
| | Practice or MVZ (Medical Care Center) | 28 | 33% |
| | No response | 14 | 17% |
| *Techniques - Currently working with* | Teletherapy | 53 | 63% |
| | Brachytherapy | 38 | 45% |
| | Adaptive | 15 | 18% |
| | SRS (stereotactic radiosurgery) | 45 | 54% |
| | SBRT (stereotactic body radiotherapy) | 54 | 64% |
| | IMRT (intensity modulated radiotherapy) | 65 | 77% |
| | SGRT (surface-guided radiotherapy) | 32 | 38% |
| | Systemic antitumor substances (e.g. chemotherapy) | 28 | 33% |
| | No response | 4 | 5% |
| *Techniques - Previous Experience* | Teletherapy | 52 | 62% |
| | Brachytherapy | 48 | 57% |
| | Adaptive | 11 | 13% |
| | SRS (stereotactic radiosurgery) | 47 | 56% |
| | SBRT (stereotactic body radiotherapy) | 53 | 63% |
| | IMRT (intensity modulated radiotherapy) | 63 | 75% |
| | SGRT (surface-guided radiotherapy) | 30 | 36% |
| | Systemic antitumor substances (e.g. chemotherapy) | 29 | 35% |

**Appendix 6: Sample Characteristics**

Article: Development and content validity of the Patient Safety in Radiation Oncology questionnaire (PaSaRO): A multi-method study

Authors: M. Grohmann, E. Christalle, F. Schwenzer, M. Jäckel, N. Michalowski, I. Scholl, A. Baehr

Sample Characteristics Delphi Study (Round 2)

| *Variable* | | *Count* | *Percent* |
|---|---|---|---|
| *Profession* | Physicians | 23 | 32% |
| | Radiation Therapists | 20 | 28% |
| | Medical Physics Experts | 19 | 26% |
| | Nurses | 4 | 6% |
| | Administration | 6 | 8% |
