## Appendix 7 Full list of PSIs for "Development and content validity of the Patient Safety in Radiation Oncology questionnaire (PaSaRO): A multi-method study"

**Appendix 7: Full list of patient safety indicators with Delphi ratings and source**

Article: Development and content validity of the Patient Safety in Radiation Oncology questionnaire (PaSaRO): A multi-method study

Authors: Maximilian Grohmann, Eva Christalle, Felicitas Schwenzer, Maria Jäckel, Nina Michalowski, Isabelle Scholl, Andrea Baehr

This supplement provides the complete list of 158 Patient Safety Indicators (PSIs) identified through our systematic literature review. The indicators are organized into main categories and subcategories, with each PSI presented in its original German formulation alongside the English translation. Translations were performed using DeepL, a service developed and operated by DeepL SE (Cologne). Each PSI is linked to its corresponding source reference, including DOI or ISBN where available.

| Category | Sub-Category | Patient Safety Indicator (German) | Patient Safety Indicator | Change in wording after round 1 | Acceptance round 1 | Acceptance round 2 | Implementation feasibility rating (mean) | Ease of data collection rating (mean) | Reference | Excerpt |
|---|---|---|---|---|---|---|---|---|---|---|
| 1. Human resources | Promotion of continuous training and skills assessment | Es wird sichergestellt, dass Mitarbeitende über aktuelle Forschungsergebnisse informiert sind (z.B. durch regelmäßige Journal Clubs, Fortbildungen, Kongressteilnahmen o.ä.). | It is ensured that employees are informed about current research findings (e.g. through regular journal clubs, training courses, participation in conferences, etc.). | no | 85,70% | 91,80% | 2,8 | 3,1 | American Society for Radiation Oncology, 2019, Safety is no accident A framework for quality radiation oncology care | Each member of the team should participate in available continuing medical education. |
| | | | | | | | | | British Institute of Radiology et al., 2008, Towards Safer Radiotherapy | Following initial attainment of competence, all professional staff need to maintain their skills by lifelong learning through continuing professional development (CPD), which is a requirement for the maintenance of registration in most regulatory frameworks. |
| | | | | | | | | | Nishioka et al., 2022, Phys. Imaging Radiat. Oncol. 23: 1–7, doi:10.1016/j.phro.2022.06.002 | The third most common corrective measure was education. |
| | | | | | | | | | British Institute of Radiology et al., 2008, Towards Safer Radiotherapy | Training records should be created and maintained for all staff involved in radiotherapy. They should be detailed and specific to particular procedures. Funding to support training should be available. |
| | | | | | | | | | Pardo Masferrer et al., 2020, Clin. Transl. Oncol. Off. Publ. Fed. Span. Oncol. Soc. Natl. Cancer Inst. Mex. 22: 751–758, doi:10.1007/s12094-020-02359-x | What is clear is that, due to the continuous technological and methodological renovation of any specialty in general and Radiation Oncology in particular, continuing education is vital to ensure the incorporation of new knowledge into clinical practice. |
| | | | | | | | | | Focus group | I would have thought it was standard. That journal clubs are really possible. That the SOPs would be presented regularly with PowerPoint slides and the like. I'm appalled that it's not standard. |
| | | | | | | | | | Focus group | I once experienced this at the bowel center, where at least two nurses simply had to undergo further specialist training. And of course, once they had completed this, the quality of care also increased because they then provided advice, gave instructions and of course also had to deal with studies. |
| 1. Human resources | Promotion of continuous training and skills assessment | Alle Mitarbeitenden, die an der Applikation systemischer Tumortherapien beteiligt sind, nehmen dazu an einem kontinuierlichen Fortbildungsprogramm teil. | All employees involved in the application of systemic tumor therapies take part in a continuous training program. | yes | 90,90% | 93,00% | 2,6 | 3,3 | Neuss et al., 2017, Oncol. Nurs. Forum 44: 31–43, doi:10.1188/17.ONF.31-43 | The health care setting uses a comprehensive education program for initial and ongoing educational requirements for all staff who prepare and administer chemotherapy. |
| | | | | | | | | | Greenall et al., 2015, J. Oncol. Pharm. Pract. Off. Publ. Int. Soc. Oncol. Pharm. Pract. 21: 26–35, doi:10.1177/1078155214556522 | [...] staff, who are involved in any aspect of the management of chemotherapy and biotherapy, from prescribing to disposal, should participate in a standardized orientation that includes a competency evaluation. |
| 1. Human resources | Promotion of continuous training and skills assessment | Eine gleichbleibend hohe Konturierungsqualität wird sichergestellt (z.B. durch regelmäßige Vorstellung von aktuellen Konturierungsrichtlinien oder durch peer-review basierte Besprechungen bei problematischen Fällen). | Consistently high contouring quality is ensured (e.g. through regular presentation of current contouring guidelines or through peer-review based discussions in problematic cases). | no | 90,20% | 90,20% | 2,8 | 2,8 | Tsang et al., 2019, Tech. Innov. Patient Support Radiat. Oncol. 10: 8–12, doi:10.1016/j.tipsro.2019.05.001 | Our study has demonstrated the statistically significant variation that exists in trial protocol compliances of TV and OAR contouring during the pre-trial QA period for two UK lung cancer radiotherapy trials and therefore highlights the importance of performing outlining QA for the purposed of clinician feedback to help minimize contouring variation. |
| | | | | | | | | | Schimek-Jasch et al., 2015, Strahlenther. Onkol. Organ Dtsch. Rontgengesellschaft Al 191: 525–533, doi:10.1007/s00066-015-0812-8 | Teaching after DR1 resulted in a reduction of absolute TVs in DR2, as well as in better concordance of TVs. |
| | | | | | | | | | Yamazaki et al., 2011, Radiat. Oncol. Lond. Engl. 6: 10, doi:10.1186/1748-717X-6-10 | To obtain reproducible outcomes using an inverse plan, consensus among the participants should be reached in advance to avoid uncertainty; for example, definitions of major violations should be provided and training sessions made available for participants to improve the conformity of their plans to an agreed upon benchmark. |
| | | | | | | | | | Tsang et al., 2019, Tech. Innov. Patient Support Radiat. Oncol. 10: 8–12, doi:10.1016/j.tipsro.2019.05.001 | There were statistically significant differences in trial protocol compliances between clinical oncologists' target volume and organs at risk contours. |

#### Appendix 7: Full list of patient safety indicators with Delphi ratings and source

Article: Development and content validity of the Patient Safety in Radiation Oncology questionnaire (PaSaRO): A multi-method study

Authors: Maximilian Grohmann, Eva Christalle, Felicitas Schwenzer, Maria Jäckel, Nina Michalowski, Isabelle Scholl, Andrea Baehr

| Category | Sub-Category | Patient Safety Indicator (German) | Patient Safety Indicator | Change in wording after round 1 | Acceptance round 1 | Acceptance round 2 | Implementation feasibility rating (mean) | Ease of data collection rating (mean) | Reference | Excerpt |
|---|---|---|---|---|---|---|---|---|---|---|
| 1. Human resources | Promotion of continuous training and skills assessment | Das Personal wird bei Einstellung auf Grundlage der für sie relevanten Standardverfahren (SOPs) geschult. | Staff are trained on the basis of the relevant standard operating procedures (SOPs) when they are hired. | no | 90,10% | 90,10% | 3,3 | 3,3 | American Society for Radiation Oncology, 2019, Safety is no accident A framework for quality radiation oncology care | Documentation of SOPs is also critical to train new staff. Both training and documentation should be updated often. In particular, it is often necessary to retrain staff after time away from a system, or to refresh current knowledge. |
| | | | | | | | | | Focus group | Further training where you don't just always do the latest things, but also present and illustrate what the standard is here for certain things, [...]. New staff come in, something changes again. And to keep everyone on the same level in the first place, so that they are valuable to the department and also feel like a valuable member. |
| 1. Human resources | Promotion of continuous training and skills assessment | Das ärztliche Personal verfügt über ausreichende Kenntnisse über Anwendung, Wirkungen und Nebenwirkungen von systemischen tumorwirksamen Therapien. | The medical staff have sufficient knowledge of the use, effects and side effects of systemic tumor therapies. | no | 98,10% | 98,10% | 3,1 | 2,7 | Focus group | One is the topic of drug-based tumor therapy, which fortunately is also required of specialists, […] I think you have to have given these chemos once and also the immunotherapies […]. |
| 1. Human resources | Staffing | Der aktuelle Personalschlüssel spiegelt die tatsächliche Abteilungs-Auslastung angemessen wieder. Dabei werden zusätzliche Aufgaben des Teams wie Administration, Forschung oder Lehre angemessen berücksichtigt. | The current staffing ratio appropriately reflects the actual departmental workload. Additional tasks of the team such as administration, research or teaching are taken into account appropriately. | no | 87,80% | 90,30% | 2 | 2,9 | Smith et al., 2014, J. Oncol. Pract. 10: 350–357, doi:10.1200/JOP.2013.001353 | […] half of respondents (53.9% of physicians and 43.7% of nonphysicians; […]) responded that overburdened workloads were a source of errors. |
| | | | | | | | | | Focus group | But that's also the case in nursing, isn't it? I mean, there's a delirious patient in one room where three nursing staff are actually working on it. But it's one out of ten, […]. So of course no distinction is made here. |
| | | | | | | | | | Focus group | How many staff do I really need to be able to work safely and to a high standard? Somehow the radiation protection legislation has some kind of vague limits that are defined, no, "in sufficient numbers". What is a sufficient number? And I think we need to reach an agreement on this so that it doesn't lead to institutions being bled dry and then at some point really only being able to work with a minimum number of staff. |
| | | | | | | | | | British Institute of Radiology et al., 2008, Towards Safer Radiotherapy | Other departmental duties, such as teaching, research and development, should be taken into account when establishing appropriate staffing levels. |
| | | | | | | | | | Focus group | There are usually one or two people who only look after one medical physicist in training. And although this means you have another member of staff in the building, you have another experienced member of staff who is basically blocked off to train the new person. |
| 1. Human resources | Staffing | Es wird regelmäßig überprüft, ob der Personalschlüssel angemessen ist. | Regular checks are carried out to ensure that the staffing ratio is appropriate. | no | 88,70% | 91,80% | 2,7 | 2,9 | British Institute of Radiology et al., 2008, Towards Safer Radiotherapy | To ensure that the safe delivery of radiotherapy is maintained, each centre should formally review its skills mix and staffing levels at intervals of no more than two years and ensure these comply with national guidance. Additional reviews should be carried out during the planning of new treatment techniques or procedures and before they are introduced. |
| 1. Human resources | Staffing | Für die Einarbeitung neuer Mitarbeitender gibt es ein strukturiertes Konzept, das bei allen Mitarbeitenden umgesetzt und dokumentiert wird. | There is a structured concept for the induction of new employees, which is implemented and documented for all employees. | yes | 89,30% | 95,20% | 2,9 | 3,1 | Focus group | And then, of course, somehow you need an induction concept, [...]. The question is, is it just something that exists on paper, or is it something that is also worked through as a curriculum. You could at least keep track of the fact that people have all found out how things work during onboarding. |
| 1. Human resources | Staffing | Die Qualifikationen und Einteilung des vorhandenen Personals im Dienstplan ermöglichen eine gleichbleibende Behandlungsqualität. | The qualifications and allocation of the existing staff in the duty roster enable a consistent quality of treatment. | no | 93,20% | 93,20% | 2,6 | 3 | Focus group | [...] we won't be able to guarantee that a nurse knows the patient from start to finish. But it's important to me that it's one of the team. […] We are now so mixed that I am grateful if there is even one regular on the shift. |
| | | | | | | | | | Focus group | And I also think what you said is an equally important point, that you deploy people where they have their qualities. So as the boss, you actually know your people and you know that one of them is fitter in this area, he'll tear up mountains, but if you put him in the other position, then he's such a stopper and won't achieve anything. |
| 1. Human resources | Staffing | Auf Station ist ein Betreuungsschlüssel definiert. Bei Unterschreitung werden Betten geschlossen. | A nurses-patients-ratio is defined for each ward. Beds are closed if this ratio is not met. | no | 89,90% | 95,20% | 2,2 | 3,1 | Focus group | So there is a limit. How much can a nurse do, so to speak, how many patients can you care for [...] they say quite clearly that today there are only two nurses, one nurse can only care for ten patients. [...] And then the beds are closed and patients are not admitted. |
| 1. Human resources | Staffing | Die Mitarbeitenden mit direktem Patient:innenkontakt verfügen über ausreichende Deutschkenntnisse, um die Kommunikation mit den Patient:innen zu gewährleisten und sicherzustellen, dass keine relevanten Informationen verloren gehen. | Employees with direct patient contact have sufficient knowledge of German to ensure communication with patients and to ensure that no relevant information is lost. | no | 98,60% | 98,60% | 3 | 3,1 | Focus group | So we have, [...] a Greek woman who has been here for three years in May. And you still can't put her in the first position on her own, which for us would be the planning CT, for example, because she simply doesn't understand the patients properly and they don't understand her either. |
| | | | | | | | | | Focus group | On the negative side, I have also seen a lack of language skills, because this can often lead to misunderstandings on the part of both patients and practitioners of all kinds, which in turn can lead to mistakes. |

### Appendix 7: Full list of patient safety indicators with Delphi ratings and source

Article: Development and content validity of the Patient Safety in Radiation Oncology questionnaire (PaSaRO): A multi-method study

Authors: Maximilian Grohmann, Eva Christalle, Felicitas Schwenzer, Maria Jäckel, Nina Michalowski, Isabelle Scholl, Andrea Baehr

| Category | Sub-Category | Patient Safety Indicator (German) | Patient Safety Indicator | Change in wording after round 1 | Acceptance round 1 | Acceptance round 2 | Implementation feasibility rating (mean) | Ease of data collection rating (mean) | Reference | Excerpt |
|---|---|---|---|---|---|---|---|---|---|---|
| 1. Human resources | Staffing | Ärztliches Personal, welches in der Bestrahlungsplanung arbeitet, ist in der radiologischen Bildbetrachtung und -befundung adäquat geschult. | Medical staff working in radiation planning are adequately trained in radiological image viewing and diagnosis. | yes | 96,70% | 94,10% | 2,5 | 2,7 | Focus group | Everyone should have had contact with radiodiagnostics. In other words, you should have assessed images before you start planning your radiation treatment. CTs and MRIs in particular. |
| 1. Human resources | Staffing | Für jedes Bestrahlungsgerät gibt es ein:e MT-R oder ein Team von MT-Rs, die fest dort eingeteilt sind und somit Gerät und Patient:innen gut kennen. | There is one RTT or a team of RTTs for each radiation device, who are permanently assigned there and therefore know the device and patients well. | no | 95,10% | 95,10% | 2,7 | 3,2 | Patient interview | Then you are called by name again and then I was sometimes addressed by name in the treatment room. Especially by those I had already seen a few times. So I always knew that they knew my name and who I was. |
| | | | | | | | | | Focus group | From the RTT`s point of view, we try to set things up in such a way that there is always an RTT at the unit who knows the patient. [...] it's always good if someone at least knows what happened last week and perhaps even knows the patient from the start. |
| | | | | | | | | | Focus group | So we have a team leader for each of the devices and a certain core team. And the rest rotate, of course. And then it's always important to have someone who always feels responsible. |
| | | | | | | | | | Patient interview | Yes, it was often the case that one person changed and I already knew one of them and so on. That was rare, (...) maybe not at all, that there were two complete strangers here. Somehow it was always a smooth transition. |
| 1. Human resources | Training for new technologies and processes | Standardvorgehen (SOPs) für die Ausbildung von Mitarbeitenden sind festgelegt, dokumentiert und für alle zugänglich. | Standard operating procedures (SOPs) for employee training are defined, documented and accessible to all. | no | 94,50% | 94,50% | 3,3 | 3,4 | American Society for Radiation Oncology, 2019, Safety is no accident A framework for quality radiation oncology care | Practice policies must exist for appropriate training and competency assessment of personnel. Each practice must ensure that clinical staff are able to maintain continued competence in their job responsibilities. |
| 1. Human resources | Training for new technologies and processes | Vor der Einführung neuer Verfahren werden alle Nutzer:innen unter Berücksichtigung möglicher Fehler- und Gefahrenquellen adäquat geschult. | Before new procedures are introduced, all users are adequately trained, taking into account potential sources of error and danger. | no | 94,40% | 94,40% | 2,9 | 3,1 | Kisling et al., 2019, Med. Phys. 46: 2567–2574, doi:10.1002/mp.13552 | Training should educate the end users of automated planning systems about the potential failure modes, the impact of these failures on patients, and the need for careful manual review of the plans to prevent these failures. |
| | | | | | | | | | Focus group | So if it's a new linear accelerator [...] which is the basic device for our radiation, then I think it's good if as many people as possible know about it. If it's an additional device that you use from time to time, then one or two people are enough. |
| | | | | | | | | | Focus group | But once you say, here are ten minutes, by the way, I've learned this, now you can do it. That's a completely different relationship to the one person who may have received three days of training. |
| 1. Human resources | Training for new technologies and processes | Alle Mitarbeitenden, die stereotaktische Bestrahlungen planen oder durchführen, sind speziell geschult worden. | All employees who plan or carry out stereotactic radiotherapy have been specially trained. | no | 87,90% | 94,70% | 2,9 | 3,2 | Sahgal et al., 2020, Pract. Radiat. Oncol. 10: 243–254, doi:10.1016/j.prro.2019.11.002 | Radiation Oncologist: Participation in a dedicated fellowship or course that provides technologyspecific training is strongly recommended |
| 1. Human resources | Training for new technologies and processes | Bei der Einführung neuer Techniken werden Mitarbeitende in externen Kursen geschult und schulen weitere Mitarbeitende in internen Fortbildungen. | When new technologies are introduced, employees are trained in external courses and train other employees in internal training courses. | yes | 88,20% | 95,20% | 2,8 | 3,1 | Sahgal et al., 2020, Pract. Radiat. Oncol. 10: 243–254, doi:10.1016/j.prro.2019.11.002 | Participation in a dedicated fellowship or course that provides technologyspecific training is strongly recommended |
| | | | | | | | | | Focus group | Then there are usually two or three people who are trained by the company and the rest can simply learn from them. |
| | | | | | | | | | Focus group | So I really don't think it makes sense to be there with ten or 15 people. Because then the two or three people who could learn well instead might also- Well, if you're in a small room with a lot of people, there's always a lot of chatting around and less concentration. |
| | | | | | | | | | Focus group | But once you say, here are ten minutes, by the way, I've learned this, now you can do it. That's a completely different relationship to the one person who may have received three days of training. |
| 1. Human resources | Training for new technologies and processes | Für die Einführung neuer Techniken arbeiten Mitarbeitende mehrerer Berufsgruppen in einem Projektteam zusammen. | | no | 82,40% | 90,00% | 2,5 | 3 | British Institute of Radiology et al., 2008, Towards Safer Radiotherapy | Each radiotherapy centre should hold regular multidisciplinary management meetings. In addition, there should be regular multidisciplinary meetings to discuss operational issues, including the introduction of new technologies and practices. These meetings should be informal to encourage interprofessional challenge, while respecting professional boundaries and qualifications. |
| 2. institutional culture | Promotion of a safety culture | Die Teilnahme aller Berufsgruppen an Versammlungen und Arbeitsgruppen, wie z.B. Morbidity and Mortality (M&M) -Konferenzen und Gruppen zur Einführung neuer Prozesse, wird durch die Führungsebene gewünscht, gefördert und ermöglicht. | The participation of all professional groups in meetings and working groups, such as morbidity and mortality (M&M) conferences and groups for the introduction of new processes, is desired, encouraged and facilitated by the management. | no | 90,40% | 90,40% | 2,7 | 3 | Focus group | [...] on this day, on this date, this conference is taking place. Why don't you come along? [...] there are emails that are sent around, but that's where it starts, that many people just aren't there. And another obstacle is, depending on the scheduling, of course, that's working time, many people don't have the motivation, maybe not after the early shift, to attend this conference [...] |

#### Appendix 7: Full list of patient safety indicators with Delphi ratings and source

Article: Development and content validity of the Patient Safety in Radiation Oncology questionnaire (PaSaRO): A multi-method study

Authors: Maximilian Grohmann, Eva Christalle, Felicitas Schwenzer, Maria Jäckel, Nina Michalowski, Isabelle Scholl, Andrea Baehr

| Category | Sub-Category | Patient Safety Indicator (German) | Patient Safety Indicator | Change in wording after round 1 | Acceptance round 1 | Acceptance round 2 | Implementation feasibility rating (mean) | Ease of data collection rating (mean) | Reference | Excerpt |
|---|---|---|---|---|---|---|---|---|---|---|
| 2. insti-tutional culture | Promotion of a safety culture | Es herrscht eine Sicherheitskultur, in der alle Mitarbeitenden unabhängig von der Hierarchie unerwünschte Ereignisse und Beinahe-Ereignisse melden können, ohne Angst vor negativen Konsequenzen zu haben. Ein offener und respektvoller Umgang wird gefördert und es wird akzeptiert, dass solche Ereignisse vorkommen. | There is a safety culture in which all employees, regardless of hierarchy, can report undesirable events and near misses without fear of negative consequences. Open and respectful interaction is encouraged and it is accepted that such events occur. | no | 93,20% | 93,20% | 2,7 | 2,3 | Ford et al., 2018, Med. Phys. 45: 100–119, doi:10.1002/mp.12800 | [...] several RT studies have explicitly noted the importance of just culture |
| | | | | | | | | | Pardo Masferrer et al., 2020, Clin. Transl. Oncol. Off. Publ. Fed. Span. Oncol. Soc. Natl. Cancer Inst. Mex. 22: 751–758, doi:10.1007/s12094-020-02359-x | The Heads of Department should encourage an open, cooperative, non-hierarchical, non-punitive environment of mutual respect and participation, and are responsible for creating a safety culture where all staff members are encouraged to report errors without fear to the punitive consequences. |
| | | | | | | | | | Smith et al., 2014, J. Oncol. Pract. 10: 350–357, doi:10.1200/JOP.2013.001353 | Three other concerns were statistically significantly more important to physicians compared with nonphysicians: getting colleagues into trouble (47.7% of physicians and 31.1% of nonphysicians), admitting liability (41.5% of physicians and 24.6% of nonphysicians), and effect on departmental reputation (46.2% of physicians and 27.6% of nonphysicians). |
| | | | | | | | | | British Institute of Radiology et al., 2008, Towards Safer Radiotherapy | The ability of staff to talk to their colleagues and superiors about safety incidents is an important feature of creating a culture which is open and fair, and which is non-punitive. |
| | | | | | | | | | Vijayakumar et al., 2019, Front. Oncol. 9: 302, doi:10.3389/fonc.2019.00302 | Anybody can Raise His/Her Hand |
| | | | | | | | | | Khader et al., 2019, Radiat. Oncol. J. 37: 60–65, doi:10.3857/roj.2019.00080 | It is unrealistic to expect zero errors or modifications in the clinical work of a busy radiotherapy department. |
| | | | | | | | | | Focus group | [...] but you always have to say that we all make mistakes, we're all human and it's not BAD. And I think that's very, very important and that you also do a bit of psychology and say, don't stress yourself, it can happen to anyone, [...] But I think it's really important that you know that - that you won't be punished for it or anything. |
| | | | | | | | | | Focus group | As a manager, you always have the intention of having a positive error culture anyway. And then I've always been told that it's really important for them that you're always their point of contact, that they can always come to you. That you don't have anything to fear. |
| | | | | | | | | | Focus group | I mean, I think it's the same here - everyone knows that mistakes can happen. And you don't really get your head ripped off. [...] accusations are made very quickly or prematurely, where you then run up and say, you did something wrong and how could that be? And that's why I don't think we're that willing to do so. |
| 2. insti-tutional culture | Promotion of a safety culture | Bei der Meldung, Aufarbeitung und Besprechung von Fehlern wird sich nicht auf meldende oder verursachende Personen konzentriert, sondern auf die Prozesse, die ursächlich oder begünstigend waren. | When reporting, processing and discussing errors, the focus is not on the persons reporting or causing them, but on the processes that caused or facilitated them. | no | 95,90% | 95,90% | 2,9 | 2,6 | Focus group | Actually, the purpose of this lecture, at least that's how we do it, is to learn something from it. So we don't say, haha, look how stupid he is, he's mixed up right and left, but we've found that we have patients with half a leg and a whole leg. And the half leg was to be irradiated, i.e. the stump was to be irradiated, and then the healthy leg was irradiated. |
| | | | | | | | | | Focus group | And then I was always told that it's really important for them that you're always their point of contact, that they can always come to you. That you don't have anything to fear. |
| 2. insti-tutional culture | Promotion of a safety culture | Die Führungsebene ermutigt alle Mitarbeitenden zum Berichten von unerwünschten Ereignissen und Beinahe-Ereignissen. | Management encourages all employees to report adverse events and near misses. | no | 97,30% | 97,30% | 3,2 | 2,6 | Ford et al., 2018, Med. Phys. 45: 100–119, doi:10.1002/mp.12800 | Engage departmental leadership. The need to engage in incident learning is compelling. It is supported by data and is recommended by professional societies and virtually every relevant national and international body. |
| 2. insti-tutional culture | Promotion of a safety culture | Durch die Führungsebene wird sichergestellt, dass jede:r Mitarbeitende aktiv bei der Verbesserung klinischer Prozesse mitwirken kann (z.B. durch regelmäßige Feedbackrunden). | The management level ensures that every employee can actively participate in the improvement of clinical processes (e.g. through regular feedback rounds). | no | 93,10% | 93,10% | 2,8 | 2,5 | American Society for Radiation Oncology, 2019, Safety is no accident A framework for quality radiation oncology care | […] must empower all staff to actively participate in improving clinical processes without fear of reprimand or reprisal. |
| 2. insti-tutional culture | Promotion of a safety culture | Die Kommunikation der Führungspersonen der einzelnen Berufsgruppen untereinander und gegenüber dem Personal sind förderlich für die Patient:innensicherheit. | Communication between the managers of the individual professional groups and with staff is conducive to patient safety. | no | 95,80% | 95,80% | 3 | 2,6 | Focus group | […] then the bosses […] they play each other off against each other, who has more power than the other. And that's (...), I think, what makes it really difficult. Although of course it would be very, very, very desirable for the patients. |

**Appendix 7: Full list of patient safety indicators with Delphi ratings and source**
Article: Development and content validity of the Patient Safety in Radiation Oncology questionnaire (PaSaRO): A multi-method study
Authors: Maximilian Grohmann, Eva Christalle, Felicitas Schwenzer, Maria Jäckel, Nina Michalowski, Isabelle Scholl, Andrea Baehr

| Category | Sub-Category | Patient Safety Indicator (German) | Patient Safety Indicator | Change in wording after round 1 | Acceptance round 1 | Acceptance round 2 | Implementation feasibility rating (mean) | Ease of data collection rating (mean) | Reference | Excerpt |
|---|---|---|---|---|---|---|---|---|---|---|
| 2. insti-tutional culture | Promotion of a safety culture | Die Kommunikation im Team ist förderlich für die Patient:innensicherheit. | Communication within the team is conducive to patient safety. | no | 97,30% | 97,30% | 3 | 2,6 | Focus group | I spontaneously wrote down that I see a lack of or poor communication, i.e. information flow, and then the documentation of this information as really dangerous. |
| | | | | | | | | | Focus group | However, communication between professional groups could definitely be improved, because there is a risk that important things simply arrive too late at the bottom of the accelerator. Yes, that's why I think communication within the professional group is good, but with other professional groups it could be improved or is a risk. |
| | | | | | | | | | Focus group | It's more about making sure that the atmosphere is good, that everyone gets along, that nobody argues. Because the moment you have these disruptive factors under control to some extent, the information transfer suddenly works. Most mistakes happen when everyone's attention is focused on arguing. That's my experience on the subject. So maintaining a good atmosphere in the team is pretty essential. |
| 2. insti-tutional culture | Promotion of a safety culture | Die Führungsebene fordert und fördert das Prinzip von Peer Review in der klinischen Arbeit. | The management level demands and promotes the principle of peer review in clinical work. | no | 94,50% | 94,50% | 3,1 | 2,8 | Marks et al., 2013, Pract. Radiat. Oncol. 3: 149–156, doi:10.1016/j.prro.2012.11.010 | Leadership need to empower the staff to be involved in peer review activities |
| 2. insti-tutional culture | Internal feedback loops | Unerwünschte Ereignisse und Beinahe-Ereignisse werden regelmäßig in neutraler Umgebung besprochen, um daraus zu lernen. | Adverse events and near-misses are regularly discussed in a neutral environment in order to learn from them. | no | 95,80% | 95,80% | 3 | 3,1 | Smith et al., 2014, J. Oncol. Pract. 10: 350–357, doi:10.1200/JOP.2013.001353 | If errors and near-miss events are presented on a regular basis in a clear and nonthreatening environment where the goal is to learn from each other's mistakes, professionals may feel less intimidated by admitting their own mistakes. |
| | | | | | | | | | Wright et al., 2021, Pract. Radiat. Oncol. 11: 92–100, doi:10.1016/j.prro.2020.05.002 | Another key role of the SAQ program is to promote education about safety and quality outcomes, most commonly through "morbidity and mortality" conferences or "locoregional failure rounds," for this review, |
| | | | | | | | | | Focus group | Yes, but it's actually desirable to go one step further and talk about it openly. It's anonymous, so no name or anything is written in. [...]We're not perfect, for God's sake, that's the way it is, right? |
| | | | | | | | | | Focus group | So if I have now irradiated the patient incorrectly due to a mistake from an incorrect plan, I would like to know, yes. |
| 2. insti-tutional culture | Internal feedback loops | Die Ergebnisse von Risikoanalysen werden allen Mitarbeitenden mitgeteilt, die im bewerteten Prozess tätig sind. | The results of risk analyses are communicated to all employees involved in the assessed process. | yes | 94,40% | 91,90% | 3,1 | 3,2 | Malicki et al., 2018, Radiother. Oncol. J. Eur. Soc. Ther. Radiol. Oncol. 127: 164–170, doi:10.1016/j.radonc.2018.04.006 | The minimum provisions necessary for such a programme include: (1) the designation of a management team to allocate dedicated resources and provide risk management training; (2) an organizational and/or departmental culture of quality management and safety; (3) a risk management committee; (4) a risk manager and multidisciplinary team within the radiation oncology department; (5) dissemination of results. |
| 2. insti-tutional culture | Internal feedback loops | Das Personal wird kontinuierlich über laufende und abgeschlossene Verbesserungsmaßnahmen informiert, insbesondere als Reaktion auf gemeldete unerwünschte Ereignisse. | Staff are continuously informed about ongoing and completed improvement measures, particularly in response to reported adverse events. | no | 95,90% | 95,90% | 3,2 | 3,2 | American Society for Radiation Oncology, 2019, Safety is no accident A framework for quality radiation oncology care | The committee must regularly educate all staff about ongoing quality initiatives and continuously perform quality improvement. |
| | | | | | | | | | Focus group | And then this is ultimately reported back to the team so that these mistakes are no longer made in the next step. This is then discussed in full with the team. |
| 2. insti-tutional culture | Internal feedback loops | Es finden regelhaft und mindestens einmal im Quartal Morbidity and Mortality (M&M)-Konferenzen statt, bei denen unerwünschte Ereignisse, Dosisabweichungen, unerwartete Verläufe, Nebenwirkungen und Therapieabbrüche diskutiert werden. | Morbidity and Mortality (M&M) conferences are held regularly and at least once a quarter, at which adverse events, dose deviations, unexpected courses, side effects and treatment discontinuations are discussed. | yes | 94,20% | 94,20% | 3 | 3,3 | American Society for Radiation Oncology, 2019, Safety is no accident A framework for quality radiation oncology care | Practices must at a minimum hold rounds quarterly, or more typically monthly, to review patient morbidity and mortality, dose discrepancies and any incident reports involving an accident, injury or untoward effect to a patient. Morbidity and mortality reviews should include unusual or severe acute complications of treatment, unexpected deaths or unplanned treatment interruptions. |
| | | | | | | | | | Focus group | We always have training sessions twice a year on which errors occurred during the year. And then the ten most interesting cases are usually presented completely anonymously and explained, what happened, why it happened, what we have done to prevent it from happening again and always with nice pictures. And anyone who wants to can come to these training sessions. |

#### Appendix 7: Full list of patient safety indicators with Delphi ratings and source

Article: Development and content validity of the Patient Safety in Radiation Oncology questionnaire (PaSaRO): A multi-method study

Authors: Maximilian Grohmann, Eva Christalle, Felicitas Schwenzer, Maria Jäckel, Nina Michalowski, Isabelle Scholl, Andrea Baehr

| Category | Sub-Category | Patient Safety Indicator (German) | Patient Safety Indicator | Change in wording after round 1 | Acceptance round 1 | Acceptance round 2 | Implementation feasibility rating (mean) | Ease of data collection rating (mean) | Reference | Excerpt |
|---|---|---|---|---|---|---|---|---|---|---|
| 2. insti-tutional culture | Internal feedback loops | Die in Nachsorgen erhobenen Nebenwirkungen der Behandlungen werden systematisch erfasst und gegen Vergleichsdaten (z.B. aus Studien) abgeglichen. | The side effects of the treatments recorded in follow-up care are systematically recorded and compared with comparative data (e.g. from studies). | no | 83,60% | 92,30% | 2,6 | 2,9 | Wright et al., 2021, Pract. Radiat. Oncol. 11: 92–100, doi:10.1016/j.prro.2020.05.002 | Departments should periodically review clinical outcomes of patients, which may include review of both oncologic outcomes and treatment toxicities. This applies to both academically focused and clinically focused departments. |
| | | | | | | | | | Vaandering et al., 2023, Radiother. Oncol. J. Eur. Soc. Ther. Radiol. Oncol. 178: 109433, doi:10.1016/j.radonc.2022.11.022 | [QI] Reported maximum acute radiodermatitis grading for breast cancer patients treated with EBRT only |
| | | | | | | | | | American Society for Radiation Oncology, 2019, Safety is no accident A framework for quality radiation oncology care | Changes in patient response to treatment may identify large or even subtle changes in technique, equipment performance or clinical decision strategies, and are a valuable independent check on the success of the practice's overall QM system |
| 2. insti-tutional culture | Involvement of the patients | Patient:innen werden ermutigt anzusprechen, wenn ihnen etwas Ungewöhnliches auffällt oder sie Sicherheitsbedenken haben. | Patients are encouraged to speak up if they notice anything unusual or have safety concerns. | no | 85,70% | 91,80% | 3,3 | 2,5 | Patient interview | Yes, they definitely said that if I had any problems, I should just say something. Not specifically, but simply in the room, so to speak, and it was also the case that if something seemed strange to me, I already had the feeling at the end of the radiation that the implant was causing or would cause a problem, then a doctor immediately looked over it. |
| | | | | | | | | | Patient interview | […] if I notice that something is not as it usually is or I'm irritated, then I usually bring it up because it's very personal what's happening. |
| | | | | | | | | | Bibault et al., 2016, Cancer Radiother. J. Soc. Francaise Radiother. Oncol. 20: 790–793, doi:10.1016/j.canrad.2016.06.006 | When such events were perceived, 61% of the patients (n = 14) talked about it with their radiation therapists. When they did not talk about it, it was because they did not deem it as significant (36%, n = 4) or because they were afraid they would seem like they were questioning their radiation therapists competencies (27%, n = 3). |
| | | | | | | | | | Sundararaman et al., 2014, Pract. Radiat. Oncol. 4: 181–188, doi:10.1016/j.prro.2013.09.003 | […] empower our patients to speak up in the cause of safety. We continually reinforce their need to speak up about anything that seems unsafe or contrary to their expectations. |
| 2. insti-tutional culture | Involvement of the patients | Patient:innen werden ermutigt, Nebenwirkungen während der Therapie auch ungefragt anzusprechen. | Patients are encouraged to address side effects during treatment, even without being asked. | no | 96,10% | 96,10% | 3,5 | 2,7 | Patient interview | Patient: [...] So I described my side effects and then I was offered the doctor's consultation. Interviewer: Okay. To what extent were you informed that you could have a visit at any time? Patient: Yes, I was informed. That's why I asked. |
| 2. insti-tutional culture | Involvement of the patients | Fragen und Hinweisen der Patient:innen zur Sicherheit werden ernst genommen und ihnen wird nachgegangen. | Patients' questions and comments on safety are taken seriously and followed up. | no | 98,70% | 98,70% | 3,3 | 2,6 | Patient interview | […] and actually my name was always there, for my control, I could check it myself, but once I actually had the wrong name, which I pointed out and they changed it again […] |
| | | | | | | | | | Patient interview | Of course, you first want to be taken seriously with your concerns, so that you're not told, "Oh, don't worry, because then you'll worry all the more," but that it's explained to you briefly, so to speak, that these are the reasons why event XY has just happened |
| | | | | | | | | | Patient interview | Interviewer: So how did you experience the reactions to this message from the people treating you? Patient: Very good. They thought it was good that I got in touch, that I said something, and then they did everything they could in concrete terms. |
| | | | | | | | | | British Institute of Radiology et al., 2008, Towards Safer Radiotherapy | Concerns raised by patients must be taken seriously and investigated promptly. |
| 2. insti-tutional culture | Involvement of the patients | Patient:innen werden ermutigt, bei der täglichen Identifikation vor Therapie aktiv mitzuwirken (z.B. auf die korrekte Nennung ihres Namens achten). | Patients are encouraged to actively participate in the daily identification before therapy (e.g. make sure they give their name correctly). | new | | 93,40% | 3,4 | 2,8 | Free text Delphi | |

**Appendix 7: Full list of patient safety indicators with Delphi ratings and source**
Article: Development and content validity of the Patient Safety in Radiation Oncology questionnaire (PaSaRO): A multi-method study
Authors: Maximilian Grohmann, Eva Christalle, Felicitas Schwenzer, Maria Jäckel, Nina Michalowski, Isabelle Scholl, Andrea Baehr

| Category | Sub-Category | Patient Safety Indicator (German) | Patient Safety Indicator | Change in wording after round 1 | Acceptance round 1 | Acceptance round 2 | Implementation feasibility rating (mean) | Ease of data collection rating (mean) | Reference | Excerpt |
|---|---|---|---|---|---|---|---|---|---|---|
| 3. quality and risk manageme nt | Existence and effectiveness of a system for reporting and analyzing errors | Es existiert ein abteilungsinternes Meldesystem, welches alle sicherheitsrelevante Vorfälle (inklusive unerwünschte Ereignisse und Beinahe-Ereignisse) erfasst, auch wenn sie keine unmittelbaren Auswirkungen auf die Patient:innen hatten. | There is an internal departmental reporting system that records all safety-related incidents (including adverse events and near misses), even if they had no direct impact on patients. | no | 87,50% | 96,60% | 2,8 | 3,1 | Ford et al., 2018, Med. Phys. 45: 100–119, doi:10.1002/mp.12800 | It is, therefore, important to expand the scope of ILS beyond incidents that reach the patient to include near-miss events and other safety-related reports. |
| | | | | | | | | | Focus group | Yes, exactly, what I meant was that it is more low-threshold, that there is simply, well, I don't know, either an e-mail box that is also written in or the classic way, that you write something in by hand, it doesn't matter which is easier for you [...]. |
| | | | | | | | | | Focus group | We now have a half self-written reporting system for radiotherapy only, i.e. without a ward, but only for RTTs, doctors and physicists. And we have three categories. Errors, near misses, these are the more critical categories, but we also have the inconvenience category. And in the inconvenience category, there are points that you can simply click on, such as bladder not filled, culture not on time, so really trivial things. But there is also the option for free text. |
| | | | | | | | | | Focus group | I think somehow it would be better for the department - or my feeling would be better - if there were more subliminal reports, so that you could simply [...] have something like a mailbox, [...]where you can handwrite what you have just noticed [...]. |
| 3. quality and risk manageme nt | Existence and effectiveness of a system for reporting and analyzing errors | In der Einrichtung ist ein niederschwelliges Fehlermeldesystem implementiert (z.B. leicht verwendbar). | A low-threshold error reporting system is implemented in the facility (e.g. easy to use). | no | 88,40% | 96,80% | 2,8 | 3 | Focus group | I think somehow it would be better for the department - or my feeling would be better - if there were more subliminal reports, so that you could simply [...] have something like a mailbox, [...]where you can handwrite what you have just noticed [...]. |
| | | | | | | | | | Focus group | I don't know whether it's helpful to have a completely - it's always - low-threshold reporting tool or something so that people can enter everything so that they don't approach anyone. We don't have that. |
| 3. quality and risk manageme nt | Existence and effectiveness of a system for reporting and analyzing errors | Nach unerwünschten Ereignissen erfolgt eine Aufarbeitung nach definiertem Vorgehen, wie z.B. einer Root-Cause-Analyse oder Fehlerbaumanalyse. | After adverse events, a defined procedure is followed, such as a root cause analysis or fault tree analysis. | no | 95,70% | 95,70% | 3 | 3,2 | Ford et al., 2018, Med. Phys. 45: 100–119, doi:10.1002/mp.12800 | Major events must be handled expediently and thoroughly and may call for a full root-cause analysis. |
| | | | | | | | | | British Institute of Radiology et al., 2008, Towards Safer Radiotherapy | Following a level 1 or 2 radiation incident, a systematic investigation should be conducted to identify the root causes. To prevent recurrence, the lessons learnt from root cause analysis should be disseminated. |
| 3. quality and risk manageme nt | Existence and effectiveness of a system for reporting and analyzing errors | Es existiert eine standardisierte Prozesskette, falls unerwünschte Ereignisse eine unmittelbare Auswirkung auf den tagesaktuellen Ablauf haben. | A standardized process chain is in place in the event that adverse events have an immediate impact on daily operations. | yes | 86,60% | 96,50% | 2,8 | 3 | Focus group | So, the route is actually first to me or to my deputy and then to the doctor. And then he discusses it with the physics department at lunchtime to see to what extent we need to do things or not. If it's something more serious, if the wrong side is irradiated or something like that, it's also possible that the boss will talk to the patient directly. |
| 3. quality and risk manageme nt | Existence and effectiveness of a system for reporting and analyzing errors | Die gemeldeten sicherheitsrelevanten Vorfälle (inklusive unerwünschte Ereignisse und Beinahe-Ereignisse) aus dem Meldesystem werden für die Erarbeitung von Verbesserungsmaßnahmen genutzt. | The reported safety-related incidents (including adverse events and near misses) from the reporting system are used to develop improvement measures. | no | 95,80% | 95,80% | 3 | 3 | American Society for Radiation Oncology, 2019, Safety is no accident A framework for quality radiation oncology care | Each practice should have a review committee […] This committee organizes the collection and analysis of safety events via an incident learning system in a non-punitive environment, works to identify potential problems in devices or processes, and then tries to mitigate these problems by modifying processes or adding new checks or actions to minimize the likelihood of further problems. |
| | | | | | | | | | Focus group | [...] As a result, [...] in order to track it better in the long term, we will always link this Vision-RT, the surface system with the Linac, only when it is green and releases can we irradiate. So, we have simply drawn a conclusion on how to make irradiation safer. |

**Appendix 7: Full list of patient safety indicators with Delphi ratings and source**
Article: Development and content validity of the Patient Safety in Radiation Oncology questionnaire (PaSaRO): A multi-method study
Authors: Maximilian Grohmann, Eva Christalle, Felicitas Schwenzer, Maria Jäckel, Nina Michalowski, Isabelle Scholl, Andrea Baehr

| Category | Sub-Category | Patient Safety Indicator (German) | Patient Safety Indicator | Change in wording after round 1 | Acceptance round 1 | Acceptance round 2 | Implementation feasibility rating (mean) | Ease of data collection rating (mean) | Reference | Excerpt |
|---|---|---|---|---|---|---|---|---|---|---|
| 3. quality and risk management | Existence and effectiveness of a system for reporting and analyzing errors | Es gibt ein interprofessionelles Risikomanagement-Team, das unerwünschte Ereignisse und Beinahe-Ereignisse bearbeitet und daraus Strategien zur Prozessoptimierung ableitet. | There is an interprofessional risk management team that deals with adverse events and near misses and derives strategies for process optimization. | no | 88,60% | 93,10% | 2,7 | 3,2 | American Society for Radiation Oncology, 2019, Safety is no accident A framework for quality radiation oncology care | Each practice should have a review committee which monitors quality issues, near-misses and errors in treatment, diagnosis, patient care or other procedural problems that might lead to errors. This committee organizes the collection and analysis of safety events via an incident learning system in a non-punitive environment, works to identify potential problems in devices or processes, and then tries to mitigate these problems by modifying processes or adding new checks or actions to minimize the likelihood of further problems. |
| | | | | | | | | | Malicki et al., 2018, Radiother. Oncol. J. Eur. Soc. Ther. Radiol. Oncol. 127: 164–170, doi:10.1016/j.radonc.2018.04.006 | The minimum provisions necessary for such a programme include: (1) the designation of a management team to allocate dedicated resources and provide risk management training; (2) an organizational and/or departmental culture of quality management and safety; (3) a risk management committee; (4) a risk manager and multidisciplinary team within the radiation oncology department; (5) dissemination of results. |
| | | | | | | | | | Sanders et al., 2020, Brachytherapy 19: 762–766, doi:10.1016/j.brachy.2020.08.014 | Review of brachytherapy incidents by teams of at least physicians, physicists, and therapists on a regular basis is highly encouraged to create feedback loops to improve quality and safety in care. |
| | | | | | | | | | American Society for Radiation Oncology, 2019, Safety is no accident A framework for quality radiation oncology care | A dedicated formal QA committee should consist of an interdisciplinary clinical team (e.g., physicians, physicists, dosimetrists, nurses, radiation therapists) and other staff (administrative and IT support) that incorporates all disciplines involved in each treatment modality, meets regularly. |
| | | | | | | | | | Khader et al., 2019, Radiat. Oncol. J. 37: 60–65, doi:10.3857/roj.2019.00080 | Another initiative at our department aiming to decrease major medications was the quality improvement committee which started by the end of 2015 responsible for standardizing radiotherapy dose and volume for each site and implementing a library for the terms used in target and OAR delineation. |
| | | | | | | | | | Ford et al., 2014, Med. Phys. 41:, doi:10.1118/1.4875687 | Second, a core group of leaders or “champions” was identified. This core group consisted of seven people selected to represent all the various professional groups within the department. |
| | | | | | | | | | Focus group | And then we also have this QM meeting once a month, where we discuss it internally with the management. And then this is ultimately reported back to the team so that we don't make these mistakes again in the next step, so to speak. This is then discussed in full with the team. And then at the next QM meeting, it's discussed again to see if we've really got it right. Have these errors occurred again? |
| 3. quality and risk management | Existence and effectiveness of a system for reporting and analyzing errors | Dem Risikomanagement-Team werden ausreichend Zeit und Ressourcen zugeteilt. | Sufficient time and resources are allocated to the risk management team. | no | 89,70% | 93,10% | 2,1 | 2,7 | American Society for Radiation Oncology, 2019, Safety is no accident A framework for quality radiation oncology care | Adequate time and resources should be allocated for the QM program, including regular peer review to monitor adherence. |
| 3. quality and risk management | Existence and effectiveness of a system for reporting and analyzing errors | Abteilungsinterne Prozesse werden auch unabhängig von gemeldeten unerwünschten Ereignissen von einem interprofessionellen Team evaluiert. | Internal departmental processes are also evaluated by an interprofessional team independently of reported adverse events. | no | 78,80% | 92,90% | 2,4 | 2,9 | British Institute of Radiology et al., 2008, Towards Safer Radiotherapy | Each radiotherapy centre should hold regular multidisciplinary management meetings. In addition, there should be regular multidisciplinary meetings to discuss operational issues, including the introduction of new technologies and practices. These meetings should be informal to encourage interprofessional challenge, while respecting professional boundaries and qualifications. |
| 3. quality and risk management | External peer review process | Für kindliche Tumore werden die Bestrahlungspläne durch ein Referenzzentrum überprüft. | The radiation plans for childhood tumors are reviewed by a reference center. | no | 87,80% | 95,90% | 2,6 | 3,1 | Carrie et al., 1999, Int. J. Radiat. Oncol. Biol. Phys. 45: 435–439, doi:10.1016/s0360-3016(99)00200-x | Pretreatment central quality assurance review or standardized computer-designed blocks would improve survival to an extent equivalent to that attributed to adjuvant chemotherapy. |
| | | | | | | | | | Lüders et al., 2014, Eur. J. Cancer Oxf. Engl. 1990 50: 425–433, doi:10.1016/j.ejca.2013.09.017 | Of 142 patients without guidance by the reference centre, 56 (39%) received a treatment deviating from protocol recommendations: chemotherapy doses were either lower or higher than recommended in 17% of patients, and deviations concerning radiotherapy dose and treatment volume occurred in 25% and 20%, respectively. |
| 3. quality and risk management | External peer review process | Bei der Neueinführung von Therapieformen wird die Planqualität mit erfahrenen Kliniken verglichen. | When new forms of therapy are introduced, the plan quality is compared with experienced clinics. | no | 86,70% | 92,60% | 2,4 | 2,9 | Hartgerink et al., 2019, Acta Oncol. Stockh. Swed. 58: 1275–1282, doi:10.1080/0284186X.2019.1633016 | To optimize the treatment planning- and delivery technique of an institution, it is advisable to compare treatment plans with other institutions, preferably institutions that have broad experience with LINAC based SRS. |

**Appendix 7: Full list of patient safety indicators with Delphi ratings and source**
Article: Development and content validity of the Patient Safety in Radiation Oncology questionnaire (PaSaRO): A multi-method study
Authors: Maximilian Grohmann, Eva Christalle, Felicitas Schwenzer, Maria Jäckel, Nina Michalowski, Isabelle Scholl, Andrea Baehr

| Category | Sub-Category | Patient Safety Indicator (German) | Patient Safety Indicator | Change in wording after round 1 | Acceptance round 1 | Acceptance round 2 | Implementation feasibility rating (mean) | Ease of data collection rating (mean) | Reference | Excerpt |
|---|---|---|---|---|---|---|---|---|---|---|
| 3. quality and risk management | Internal peer review process | Jeder Fall und das Behandlungskonzept werden in einer Besprechung mit mehreren fachärztlichen Mitarbeitenden diskutiert. Dabei sind ggf. auch andere Berufsgruppen anwesend. | Each case and the treatment concept are discussed in a meeting with several board certified physicians. Other professional groups may also be present. | no | 89,90% | 98,20% | 3,1 | 3,3 | American Society for Radiation Oncology, 2019, Safety is no accident A framework for quality radiation oncology care | […] details of each patient's evaluation and intent for treatment is briefly presented to the other radiation oncologists and clinical team and is used as early peer review for the basic treatment decisions and plan. |
| 3. quality and risk management | Internal peer review process | Jeder Bestrahlungsplan wird in einer Besprechung mit mehreren Fachkolleg:innen diskutiert. | Each treatment plan is discussed in a meeting with several specialist colleagues. | no | 82,60% | 93,10% | 3,1 | 3,2 | Fogarty et al., 2001, Australas. Radiol. 45: 189–194, doi:10.1046/j.1440-1673.2001.00901.x | The Chart Round performs QA of radiation treatment and delivery in real time and has rapid feedback loops, so that such deficiencies in patient evaluation and treatment can be identified and corrected while these factors still have a bearing on outcome. |
| | | | | | | | | | Khader et al., 2019, Radiat. Oncol. J. 37: 60–65, doi:10.3857/roj.2019.00080 | Out of the 7,149 RT plans, our study showed that on average 7% needed modifications. |
| | | | | | | | | | Brunskill et al., 2017, Int. J. Radiat. Oncol. Biol. Phys. 97: 27–34, doi:10.1016/j.ijrobp.2016.09.015 | It is easy to begin to believe that peer review will result in the most correct plan. Of the 2830 cases presented for peer review, a change was recommended in 346 cases (12.2%) and categorized as a dose change in 28.3%, a target change in 69.1%, and a major treatment change in 2.6%. |
| | | | | | | | | | Focus group | I also consider this to be a very important aspect of the training. It's also because it's an early meeting, doctors and a person from physics. And that's simply how I think you learn as a junior doctor over a long period of time what is being done. So without all that, I think I would have seen far fewer images and far fewer plans in my career. |
| | | | | | | | | | Focus group | [...] if there are several people in charge of the plan, it's more likely to be noticed if something is wrong than if there's only one person working on it. |
| 3. quality and risk management | Internal peer review process | In täglichen Besprechungen von Fall und Behandlungskonzept sind Vertreter:innen der Medizinphysik, Ärzteschaft und MT-Rs anwesend. | Representatives of medical physics, physicians and RTTs are present at daily case and treatment concept meetings. | no | 79,40% | 96,60% | 2,6 | 3,2 | Focus group | Interviewer: Would you see an advantage in [RTTs] having a fixed appearance in the early meeting, where they know that I could ask such a question in a standardized way, […]? Participant: Absolutely. |
| | | | | | | | | | Focus group | The RTTs know from this discussion what is important later. […] In other words, things that are important are already specifically written down or communicated to the patient. |
| | | | | | | | | | Focus group | But I think it's important that it's discussed in an interdisciplinary team, that you know, okay, something complicated is coming up, you should be careful […] |
| 3. quality and risk management | Internal peer review process | Die Besprechung von Fall und Plan mit Fachkolleg:innen findet vor der ersten Bestrahlung statt. | The case and plan are discussed with specialist colleagues before the first radiation treatment. | no | 89,70% | 100,00% | 3,1 | 3,3 | West et al., 2022, J. Med. Imaging Radiat. Oncol. 66: 129–137, doi:10.1111/1754-9485.13346 | The timing of the peer review occurred pre-treatment for 58% (261/452) of cases and 35% (158/452) occurred during treatment in 2018. […] Application of the PRAT has identified radiation treatment plan modifications that would otherwise go undetected and without opportunity to improve the quality of patients' treatment or avoid harm. |
| | | | | | | | | | Vijayakumar et al., 2019, Front. Oncol. 9: 302, doi:10.3389/fonc.2019.00302 | Peer review performed early on during treatment may be acceptable, but for "zero harm," its performance prior to the first treatment is essential. |
| | | | | | | | | | American Society for Radiation Oncology, 2019, Safety is no accident A framework for quality radiation oncology care | Prospective peer review is critical because once treatment has been initiated, the threshold for making a meaningful change is relatively high because of time-consuming replanning and QA requirements. |
| 3. quality and risk management | Internal peer review process | Die Ergebnisse von internen Fall- und Plan-Besprechungen werden dokumentiert. | The results of internal case and plan discussions are documented. | no | 83,80% | 90,00% | 3,1 | 3,4 | Marks et al., 2013, Pract. Radiat. Oncol. 3: 149–156, doi:10.1016/j.prro.2012.11.010 | The specific goals and targets of peer review should be clearly specified, and the results of each peer review effort ideally should be tracked. |
| 3. quality and risk management | Internal peer review process | In peer-review basierten Besprechungen verabschiedete Planänderungen erfordern eine erneute Vorstellung des geänderten Plans. | Plan amendments adopted in peer-review based meetings require a new presentation of the amended plan. | no | 85,50% | 96,60% | 3,1 | 3,1 | Khader et al., 2019, Radiat. Oncol. J. 37: 60–65, doi:10.3857/roj.2019.00080 | Out of 351 radiotherapy plans that needed major modifications only 158 (45%) were represented. |
| 3. quality and risk management | Internal peer review process | Die Konturierung für alle Patient:innen wird mittels Peer Review überprüft. | The contouring for all patients is checked by means of a peer review. | no | 83,80% | 95,00% | 2,9 | 3,2 | Riegel et al., 2022, J. Appl. Clin. Med. Phys. 23: e13640, doi:10.1002/acm2.13640 | Over half of these (14/24, 58.3%) were solely covered by Daily Contouring Rounds and 6 of the top 10 riskiest failure modes could be detected at this step. […] |
| | | | | | | | | | American Society for Radiation Oncology, 2019, Safety is no accident A framework for quality radiation oncology care | After the radiation oncologist defines target volumes and normal tissues, when possible, another physician should review and confirm the contours before treatment planning begins. |
| | | | | | | | | | Marks et al., 2013, Pract. Radiat. Oncol. 3: 149–156, doi:10.1016/j.prro.2012.11.010 | Since there are significant inter-patient variations in the target volumes and since mis-targeting can lead to poor clinical outcomes, this is considered as one of the most critical areas for peer review […] |

**Appendix 7: Full list of patient safety indicators with Delphi ratings and source**
Article: Development and content validity of the Patient Safety in Radiation Oncology questionnaire (PaSaRO): A multi-method study
Authors: Maximilian Grohmann, Eva Christalle, Felicitas Schwenzer, Maria Jäckel, Nina Michalowski, Isabelle Scholl, Andrea Baehr

| Category | Sub-Category | Patient Safety Indicator (German) | Patient Safety Indicator | Change in wording after round 1 | Acceptance round 1 | Acceptance round 2 | Implementation feasibility rating (mean) | Ease of data collection rating (mean) | Reference | Excerpt |
|---|---|---|---|---|---|---|---|---|---|---|
| 3. quality and risk management | Internal peer review process | In Einrichtungen mit mehreren Standorten finden gemeinsame Besprechungen statt (z.B. durch Videokonferenzsysteme). | In facilities with several locations, joint meetings are held (e.g. using video conferencing systems). | new | | 92,00% | 2,9 | 3,3 | Free text Delphi | For patient safety, it is important that companies with multiple locations hold meetings across locations, e.g. using suitable teleconferencing systems. |
| 3. quality and risk management | Internal peer review process | Es existiert ein Standard, wie assistenzärztliche Kolleg:innen durch fachärztliches Personal in der Patient:innenbetreuung angeleitet werden. | There is a standard for how residents are instructed in patient care by senior physicians. | no | 84,50% | 91,70% | 2,7 | 3 | Focus group | Since they are residents who are just in training, [...] have you done a PET-CT, have you done an MR, have you registered a tumor board? Things like that are constantly forgotten. |
| | | | | | | | | | Patient interview | Patient: She then ticked something I said on the computer. And then she probably didn't feel safe and immediately sent for another doctor, the senior doctor. Yes, and I thought that was very good. |
| 3. quality and risk management | Internal peer review process | Interne intrafraktionelle Positionierungsdaten werden verwendet, um image-guidance-radiotherapy (IGRT)-Toleranzen regelmäßig zu reevaluieren. | Internal intrafractional positioning data is used to regularly re-evaluate image-guidance-radiotherapy (IGRT) tolerances. | no | 88,90% | 93,80% | 2,6 | 2,9 | Al-Hallaq et al., 2022, Med. Phys. 49: 82–112, doi:10.1002/mp.15532 | It is recommended that institutions investigate further if this is not attainable and also update their tolerances as necessary on an annual basis based on analysis of setup reproducibility in their clinic. |
| 3. quality and risk management | Control points and checklists | Vor bestimmten kritischen Prozessstellen kommen Checklisten und kurze Besprechungen aller Beteiligten (sogenannte Time-outs) zum Einsatz (z.B. bei erster Bestrahlung). | Checklists and short meetings of all those involved (so-called time-outs) are used before certain critical process points (e.g. during the first irradiation). | no | 81,40% | 91,20% | 3 | 3 | Kalapurakal et al., 2013, Int. J. Radiat. Oncol. Biol. Phys. 86: 241–248, doi:10.1016/j.ijrobp.2013.02.003 | The introduction of checklists and timeouts resulted in the reduction of the total number of errors from 221 errors in 126 patients to 35 errors in 13 patients. […] The most serious errors such as treatment of the wrong patient, wrong site, and wrong dose reduced from 23 errors in 14 patients to none during the past 3 years. |
| | | | | | | | | | Focus group | There's actually a little checklist on there for the doctors, who then really have to tick it off when the new patient is set up. Is it the right patient? Are we on the right side? Have we made the transfer? Is there a portrait photo? |
| | | | | | | | | | Focus group | [...] where you only find out WHEN the CT is already running or the radiation is already running. Damn, he should definitely have this one special treatment first or then he shouldn't have radiation. Then we have to clarify this again […] |
| | | | | | | | | | Focus group | I've also had positive experiences with checklists, in my case it's about metothrexate administration, for example, from chemo administration. The doctors are obliged to work through this checklist, and so are we as nursing staff [...] So you have a timetable, you can't really forget anything and then everything is really guaranteed to go well. |
| 3. quality and risk management | Control points and checklists | Vor der Tele- oder Brachytherapie mit hohen Einzeldosen kommen Checklisten und kurze Besprechungen aller Beteiligten (sogenannte Time-outs) zum Einsatz. | Before teletherapy or brachytherapy with high individual doses, checklists and short meetings of all those involved (so-called time-outs) are used. | no | 89,10% | 94,40% | 3 | 3 | Sanders et al., 2020, Brachytherapy 19: 762–766, doi:10.1016/j.brachy.2020.08.014 | The vast majority of survey respondents, but not all, performed patient verification before brachytherapy, or had a physician or physicist present during the time-out which is recommended to happen before the procedure and/or treatment delivery. |
| | | | | | | | | | Ford et al., 2017, Semin. Radiat. Oncol. 27: 190–196, doi:10.1016/j.semradonc.2017.02.003 | Many of the issues encountered in treatment delivery can be identified with a well-designed time-out that is specific to SBRT. |
| | | | | | | | | | Focus group | Participant 1: Do you do a timeout before starting the procedure? Participant 2: [...], after we used the wrong applicator once. So we noticed it, but since then we've said, okay, we'll do it from the start. We place the applicator and then discuss it again with the physicians and the RTTs or clearly state the applicator again. |
| 3. quality and risk management | Control points and checklists | Vorhandene Checklisten sind verpflichtend und werden zuverlässig genutzt. | Existing checklists are mandatory and are used reliably. | no | 86,80% | 94,80% | 3 | 3,31 | American Society for Radiation Oncology, 2019, Safety is no accident A framework for quality radiation oncology care | Checklists and time-outs are effective especially if they are focused on the task at hand; the user believes in their utility; and the user is forced to use them (e.g., "hard stop"). |
| | | | | | | | | | Focus group | So they really come to the device, they come in, so to speak, the patient lies on the table, the note is on the table and they take it in their hand and have to look at it. |
| | | | | | | | | | Focus group | But checklists like this help enormously to at least maintain a standard of quality. For example, we have a certain procedure for first treatments, how they are prepared by the RTTs and what then has to be done. It's not easy for every colleague to accept this, [...] And in the end, in one out of ten cases perhaps, this point is very important [...]. |
| | | | | | | | | | Focus group | I've also had positive experiences with checklists, in my case it's about metothrexate administration [...]. And only then, when everything has really been ticked off, are we allowed to start. |

**Appendix 7: Full list of patient safety indicators with Delphi ratings and source**

Article: Development and content validity of the Patient Safety in Radiation Oncology questionnaire (PaSaRO): A multi-method study

Authors: Maximilian Grohmann, Eva Christalle, Felicitas Schwenzer, Maria Jäckel, Nina Michalowski, Isabelle Scholl, Andrea Baehr

| Category | Sub-Category | Patient Safety Indicator (German) | Patient Safety Indicator | Change in wording after round 1 | Acceptance round 1 | Acceptance round 2 | Implementation feasibility rating (mean) | Ease of data collection rating (mean) | Reference | Excerpt |
|---|---|---|---|---|---|---|---|---|---|---|
| 3. quality and risk management | Control points and checklists | An kritischen Stellen verhindern technische Lösungen das Fortsetzen des Prozesses, falls die vorigen Sicherheitschecks nicht bearbeitet wurden. | | yes | 90,00% | 92,90% | 2,6 | 3 | Focus group | Yes, even better than you can do it first, is you can do it first. Otherwise it simply won't work. Of course, this is always the most time-consuming to implement, but it is also the most effective. |
| | | | | | | | | | Focus group | [...], what do we want to ensure with the checklist? Point B can only work if point A has been completed. Ultimately, that is the causality we want to establish. |
| | | | | | | | | | Focus group | The best example is when you have the systems to release plans or something. [...] If you have the status of resident, you can play around as much as you want, but you'll never be able to release the plan because you're simply not activated. [...] And such limits, I think, as you say, are actually the most ideal. |
| 3. quality and risk management | Control points and checklists | Die Mitarbeitenden bewerten vorhandene Checklisten im Arbeitsablauf als sinnvoll. | Employees rate existing checklists in the workflow as useful. | no | 85,90% | 96,60% | 2,8 | 2,7 | American Society for Radiation Oncology, 2019, Safety is no accident A framework for quality radiation oncology care | Checklists and time-outs are effective especially if they are focused on the task at hand; the user believes in their utility; and the user is forced to use them (e.g., "hard stop"). |
| | | | | | | | | | Focus group | I can be a complete stranger to the device. I take this list, I know exactly how it works. I also know, because it has to be countersigned, who did it, and I can ask them in case of doubt if anything deviates from the routine. |
| | | | | | | | | | Focus group | In my opinion, nothing works without lists. Especially checklists. I mean, everyone probably has them somehow in their everyday life. But I think they're short, concise, they describe exactly the most important things and you can just go through them point by point. I somehow think that's the be-all and end-all. |
| 3. quality and risk management | Control points and checklists | Es ist festgelegt, welche Person oder Berufsgruppe die Qualitätskontrollen/Checks im Behandlungspfad durchführt und dokumentiert. | It is specified which person or professional group carries out and documents the quality controls/checks in the treatment pathway. | yes | 90,90% | 90,90% | 3,3 | 3,3 | British Institute of Radiology et al., 2008, Towards Safer Radiotherapy | Each radiotherapy centre should have protocols within its quality system which define what data are to be checked by planners and prescribers along the radiotherapy pathway and how the results of these checks are to be recorded. |
| 3. quality and risk management | Control points and checklists | Die Identität der Patient:innen wird mittels festgelegtem Standard mit zwei unabhängigen Methoden überprüft (z.B. Abfrage des Namens und Abgleich mit einem Foto). | The identity of the patient is checked using a defined standard with two independent methods (e.g. name query and comparison with a photo). | no | 94,60% | 94,60% | 3,5 | 3,2 | Focus group | Nevertheless, if someone refuses a photo, which is still possible with us, they actually have to show their ID every day. I think that makes us quite good. Nonetheless, I think everyone knows that some patients change as a result of the therapy and then it just doesn't really fit. Then we call them by name. They may not hear well or the coffee machine may drown out everything in our waiting area. You still have to pay close attention to this. |
| | | | | | | | | | Focus group | We now also have this surface system, for example. And this is actually used to preset all patients. This means that if the surface, i.e. that of the patient we want to irradiate, does not match the patient, you can see that something has gone wrong. |
| | | | | | | | | | Focus group | [...] we make sure that we really do take photos of the patients, as long as they consent of course and we always have them on our monitor and check to see if it's the right patient. Or, in the case of first radiation sessions, we actually ask them for their first name and date of birth and if they refuse the photo, we always have to ask in between whether it really is them. |
| 3. quality and risk management | Control points and checklists | Die zur Verfügung stehenden Plan-Verifikation-Methoden und Akzeptanz-Kriterien werden auf Wirksamkeit (Sensitivität und Spezifität) überprüft. | The available plan verification methods and acceptance criteria are checked for effectiveness (sensitivity and specificity). | no | 92,60% | 92,60% | 2,8 | 2,8 | McKenzie et al., 2014, Med. Phys. 41: 121702, doi:10.1118/1.4899177 | The same cutoff criteria do not yield the same classification abilities across all devices. Also, this work has shown that QA systems have different abilities to accurately sort acceptable and unacceptable plans. |
| 3. quality and risk management | Control points and checklists | Am Bestrahlungsgerät herrscht eine ruhige Arbeitsatmosphäre, die konzentriertes Arbeiten fördert und Ablenkungen vermeidet. | A quiet working atmosphere prevails at the irradiation unit, which promotes concentrated work and avoids distractions. | no | 88,70% | 98,30% | 2,6 | 2,8 | Focus group | [...] And then there are the two or three people who like to fiddle around on their cell phones. And then one of them comes and says, here, have another look at the plans for right now. Then one of them wants to order another transport and suddenly there's chaos and then Meier is put on instead of Müller. |
| 3. quality and risk management | Failure of therapy-critical processes | Das Personal ist hinsichtlich möglicher Szenarien, die zu einem Ausfall therapiekritischer Prozesse führen könnten (z.B. Stromausfall, Starkwetterereignis, Cyberangriff) durch regelmäßige Information und Notfallübungen vorbereitet. | Staff are prepared for possible scenarios that could lead to a failure of treatment-critical processes (e.g. power failure, severe weather event, cyber attack) through regular information and emergency drills. | new | | 90,00% | 2,5 | 2,9 | Free text Delphi | "The staff involved are regularly informed about the emergency concepts for cyber attacks, power failure, equipment failure (e.g. training, emergency drills)/ Proposal: "Possible scenarios such as equipment failure, power failure, ... are trained at regular intervals/prepared through emergency drills." |

**Appendix 7: Full list of patient safety indicators with Delphi ratings and source**

Article: Development and content validity of the Patient Safety in Radiation Oncology questionnaire (PaSaRO): A multi-method study

Authors: Maximilian Grohmann, Eva Christalle, Felicitas Schwenzer, Maria Jäckel, Nina Michalowski, Isabelle Scholl, Andrea Baehr

| Category | Sub-Category | Patient Safety Indicator (German) | Patient Safety Indicator | Change in wording after round 1 | Acceptance round 1 | Acceptance round 2 | Implementation feasibility rating (mean) | Ease of data collection rating (mean) | Reference | Excerpt |
|---|---|---|---|---|---|---|---|---|---|---|
| 3. quality and risk management | Failure of therapy-critical processes | Es gibt schriftlich dokumentierte Notfallkonzepte für den Ausfall von therapiekritischen Prozessen. | There are written emergency concepts documented for the failure of therapy-critical processes. | no | 92,30% | 92,30% | 2,9 | 3,2 | Bogusz-Czerniewicz et al., 2012, Rep. Pract. Oncol. Radiother. J. Gt. Cancer Cent. Poznan Pol. Soc. Radiat. Oncol. 17: 190–199, doi:10.1016/j.rpor.2012.05.001 | The institution has implemented and documented an emergency procedures in case of a defect or faulty operation of therapeutic apparatus/measuring equipment. |
| 3. quality and risk management | Standard operating procedures (SOPs) | Alle therapierelevanten Vorgänge sind schriftlich in Standardverfahren (SOPs) geregelt. | All therapy-relevant processes are regulated in written standard operating procedures (SOPs). | no | 93,40% | 93,40% | 3,1 | 3,4 | British Institute of Radiology et al., 2008, Towards Safer Radiotherapy | […] all routine procedures should be carried out in accordance with documented and approved management protocols and all non-routine work that may affect treatment outcome is to be approved through a system of written 'concessions'. |
| 3. quality and risk management | Standard operating procedures (SOPs) | Alle therapierelevanten Vorgänge werden gemäß den Standardverfahren (SOPs) durchgeführt. | All therapy-relevant procedures are carried out in accordance with the standard operating procedures (SOPs). | no | 93,40% | 93,40% | 3 | 2,9 | British Institute of Radiology et al., 2008, Towards Safer Radiotherapy<br>Focus group | […] all routine procedures should be carried out in accordance with documented and approved management protocols and all non-routine work that may affect treatment outcome is to be approved through a system of written 'concessions'.<br>And the point that I think is particularly important for patient safety, or ensures it particularly well, is clearly defined procedures. And then, of course, compliance with these procedures. |
| 3. quality and risk management | Standard operating procedures (SOPs) | Standardverfahren (SOPs) für therapierelevante Vorgänge werden regelmäßig, mindestens aber alle 2 Jahre auf Aktualität überprüft. | Standard operating procedures (SOPs) for therapy-relevant procedures are reviewed regularly, but at least every 2 years, to ensure that they are up to date. | no | 91,70% | 91,70% | 3 | 3,4 | British Institute of Radiology et al., 2008, Towards Safer Radiotherapy<br>American Society for Radiation Oncology, 2019, Safety is no accident A framework for quality radiation oncology care<br>Papakostidi et al., 2014, J. BUON Off. J. Balk. Union Oncol. 19: 47–52. | All procedures should be documented and subject to review every two years or whenever there are significant changes.<br>Documentation of SOPs is also critical to train new staff. Both training and documentation should be updated often. In particular, it is often necessary to retrain staff after time away from a system, or to refresh current knowledge.<br>Quality in radiotherapy is a dynamic concept that needs to be measured and re-evaluated using scientific methods and feedback by the users. |
| 3. quality and risk management | Standard operating procedures (SOPs) | Die Benennung von Terminen, Aufgaben, Konturen, Plänen, Feldern, CT-/IGRT-Protokollen und Berichten ist standardisiert. | The naming of appointments, tasks, contours, plans, fields, CT/IGRT protocols and reports is standardized. | no | 94,30% | 94,30% | 3 | 3,3 | Ford et al., 2020, Med. Phys. 47: 236–272, doi:10.1002/mp.14030<br>Mayo et al., 2018, Int. J. Radiat. Oncol. 100: 1057–1066, doi:10.1016/j.ijrobp.2017.12.013<br>Focus group | Standardization is a common theme for automation, i.e. that standards need to be in place for automation to be possible, as illustrated by some of the checks in Table 1.A.ii (e.g. technique, regimen, contour density overrides, motion management, all of which require standardization).<br>Standardized nomenclatures add value to the radiation oncology by providing a basis for improved communication and the ability to develop automated solutions for data extraction and QA to improve clinical workflow, safety, and research.<br>And what I also see from an RTT perspective is that, for example, target volumes are labeled ZV1 or 2, which is also difficult if I don't know exactly what is behind this. |
| 3. quality and risk management | Standard operating procedures (SOPs) | Die Konstanzprüfungen der Bestrahlungsausstattung wird regelmäßig auf Übereinstimmung mit aktuellen Normen und Leitlinien überprüft (z.B. DIN-Normen). | The constancy tests of the irradiation equipment are regularly checked for compliance with current standards and guidelines (e.g. DIN standards). | no | 92,80% | 92,80% | 3,1 | 3,2 | Bogusz-Czerniewicz et al., 2012, Rep. Pract. Oncol. Radiother. J. Gt. Cancer Cent. Poznan Pol. Soc. Radiat. Oncol. 17: 190–199, doi:10.1016/j.rpor.2012.05.001 | The institution has introduced a list of tolerances admissible in technical inspection and periodical equipment control tests, as required by national regulations. |
| 3. quality and risk management | Standard operating procedures (SOPs) | Die Patient:innenakte wird so geführt, dass sie eine zuverlässige Weitergabe aller sicherheitsrelevanter Informationen zwischen den Mitarbeitenden sicherstellt. | The patient file is managed in such a way that it ensures the reliable transfer of all security-relevant information between employees. | no | 98,60% | 98,60% | 3 | 3,1 | Focus group<br>Focus group<br>Patient interview | I see a lack of or poor communication, i.e. the flow of information, and then the documentation of this information as really dangerous.<br>I also see the lack of communication as a pretty big danger, because it happens relatively often that subsequent changes are made, for example to prescriptions or target volumes or whatever, that are not passed on and that could then slip through if it is not properly documented.<br>[...] there's always a file that's kept about you, which is also kept up to date. You can also have a look at it, I had a look at it too. Everything seems to be well kept in the file. And I think it is passed on in the same way. Or I think it's also passed on again - even those who are next in line, who don't know you yet, know a lot about you. |

#### Appendix 7: Full list of patient safety indicators with Delphi ratings and source

Article: Development and content validity of the Patient Safety in Radiation Oncology questionnaire (PaSaRO): A multi-method study

Authors: Maximilian Grohmann, Eva Christalle, Felicitas Schwenzer, Maria Jäckel, Nina Michalowski, Isabelle Scholl, Andrea Baehr

| Category | Sub-Category | Patient Safety Indicator (German) | Patient Safety Indicator | Change in wording after round 1 | Acceptance round 1 | Acceptance round 2 | Implementation feasibility rating (mean) | Ease of data collection rating (mean) | Reference | Excerpt |
|---|---|---|---|---|---|---|---|---|---|---|
| 3. quality and risk management | Technology assessment and risk analyses for new processes | Risikoanalysen werden für alle Prozesse und Therapieformen durchgeführt. | Risk analyses are carried out for all processes and forms of therapy. | no | 88,20% | 91,70% | 2,4 | 2,9 | Malicki et al., 2018, Radiother. Oncol. J. Eur. Soc. Ther. Radiol. Oncol. 127: 164–170, doi:10.1016/j.radonc.2018.04.006 | Risk assessment should be performed regularly and may include any or all of the following: (a) equipment; (b) processes; (c) human and organizational factors; and (d) external factors |
| | | | | | | | | | Klüter et al., 2021, Phys. Imaging Radiat. Oncol. 17: 53–57, doi:10.1016/j.phro.2020.12.005 | In total, 89 risks were identified for the entire MR-guided online adaptive workflow. After mitigation, all risks could be minimized to an acceptable level. |
| | | | | | | | | | British Institute of Radiology et al., 2008, Towards Safer Radiotherapy | When new or changed treatment techniques or processes are to be introduced, a risk assessment should be undertaken and consideration given to additional verification procedures for the initial cohort of patients. |
| | | | | | | | | | American Society for Radiation Oncology, 2019, Safety is no accident A framework for quality radiation oncology care | Hazard analysis, the active evaluation of the potential for failures that will cause incorrect results or harm to the patient, may be performed for any new system, to help delineate issues which can benefit from QC, QA, training or other mitigation strategies |
| 3. quality and risk management | Technology assessment and risk analyses for new processes | Risikoanalysen werden regelmäßig auf Aktualität überprüft. | Risk analyses are regularly reviewed to ensure that they are up to date. | no | 90,50% | 90,50% | 2,7 | 3,1 | Malicki et al., 2018, Radiother. Oncol. J. Eur. Soc. Ther. Radiol. Oncol. 127: 164–170, doi:10.1016/j.radonc.2018.04.006 | Risk assessment should be performed regularly and may include any or all of the following: (a) equipment; (b) processes; (c) human and organizational factors; and (d) external factors |
| | | | | | | | | | British Institute of Radiology et al., 2008, Towards Safer Radiotherapy | When new or changed treatment techniques or processes are to be introduced, a risk assessment should be undertaken and consideration given to additional verification procedures for the initial cohort of patients. |
| | | | | | | | | | American Society for Radiation Oncology, 2019, Safety is no accident A framework for quality radiation oncology care | Hazard analysis, the active evaluation of the potential for failures that will cause incorrect results or harm to the patient, may be performed for any new system, to help delineate issues which can benefit from QC, QA, training or other mitigation strategies. |
| 3. quality and risk management | Technology assessment and risk analyses for new processes | Das Team, das Risikoanalysen durchführt, wird durch eine Person mit Erfahrung im Risikomanagement moderiert. | The team that carries out risk analyses is moderated by a person with experience in risk management. | yes | 91,00% | 96,60% | 2,5 | 3,1 | Ford et al., 2014, Med. Phys. 41:, doi:10.1118/1.4875687 | The crucial role of the facilitator has been noted and supported by surveys of FMEA participants. |
| 3. quality and risk management | Technology assessment and risk analyses for new processes | Bei der Etablierung neuer Image-guided-radiotherapy-Hardware (z.B. surface-guided-radiotherapy, SGRT) werden PTV-Margins und Lagerung reevaluiert. | When establishing new image-guided radiotherapy hardware (e.g. surface-guided radiotherapy, SGRT), PTV margins and positioning are re-evaluated. | no | 96,40% | 96,40% | 2,9 | 3 | Al-Hallaq et al., 2022, Med. Phys. 49: 82–112, doi:10.1002/mp.15532 | Analogously to PTV margins, selection of these parameters is at the discretion of individual clinics and it is recommended that they be continuously assessed and updated. |
| 4. patient-specific processes | Selection and administration of tumor-effective systemic therapies | Bei Patient:innen, die systemische intravenöse Tumortherapien erhalten, werden an jedem Tag der Systemtherapie Vitalzeichen, Gewicht, Nebenwirkungen, Schmerzen und Reaktionen auf vorige Systemtherapien erhoben. | For patients receiving systemic intravenous tumor therapies, vital signs, weight, side effects, pain and reactions to previous systemic therapies are recorded on each day of systemic therapy. | yes | 76,90% | 97,30% | 3 | 3,3 | Neuss et al., 2017, Oncol. Nurs. Forum 44: 31–43, doi:10.1188/17.ONF.31-43 | On each clinical encounter or day of treatment, staff performs and documents a patient assessment that includes at least the following eight elements, and takes appropriate action: Functional status and/or performance status./Vital signs./Weight is measured at least weekly when present in the health care setting./ [...] /Allergies and previous treatment-related reactions./ Treatment toxicities./Pain assessment. |
| | | | | | | | | | Focus group | And at the same time they were weighed, which I have already said a few times because I miss it. Vital signs were taken as normal. And that was recorded so that you had a beautifully kept record and not what we know here, patients who lose 5 kilos and whoops, nobody noticed. |
| 4. patient-specific processes | Selection and administration of tumor-effective systemic therapies | Formulare für die Bestellung systemischer Tumortherapien enthalten in standardisierter Weise alle relevanten Informationen zu Medikation und Komedikation. | Forms for ordering systemic tumor therapies contain all relevant information on medication and co-medication in a standardized manner. | no | 91,70% | 91,70% | 3,2 | 3,3 | Neuss et al., 2017, Oncol. Nurs. Forum 44: 31–43, doi:10.1188/17.ONF.31-43 | Chemotherapy orders include at least the following elements: [...] Regimen or protocol name and number. Cycle number and day, when applicable. /All medications [...] Date of administration and Route of administration/[...] Sequencing of drug administration, when applicable./Rate of drug administration, when applicable. |
| 4. patient-specific processes | Selection and administration of tumor-effective systemic therapies | Die Verordnung von systemischen Tumortherapien wird durch Apotheker:innen gegengeprüft. | The prescription of systemic tumor therapies is cross-checked by pharmacists. | no | 92,30% | 92,30% | 3,2 | 3,2 | Huertas-Fernández et al., 2017, Clin. Transl. Oncol. Off. Publ. Fed. Span. Oncol. Soc. Natl. Cancer Inst. Mex. 19: 1099–1106, doi:10.1007/s12094-017-1645-y | […] over 90% of errors were detected during pharmacist validation. |

#### Appendix 7: Full list of patient safety indicators with Delphi ratings and source

Article: Development and content validity of the Patient Safety in Radiation Oncology questionnaire (PaSaRO): A multi-method study

Authors: Maximilian Grohmann, Eva Christalle, Felicitas Schwenzer, Maria Jäckel, Nina Michalowski, Isabelle Scholl, Andrea Baehr

| Category | Sub-Category | Patient Safety Indicator (German) | Patient Safety Indicator | Change in wording after round 1 | Acceptance round 1 | Acceptance round 2 | Implementation feasibility rating (mean) | Ease of data collection rating (mean) | Reference | Excerpt |
|---|---|---|---|---|---|---|---|---|---|---|
| 4. patient-specific processes | Selection and administration of tumor-effective systemic therapies | Für die Verabreichung von systemischen Tumortherapien sind Standards zum Umgang mit Paravasaten definiert und entsprechende Antidote sind vorhanden. | Standards for handling extravasates are defined for the administration of systemic tumor therapies and corresponding antidotes are available. | no | 100% | 100% | 3,5 | 3,4 | Vera et al., 2019, Clin. Transl. Oncol. Off. Publ. Fed. Span. Oncol. Soc. Natl. Cancer Inst. Mex. 21: 467–478, doi:10.1007/s12094-018-1945-x | Extravasation management procedures are defined and antidotes with protocols for using them are available. |
| 4. patient-specific processes | Selection and administration of tumor-effective systemic therapies | Patient:innen mit oraler systemischer Tumortherapie erhalten eine auf einer Checkliste basierende Schulung und einen Medikationsplan für die Einnahme. | Patients with oral systemic tumor therapy receive training based on a checklist and a medication plan for their intake. | no | 85,70% | 100% | 3,1 | 3,2 | Weingart et al., 2011, J. Oncol. Pract. 7: 2–6, doi:10.1200/JOP.2010.000064 | Create checklists to guide and remind clinicians about key elements required for patient education |
| | | | | | | | | | Weingart et al., 2011, J. Oncol. Pract. 7: 2–6, doi:10.1200/JOP.2010.000064 | Provide patients with dosing calendars similar to those provided in clinical trials |
| 4. patient-specific processes | Selection and administration of tumor-effective systemic therapies | Patient:innen mit oraler systemischer Tumortherapie erhalten in definierten Abständen ärztliche Visiten hinsichtlich Verträglichkeit und Therapieadhärenz zu ihrer Medikation. | Patients with oral systemic tumor therapy receive medical visits at defined intervals to check their tolerance and adherence to their medication. | no | 85,70% | 97,10% | 3,2 | 3,3 | Dürr et al., 2021, J. Clin. Oncol. Off. J. Am. Soc. Clin. Oncol. 39: 1983–1994, doi:10.1200/JCO.20.03088 | [The intervention group received] an additional, intensified clinical pharmacological/pharmaceutical care, which included medication management and structured patient counseling, over a period of 12 weeks [...] Antitumor drug-related problems were significantly lower in the intervention compared with the control group. (3.85 v 5.81 [mean], P < .001). |
| 4. patient-specific processes | Selection and administration of tumor-effective systemic therapies | Die Verordnung der Medikation von stationären Patient:innen wird durch Apotheker:innen überprüft. | The prescription of medication for inpatients is checked by pharmacists. | no | 81,80% | 96,70% | 2,6 | 3,3 | Eishy Oskuyi et al., 2021, Casp. J. Intern. Med. 12: 53–58, doi:10.22088/cjim.12.1.53 | The clinical pharmacologist identified 936 drug-related problems (55.98% of the prescriptions). |
| 4. patient-specific processes | Treatment of patients with pacemakers and defibrillators | Bestrahlungspläne für Patient:innen mit Herzschrittmachern und Defibrillatoren werden mit Energien unter 10 MV geplant. | Radiation plans for patients with pacemakers and defibrillators are planned with energies below 10 MV. | no | 81,80% | 93,80% | 3,7 | 3,6 | Zaremba et al., 2016, Eur. Eur. Pacing Arrhythm. Card. Electrophysiol. J. Work. Groups Card. Pacing Arrhythm. Card. Cell. Electrophysiol. Eur. Soc. Cardiol. 18: 479–491, doi:10.1093/europace/euv135 | The Heart Rhythm Society/American Society of Anesthesiologists Expert Consensus Statement mentions that usage of high-energy photon beams might lead to device malfunctions. Similarly, recent multidisciplinary Dutch guidelines warn of using .10 MV photons in PM/ICD patients due to high risk of device malfunctions. |
| 4. patient-specific processes | Treatment of patients with pacemakers and defibrillators | Die Einteilung von Patient:innen mit Herzschrittmachern und Defibrillatoren zu verschiedenen Risikoklassen geschieht in Zusammenarbeit mit einer kardiologischen Abteilung und entsprechend der Empfehlung von Fachgesellschaften. | Patients with pacemakers and defibrillators are assigned to different risk classes in collaboration with a cardiology department and in accordance with the recommendations of specialist associations. | yes | 91,70% | 98,10% | 2,9 | 3,2 | Zaremba et al., 2016, Eur. Eur. Pacing Arrhythm. Card. Electrophysiol. J. Work. Groups Card. Pacing Arrhythm. Card. Cell. Electrophysiol. Eur. Soc. Cardiol. 18: 479–491, doi:10.1093/europace/euv135 | [...] clinical consequences of device malfunction during RT by classifying the risk to the patient into low, medium, and high. |
| 4. patient-specific processes | Treatment of patients with pacemakers and defibrillators | Die Schockfunktion von implantierbaren Defibrillatoren (ICDs) wird während der Bestrahlung deaktiviert. | The shock function of implantable defibrillators (ICDs) is deactivated during irradiation. | no | 88,90% | 93,20% | 3,2 | 3,3 | Zaremba et al., 2016, Eur. Eur. Pacing Arrhythm. Card. Electrophysiol. J. Work. Groups Card. Pacing Arrhythm. Card. Cell. Electrophysiol. Eur. Soc. Cardiol. 18: 479–491, doi:10.1093/europace/euv135 | Inactivation of antitachycardia therapies before RT by either reprogramming or application of a magnet to ICDs is recommended in several publications. |
| | | | | | | | | | Dobson et al., 2018, Curr. Probl. Cancer 42: 443–448, doi:10.1016/j.currproblcancer.2018.06.015 | The Society of Radiographers advocates that all 87 ICDs are temporarily deactivated with a magnet during delivery of RT therapy. |
| | | | | | | | | | Dorenkamp et al., 2013, Strahlenther. Onkol. Organ Dtsch. Rontgengesellschaft Al 189: 5–17, doi:10.1007/s00066-012-0243-8 | Antitachycardic ICD therapies should be deactivated for the duration of the irradiation procedure. This can be achieved by prior reprogramming or by continuous magnetic application to the unit during the irradiation procedure (with appropriately programmed magnetic mode; [...] |
| 4. patient-specific processes | Radiation planning and plan evaluation | Ein:e unbeteiligte:r Medizinphysikexpert:in überprüft, ob der Bestrahlungsplan mit der Verordnung übereinstimmt und sinnvoll ist. | An uninvolved medical physics expert checks whether the radiation plan is in line with the prescription and makes sense. | yes | 100% | 96,20% | 3,1 | 3,3 | Wilkinson et al., 2013, Brachytherapy 12: 382–386, doi:10.1016/j.brachy.2013.03.002 | In our clinic, treatment plans are checked by a second person using a volume-based method. For single catheter applicators and "fixed" geometry applicators such as a tandemering combination, prescriptions often are to points (e.g., the gynecologic point A), and our method not only checks that the treatment time is within tolerance for the treatment volume but also that the volume is appropriate. |

#### Appendix 7: Full list of patient safety indicators with Delphi ratings and source

Article: Development and content validity of the Patient Safety in Radiation Oncology questionnaire (PaSaRO): A multi-method study

Authors: Maximilian Grohmann, Eva Christalle, Felicitas Schwenzer, Maria Jäckel, Nina Michalowski, Isabelle Scholl, Andrea Baehr

| Category | Sub-Category | Patient Safety Indicator (German) | Patient Safety Indicator | Change in wording after round 1 | Acceptance round 1 | Acceptance round 2 | Implementation feasibility rating (mean) | Ease of data collection rating (mean) | Reference | Excerpt |
|---|---|---|---|---|---|---|---|---|---|---|
| 4. patient-specific processes | Radiation planning and plan evaluation | Für die Planqualitätsüberprüfung werden festgelegte Checklisten verwendet. | Defined checklists are used for the plan quality review. | no | 90,00% | 96,20% | 3,2 | 3,4 | Nealon et al., 2022, J. Appl. Clin. Med. Phys. 23: 13694, doi:10.1002/acm2.13694 | […] the checklist significantly improved the rate of error detection from 3.4 ± 1.1to4.4± 0.74 errors per participant without and with the checklist. |
| 4. patient-specific processes | Radiation planning and plan evaluation | Bei der Überprüfung des Bestrahlungsplans werden DVH-Constraint-Checks dokumentiert. | DVH constraint checks are documented when the treatment plan is reviewed. | no | 90,90% | 90,90% | 3,3 | 3,5 | IMRT Documentation Working Group et al., 2009, Int. J. Radiat. Oncol. Biol. Phys. 74: 1311–1318, doi:10.1016/j.ijrobp.2009.04.037 | Treatment plan goals such as acceptable target dose uniformity variation and minimum dose-volume constraint, and dose-volume limits on OARs, should be provided. |
| 4. patient-specific processes | Radiation planning and plan evaluation | Die Organbewegung wird durch definierte Sicherheitssäume berücksichtigt (planning risk volumes). Die resultierenden Strukturen bekommen eigene Dose-Constraints. | The organ movement is taken into account by defined safety margins (planning risk volumes). The resulting structures are given their own dose constraints. | yes | 90,90% | 91,30% | 3 | 3,2 | Expert opinion | |
| 4. patient-specific processes | Radiation planning and plan evaluation | Die Dosisberechnungseinstellungen wie Berechnungs-Algorithmus und -Auflösung werden in festgelegten Szenarien, z.B. bei Stereotaxie, auf definierte Weise angepasst. | The dose calculation settings such as the calculation algorithm and resolution are adjusted in a defined manner in specified scenarios, e.g. for stereotaxy. | no | 83,30% | 100% | 3,2 | 3,2 | Kroon et al., 2013, Radiat. Oncol. 8:, doi:10.1186/1748-717X-8-149 | AXB is recommended instead of AAA for avoiding serious overestimation of the minimum target doses compared to the actual delivered dose. |
| 4. patient-specific processes | Radiation planning and plan evaluation | Die Bestrahlungsanweisung/Verordnung ist im Bestrahlungsplanungssystem digital hinterlegt und wird bei Erstellung des Planes automatisch übernommen (z.B. Fraktionierung und Dosis). | The irradiation instruction/prescription is stored digitally in the irradiation planning system and is automatically adopted when the plan is created (e.g. fractionation and dose). | yes | 100% | 100% | 3,2 | 3,5 | Siebert et al., 2022, Phys. Imaging Radiat. Oncol. 24: 53–58, doi:10.1016/j.phro.2022.09.006 | The process of reviewing treatment plans is a relevant topic to consider in risk analysis of the radiotherapy workflow. The review process could be improved by enhancements in the treatment planning systems, use of digital dose prescription, and treatment planning templates. |
| 4. patient-specific processes | Radiation planning and plan evaluation | Das Planungs-CT wird auf Artefakte überprüft und diese bei der Bestrahlungsplanung berücksichtigt (z.B. durch Dichteüberschreibung, Artefaktminderungs-Methoden oder Ausblockung). | The planning CT is checked for artifacts and these are taken into account during treatment planning (e.g. through density override, artifact reduction methods or blocking out). | no | 90,90% | 90,90% | 3,4 | 3,1 | Expert opinion | |
| 4. patient-specific processes | Radiation planning and plan evaluation | Es wird sichergestellt, dass das gewählte Planungs-CT zur Dosisberechnung geeignet ist (z.B. aktuell, ohne Kontrastmittel und mit passender Hounsfield-Kalibrierkurve). | It is ensured that the selected planning CT is suitable for dose calculation (e.g. current, without contrast agent and with a suitable Hounsfield calibration curve). | no | 100% | 100% | 3,4 | 3,3 | Expert opinion | |
| 4. patient-specific processes | Radiation planning and plan evaluation | Der Allgemeinzustand der Patient:innen, insbesondere die Fähigkeit, die Lagerung zu tolerieren, findet Eingang in die Wahl der Bestrahlungstechnik (z.B. wenige Stehfelder statt modulierte Technik). | The patient's general condition, in particular their ability to tolerate positioning, is taken into account when choosing the radiation technique (e.g. few standing fields instead of modulated technique). | no | 100% | 100% | 3 | 2,9 | Expert opinion | |
| 4. patient-specific processes | Contouring | Mögliche Verzerrungen im MRT-Bild (sogenannte MRT-Verzeichnungen) von Patient:innen und dem MRT-Gerät werden berücksichtigt (z.B. durch Abgleich mit Kontrastmittel-CT) und möglichst minimiert. | Possible distortions in the MRI image (so-called MRI distortions) of the patient and the MRI device are taken into account (e.g. by comparison with contrast agent CT) and minimized as far as possible. | yes | 100% | 97,90% | 2,6 | 2,5 | Reynaert, 2019, Cancer Radiother. J. Soc. Francaise Radiother. Oncol. 23: 753–760, doi:10.1016/j.canrad.2019.08.002 | MRI sequences used for radiotherapy should thus be optimized to minimize geometrical distortions. |

#### Appendix 7: Full list of patient safety indicators with Delphi ratings and source

Article: Development and content validity of the Patient Safety in Radiation Oncology questionnaire (PaSaRO): A multi-method study

Authors: Maximilian Grohmann, Eva Christalle, Felicitas Schwenzer, Maria Jäckel, Nina Michalowski, Isabelle Scholl, Andrea Baehr

| Category | Sub-Category | Patient Safety Indicator (German) | Patient Safety Indicator | Change in wording after round 1 | Acceptance round 1 | Acceptance round 2 | Implementation feasibility rating (mean) | Ease of data collection rating (mean) | Reference | Excerpt |
|---|---|---|---|---|---|---|---|---|---|---|
| 4. patient-specific processes | Contouring | Radiolog:innen und Chirurg:innen können bei Bedarf zur Konturierung dazugerufen werden. | Radiologists and surgeons can be called in for contouring if required. | no | 90,00% | 100% | 2,3 | 2,7 | Sahgal et al., 2020, Pract. Radiat. Oncol. 10: 243–254, doi:10.1016/j.prro.2019.11.002 | In an ideal setting, the neurosurgeon would be involved in determining target volume and normal tissues, in particular for benign indications, functional indications, and complex metastasis including post-operative radiosurgery. |
| | | | | | | | | | Dimigen et al., 2014, Clin. Oncol. R. Coll. Radiol. G. B. 26: 630–635, doi:10.1016/j.clon.2014.04.030 | For the quality assurance audit meetings, the radiologist's review of 99 patients' planning contours resulted in a significant change in management in 6% of cases. |
| | | | | | | | | | Rosenthal et al., 2006, Head Neck 28: 967–973, doi:10.1002/hed.20446 | We suggest that the accuracy of head and neck radiation oncology treatment plans might be increased by co-examination by another head and neck cancer specialist, typically a radiation oncologist or head and neck surgeon, to confirm RT target volumes. |
| 4. patient-specific processes | First radiation treatment and subsequent radiation treatments | Alle strahlentherapeutischen Behandlungen werden von mindestens zwei Mitarbeitenden (z.B. zwei MT-Rs) durchgeführt. | All radiotherapy treatments are carried out by at least two employees (e.g. two RTTs). | no | 100% | 100% | 3 | 3 | Sundararaman et al., 2014, Pract. Radiat. Oncol. 4: 181–188, doi:10.1016/j.prro.2013.09.003 | The written protocols, procedures, and forms we developed for each type of treatment require a dialog between 2 staff members to check and verify such things as the identity of the patient, the site and side to be treated, the original prescription and plan, the expectation for the day's treatment, the consistency of equipment settings with the treatment plan, and the completion of all necessary documentation, etc. |
| | | | | | | | | | Marks et al., 2013, Pract. Radiat. Oncol. 3: 149–156, doi:10.1016/j.prro.2012.11.010 | A radiation therapist is responsible for daily setup accuracy, and thus a second radiation therapist should ideally provide daily review; i.e., 1 therapist setting the patient up with a second verifying, or the 2 therapists working together, and checking each other. |
| | | | | | | | | | Marks et al., 2013, Pract. Radiat. Oncol. 3: 149–156, doi:10.1016/j.prro.2012.11.010 | Similar peer review should ideally be performed for other therapist activities such as review of daily pre-treatment setup images, or review of respiratory gating parameters. |
| 4. patient-specific processes | First radiation treatment and subsequent radiation treatments | Bei nicht-koplanaren Feldern und Off-Isocenter-Bestrahlungen wird die Positionierungsgenauigkeit des Bestrahlungssystems berücksichtigt (z.B. in Bezug auf die Größe von Sicherheitssäumen). | For non-coplanar fields and off-isocenter irradiations, the positioning accuracy of the irradiation system is taken into account (e.g. with regard to the size of safety margins). | no | 100% | 100% | 3 | 2,8 | Pudsey et al., 2022, J. Appl. Clin. Med. Phys. 23: 13665, doi:10.1002/acm2.13665 | Rotational errors significantly impacted PTV dose coverage,especially in the couch angle. |
| | | | | | | | | | Hartgerink et al., 2019, Acta Oncol. Stockh. Swed. 58: 1275–1282, doi:10.1080/0284186X.2019.1633016 | The patient can be accurately positioned at couch 0 degrees using CBCT online images. If a non-coplanar technique is applied, the LINAC should also have a means of verifying patient position at different couch angles. |
| 4. patient-specific processes | First radiation treatment and subsequent radiation treatments | Während der gesamten Bestrahlungssitzung findet eine kontinuierliche Positionsüberwachung mittels Surface-guided radiotherapy (SGRT) oder MRT statt. | Continuous position monitoring by means of surface-guided radiotherapy (SGRT) or MRI takes place during the entire radiotherapy session. | no | 90,90% | 90,90% | 2,5 | 3,1 | Al-Hallaq et al., 2021, Radiother. Oncol. J. Eur. Soc. Ther. Radiol. Oncol. 163: 229–236, doi:10.1016/j.radonc.2021.08.008 | A total of 849/9737 events occurred during the pre-treatment review/verification and treatment stages. Of these, 179 (21%) events were predicted to have been preventable with SGRT. The most common preventable events were wrong isocentre (43%) and incorrect accessories (34%). |
| | | | | | | | | | Jiang et al., 2008, Int. J. Radiat. Oncol. Biol. Phys. 71: 103–107, doi:10.1016/j.ijrobp.2007.07.2386 | A program of frequent imaging throughout treatment is important for monitoring of and, if necessary, correcting for interfractional variations. |
| 4. patient-specific processes | First radiation treatment and subsequent radiation treatments | Es existiert ein Standard zum regelmäßigen Abgleich von Planungs-CT und Bildgebung am Bestrahlungsgerät (optisch oder automatisiert). Es sind Grenzwerte für Abweichungen definiert und die Abläufe beim Überschreiten der Grenzwerte sind standardisiert. | There is a standard for regular comparison of planning CT and imaging on the irradiation device (optical or automated). Limit values for deviations are defined and the procedures for exceeding the limit values are standardized. | no | 100% | 100% | 3,2 | 3,1 | Schaly et al., 2021, J. Appl. Clin. Med. Phys. 22: 168–174, doi:10.1002/acm2.13342 | Despite the false negatives and false positives, analysis of the timing of alert triggers showed that the alert system could have resulted in seven fewer clinical misses. The alert system has the potential to be a valuable tool to complement human judgment and to provide a quality assurance safeguard to help improve the delivery of radiation treatment of head and neck cancer. |
| | | | | | | | | | Focus group | I would very much like our doctors to review all CTs at least once a week, and we do ConeBeam CTs on almost every patient, so that they review them at least once a week. [...] I would like to see that, I keep bringing it up because I know that there are very big differences in quality in my team [...]. |
| | | | | | | | | | Focus group | And if there is anything greater than 1.5-2, the doctors speak to the [...] RTTs of the week. What happened there? Why do you suddenly have a shift of 5 cm? Did something go wrong? And they also check the quality of the cone beam every day and whether the fusion is good [...]. |
| | | | | | | | | | Focus group | It should simply be written down in the system so that you can see that I make a comment and then somehow a comment comes back. Yes, it's always taken into account. The patient simply can't do any better. |
| 4. patient-specific processes | First radiation treatment and subsequent radiation treatments | Vor der Neueinstellung werden in einem Patient:innengespräch wesentliche Punkte der Therapie erneut besprochen. Dabei wird ggf. die adäquate Applikation parallel applizierter systemischer tumorwirksamer Therapien erfragt. | Prior to first radiation, key points of the therapy are discussed again in a patient consultation. If necessary, the adequate application of systemic tumor-effective therapies administered in parallel will be discussed. | new | | 92,00% | 2,7 | 2,9 | Free text Delphi | A patient consultation directly before the first radiation treatment helps to establish whether the patient has understood everything and whether, for example, systemic therapy etc. has been initiated. At the same time, it is also an opportunity to show and explain the treatment plan so that the patient understands what they are about to receive. |

**Appendix 7: Full list of patient safety indicators with Delphi ratings and source**

Article: Development and content validity of the Patient Safety in Radiation Oncology questionnaire (PaSaRO): A multi-method study

Authors: Maximilian Grohmann, Eva Christalle, Felicitas Schwenzer, Maria Jäckel, Nina Michalowski, Isabelle Scholl, Andrea Baehr

| Category | Sub-Category | Patient Safety Indicator (German) | Patient Safety Indicator | Change in wording after round 1 | Acceptance round 1 | Acceptance round 2 | Implementation feasibility rating (mean) | Ease of data collection rating (mean) | Reference | Excerpt |
|---|---|---|---|---|---|---|---|---|---|---|
| 4. patient-specific processes | Patient presentation, information, indication and prescription | Die psychosoziale Situation und die aktuelle psychische Belastung der Patient:innen wird in der Anamnese erfragt. | The patient's psychosocial situation and current psychological stress is asked about in the medical history. | no | 77,80% | 92,50% | 3 | 2,9 | Lennes et al., 2009, Clin. Lung Cancer 10: 341–346, doi:10.3816/CLC.2009.n.046 | Quality indicators: […] Patient reported provider attention to psychosocial stress factors and psychologic symptoms Patient reported provider attention to family members' psychosocial problems and problems related to living conditions. |
| 4. patient-specific processes | Patient presentation, information, indication and prescription | Vor Indikationsstellung zur Seedimplantation bei Prostatakarzinom werden die Kontraindikationen abgefragt: großer TURP Defekt (transurethrale Resektion der Prostata) und Ataxia teleangiektasia. Außerdem wird der IPSS (Internationaler Prostata-Symptom-Score) erhoben. | Before the indication for seed implantation in prostate cancer is established, the contraindications are queried: large TURP defect (transurethral resection of the prostate) and ataxia telangiectasia. The IPSS (International Prostate Symptom Score) is also recorded. | no | 100% | 100% | 3,5 | 3,3 | Davis et al., 2012, Brachytherapy 11: 6–19, doi:10.1016/j.brachy.2011.07.005 | Large TURP defects, which preclude seed placement and acceptable radiation dosimetry Ataxia telangiectasia |
| | | | | | | | | | Davis et al., 2012, Brachytherapy 11: 6–19, doi:10.1016/j.brachy.2011.07.005 | The recommended cutoff values for recent RTOG clinical trial eligibility range from 15 to 18. |
| 4. patient-specific processes | Patient presentation, information, indication and prescription | Vorbefunde und Krankengeschichte werden durch eine individuelle Anamnese im Patient:innen-Gespräch validiert. | Preliminary findings and medical history are validated through an individual anamnesis in the patient interview. | no | 100% | 100% | 3,2 | 2,9 | Patient interview | He asked me a lot of questions about the entire medical process. Sure, they have the file, but I think that's also super important to feel safe when the doctor or employee, whoever, asks questions about my illness and about me personally. That I have the feeling that they know exactly what has been done to me, what my case is. |
| | | | | | | | | | Patient interview | So he could have said, I've read everything about you, but it's different if he says, I've read everything about you or to ask, so now let's go through this together so that I'm fully informed, I'll ask you a few questions. |
| | | | | | | | | | Patient interview | Were there any other questions that you felt were particularly relevant to your safety? Participant: So [...] allergies and whether I take any medication or- And AH and what other tests I've had and things like that. Whether I had any illnesses and things like that. So, everything that's important, I think. |
| 4. patient-specific processes | Patient presentation, information, indication and prescription | Eventuelle Vorbestrahlungsunterlagen werden bei dosimetrisch relevanter Nähe zum neuen Zielgebiet eingelesen, registriert und bei der Planung berücksichtigt. | Previous irradiation plans are imported, registered, and considered during treatment planning if they are dosimetrically relevant to the new target volume. | yes | 100% | 100% | 2,9 | 3,2 | Gopan et al., 2016, Med. Phys. 43: 5181, doi:10.1118/1.4961010 | Of the 356 applicable events from the institutional database, 180/356 (51%) were detected or could have been detected by the pretreatment physics plan review. |
| 4. patient-specific processes | Patient presentation, information, indication and prescription | Alle Fälle werden in multidisziplinären Tumor-Boards besprochen. | All cases are discussed in multidisciplinary tumor boards. | no | 87,50% | 98,00% | 3,1 | 3,4 | American Society for Radiation Oncology, 2019, Safety is no accident A framework for quality radiation oncology care | Modern oncology patient care often involves multiple modalities and can benefit from the review and discussion of experts in various oncology-related disciplines. [...] Therefore, regular presentation of cases at multidisciplinary physician conferences (tumor boards) is encouraged [...] |
| | | | | | | | | | Focus group | [...] there is now a certain obligation to discuss cases on an interdisciplinary basis, that attention is paid in the context of certifications to how the presentation rate is, how the tumor board decision adherence is. Yes, and I think it's absolutely important that one specialist discipline doesn't decide anything on its own. |
| 4. patient-specific processes | Patient presentation, information, indication and prescription | Bei Patient:innen mit starker Verschlechterung des Allgemeinzustandes unter Therapie werden mögliche Anpassungen des Konzeptes multiprofessionell in strukturierter Form besprochen. | For patients with a severe deterioration in their general condition during treatment, possible adjustments to the concept are discussed in a structured, multi-professional manner. | no | 87,50% | 100% | 2,9 | 3 | Focus group | I was actually just thinking about whether it would be possible to stop these prematury ends of treatments, so to speak, whether it would be possible to say, hey, we should somehow organize it again, [...], that we take it up once and say, guys, we need to talkabout whether we should continue like this. |
| | | | | | | | | | Focus group | [...] and we then pass this on to our doctor, then there is a high probability that it will actually be discontinued or at least paused for a while or whatever. But it has to go that way first […]. |
| | | | | | | | | | Focus group | So I can decide for myself which concept I use for a breast [radiation] or wherever. But when it comes to really crucial things, there is no such thing. Why is there no consensus? Well, I could, for example, we have early meetings, I could use them for such relevant things. |

**Appendix 7: Full list of patient safety indicators with Delphi ratings and source**
Article: Development and content validity of the Patient Safety in Radiation Oncology questionnaire (PaSaRO): A multi-method study
Authors: Maximilian Grohmann, Eva Christalle, Felicitas Schwenzer, Maria Jäckel, Nina Michalowski, Isabelle Scholl, Andrea Baehr

| Category | Sub-Category | Patient Safety Indicator (German) | Patient Safety Indicator | Change in wording after round 1 | Acceptance round 1 | Acceptance round 2 | Implementation feasibility rating (mean) | Ease of data collection rating (mean) | Reference | Excerpt |
|---|---|---|---|---|---|---|---|---|---|---|
| 4. patient-specific processes | Patient presentation, information, indication and prescription | Im Aufklärungsgespräch werden die Hintergrundinformationen und Anweisungen zum Erreichen einer sinnvollen Blasen- und Mastdarmfüllung so vermittelt, dass Patient:innen sie gut verstehen und erinnern können (z.B. schriftliche Zusammenfassung der wichtigsten Informationen und Wiederholung). | During the pretreatment consultation, the background information and instructions for achieving sensible bladder and rectal filling are conveyed in such a way that patients can understand and remember them well (e.g. written summary of the most important information and repetition). | no | 100% | 100% | 3,2 | 2,9 | Focus group | I could think of something about prostate cancer, with the filled bladder and empty rectum, for example. If this is not properly communicated to the patient in the informative discussion, it can of course happen that he is first pushed through the CT, which would be an unnecessary exposure to radiation, only to find out that we have to repeat it. |
| | | | | | | | | | Focus group | We give the patients an information brochure on the subject, which tells them what they should eat, what they shouldn't eat, how and when they should go to the toilet and drink half a liter beforehand. It works [...] okay. But not always. |
| | | | | | | | | | Focus group | Go to the toilet an hour beforehand so that your bladder is empty and then drink half a liter, up to three quarters of a liter. But it's important that you do this. And I also say in principle when I say goodbye, remember to go to the toilet an hour before and then drink half a liter. |
| 4. patient-specific processes | Patient presentation, information, indication and prescription | Im Aufklärungsgespräch werden Informationen, die für die Organisation des Therapieablaufes wichtig sind, so vermittelt, dass Patient:innen sie gut verstehen und erinnern können (z.B. schriftliche Zusammenfassung der wichtigsten Informationen und Wiederholung). | Information that is important for the organization of the treatment process is conveyed in the pretreatment consultation in such a way that patients can understand and remember it well (e.g. written summary of the most important information and repetition). | no | 100% | 100% | 3,1 | 2,9 | Patient interview | I would have liked it if the radiotherapy procedure had been explained to me during the first consultation. You come in, you get marks on your body, you lie down, you have to breathe in, you have to hold your breath. The whole procedure takes about 20 minutes. Then they are ready, can get dressed again and leave. And they do this at the same time every day. I would have liked that. |
| | | | | | | | | | Focus group | Patients sometimes arrive at CT very unprepared. Sometimes I don't really know whether they are actually poorly informed or whether the patients simply receive such a flood of information, even during the consultation, whether a lot of things simply leak out. |
| | | | | | | | | | Focus group | And maybe you have to make sure that you give very clear, precise information and requests or information about what you expect and how you expect it. I always have the feeling that somehow it can be made a bit more precise, [...] I think it really is a bit like that, that the patient also has difficulties weighing up what is actually important for me the next day when I go for a CT scan or something like that, when he has been given information for an hour. |
| | | | | | | | | | Focus group | […] that patients really do receive a flood of information. It's really overwhelming. [...] Which at least helps to some extent, but I can't promise that I've learned to simply repeat the important things for everyone. In other words, during the prostate information session in particular, the reference to the planning CT and the radiotherapy is repeated at least three to five times. |
| | | | | | | | | | Patient interview | And the second appointment, the more practical one, naturally gave me the feeling that when I go there next week, I'll know what to expect. |
| 4. patient-specific processes | Patient presentation, information, indication and prescription | Den Patient:innen wird empfohlen, eine Begleitperson zum Aufklärungsgespräch mitzubringen. | Patients are advised to bring an accompanying person to the consultation. | no | 87,50% | 90,40% | 3,2 | 3,1 | Patient interview | So when you're in a doctor's appointment like that, you can't remember everything. That's why we often go there in pairs, so that one person knows something that the other might not have heard. |
| | | | | | | | | | Patient interview | As I said, I'm generally not alone, because it's a lot of information and you're also very emotionally affected by a conversation like this. That's why it's very important to me to do it in pairs. |
| | | | | | | | | | Patient interview | So it was important that someone was there, my wife in this case, so that we could go through it again afterwards. And [...] how did she mean it and what else did she say and so on. I thought that was very important. |
| 4. patient-specific processes | Patient-specific quality assurance (QA) | Alle Bestrahlungspläne werden mittels einer Berechnungsmethode überprüft, die unabhängig vom genutzten Bestrahlungsplanungssystem ist. | All treatment plans are checked using a calculation method that is independent of the treatment planning system used. | no | 93,20% | 93,20% | 3,2 | 3,6 | Thomadsen et al., 2014, Pract. Radiat. Oncol. 4: 65–70, doi:10.1016/j.prro.2013.12.005 | Treatment plans and programs are checked through independent verification before treatment delivery. |
| | | | | | | | | | Thomadsen, 2019, Health Phys. 116: 189–204, doi:10.1097/HP.0000000000001005 | Verification that the treatment plan does not contain errors, usually through comparison with data from the histories of previous similar plans or redundant calculations. To truly be an independent, redundant check through the use of a second calculation, the input data to the second calculation must come from the original images and not from the output of the primary treatment plan. |
| | | | | | | | | | Gabriele et al., 2016, Crit. Rev. Oncol. Hematol. 108: 52–61, doi:10.1016/j.critrevonc.2016.10.013 | […] to prevent dosimetric errors before dose delivery by means of patient specific pre-treatment QA that allows the detection of possible mismatches between the dose delivered by treatment machine and the dose calculated by the treatment planning system. |
| | | | | | | | | | British Institute of Radiology et al., 2008, Towards Safer Radiotherapy | The checking of any procedure or calculation should usually be carried out following a different method from that originally used (see Box 5.1 and Section 5.7.1). Checking a result by a different method avoids the possibility of repeating the same mistake. |
| 4. patient-specific processes | Planning imaging and positioning | PET- und MRT- Untersuchungen zur Bestrahlungsplanung werden in Bestrahlungsposition aufgenommen. | PET and MRI examinations for radiation planning are performed in the radiation position. | no | 78,6o% | 94,20% | 2,2 | 3,2 | Reynaert, 2019, Cancer Radiother. J. Soc. Francaise Radiother. Oncol. 23: 753–760, doi:10.1016/j.canrad.2019.08.002 | In general, for PET or MRI, imaging should be performed in RT treatment position (flat table with immobilization tools). This allows a much more precise image registration. |

**Appendix 7: Full list of patient safety indicators with Delphi ratings and source**

Article: Development and content validity of the Patient Safety in Radiation Oncology questionnaire (PaSaRO): A multi-method study

Authors: Maximilian Grohmann, Eva Christalle, Felicitas Schwenzer, Maria Jäckel, Nina Michalowski, Isabelle Scholl, Andrea Baehr

| Category | Sub-Category | Patient Safety Indicator (German) | Patient Safety Indicator | Change in wording after round 1 | Acceptance round 1 | Acceptance round 2 | Implementation feasibility rating (mean) | Ease of data collection rating (mean) | Reference | Excerpt |
|---|---|---|---|---|---|---|---|---|---|---|
| 4. patient-specific processes | Planning imaging and positioning | Patient:innen mit linksseitigem Mammakarzinom werden, falls möglich, in Atemanhaltetechnik behandelt. | Patients with left-sided breast cancer are treated using the breath-hold technique if possible. | no | 92,90% | 92,90% | 3,3 | 3,6 | Best et al., 2017, Radiother. Oncol. J. Eur. Soc. Ther. Radiol. Oncol. 123: 288–293, doi:10.1016/j.radonc.2017.03.022 | Patients with left sided breast cancer have access to deep inspiration breath hold (DIBH) techniques or alternate methods of sparing. |
| 4. patient-specific processes | Planning imaging and positioning | Die Bilder des Planungs-CT werden hinsichtlich Auffälligkeiten (z.B. neu aufgetretene Raumforderungen) gescreent. | The images of the planning CT are screened for abnormalities (e.g. newly occurring masses). | no | 100% | 100% | 3,2 | 2,9 | Patient interview | I then wondered whether anyone had even looked at this CT scan, because even as a layman it was obvious to me when I saw this image that something was wrong. |
| 4. patient-specific processes | Planning imaging and positioning | Den Patient:innen wird das richtige Verhalten während der Bestrahlung (z.B. Umgang mit Atembefehlen) so erklärt, dass sie es gut umsetzen können. | The correct behavior during irradiation (e.g. handling breathing commands) is explained to patients in such a way that they can implement it well. | no | 93,30% | 93,30% | 3,2 | 2,8 | Patient interview | During my first radiotherapy, my very first radiotherapy, [...] there was a very nice team who explained to me exactly what was happening now, what I had to do with the breathing command, because I didn't know how, what I had to do now to successfully carry out this radiotherapy. |
| 4. patient-specific processes | Planning imaging and positioning | Für die Durchführung des Planungs-CT werden den MT-Rs ausreichend Informationen zur Lagerung, Behandlungsgebiet und Besonderheiten zur Verfügung gestellt (z.B. mittels standardisierter Anmeldung). | For the planning CT, the RTTs are provided with sufficient information on positioning, treatment area and special features (e.g. by means of standardized registration). | no | 100% | 100% | 3,3 | 3,3 | Focus group<br>Focus group | That the target volumes are perhaps defined a little more specifically. It is often the case that it is not said exactly where radiation should actually be applied. Then you're just told to go from there to there.<br>We introduced a digital form for our CT a year and a half ago. On the subject of the scanning area. We have certain standards that you can simply click on with a tick. [...] And since then, the only thing we've really rarely had is that it goes wrong. |
| 4. patient-specific processes | Planning imaging and positioning | Der Abstand zwischen aktuellstem MRT und erster Bestrahlung für die Stereotaxie von Hirnmetastasen beträgt maximal 7 Tage. | The interval between the most recent MRI and the first radiation treatment for stereotactic brain metastases is a maximum of 7 days. | new | | 96,00% | 2,3 | 3,3 | Free text Delphi | Appropriate interval between diagnostic imaging and planning CT (e.g. MRI for SRS max. 7 days before radiotherapy) |
| 4. patient-specific processes | Prophylaxis of adverse effects | Patient:innen mit Kinderwunsch und erwarteter Beeinträchtigung durch eine Radio(chemo)therapie wird eine Vorstellung zum Fertilitätserhalt/zur Kryokonservierung unter Berücksichtigung der onkologischen Erkrankungssituation ermöglicht | Patients who wish to have children and are expected to be affected by radio(chemo)therapy are offered a presentation on fertility preservation/cryopreservation, taking into account the oncological disease situation | yes | 90,00% | 92,30% | 2,8 | 3 | Zavras et al., 2016, Clin. Med. Insights Oncol. 10: 49–57, doi:10.4137/CMO.S32811<br>Zavras et al., 2016, Clin. Med. Insights Oncol. 10: 49–57, doi:10.4137/CMO.S32811<br>Rae et al., 2020, Value Health J. Int. Soc. Pharmacoeconomics Outcomes Res. 23: 74–88, doi:10.1016/j.jval.2019.08.004 | In a former study, it has been estimated that a total radiation exposure of 20 Gy fractionated over 6 weeks in younger women and children produces sterility with 95% confidence. Similarly, in a recent retrospective study including prepubertal and pubertal girls, all patients receiving >15 Gy radiotherapy to the ovaries developed ovarian failure.<br>The likelihood of infertility after radiation of the testes depends on the dose to the testes, shielding, and fractionation (single dose vs. multiple doses). Doses as small as 0.1 Gy can result in decreased sperm counts, and doses of 1.5–4 Gy can result in permanent sterility.<br>Proportion of AYA patients who had fertility preservation discussion before treatment. |
| 4. patient-specific processes | Prophylaxis of adverse effects | Patient:innen unter Bestrahlung der Mundhöhle werden vor Bestrahlung zahnärztlich vorgestellt und nicht erhaltungsfähige Zähne werden entfernt. | Patients undergoing irradiation of the oral cavity are presented to the dentist before irradiation and teeth that cannot be preserved are removed. | no | 83,30% | 96,00% | 3,1 | 3,3 | Goh et al., 2023, Br. Dent. J. 234: 800–804, doi:10.1038/s41415-023-5864-z<br>Patient interview<br>Deutsche Gesellschaft für Mund-, Kiefer- und Gesichtschirurgie et al., 2021, S3-Leitlinie Diagnostik und Therapie des Mundhöhlenkarzinoms | Unrestorable or periodontally hopeless teeth are extracted with minimal trauma before radiotherapy.<br>Yes, at the very beginning, when it started, together with the radiotherapy and immunotherapy, I had four teeth extracted on both sides, yes.<br>Quality objective: Dental examination as frequently as possible before the start of radio(chemo)therapy for oral cavity carcinoma |
| 4. patient-specific processes | Prophylaxis of adverse effects | Patient:innen unter Bestrahlung der Mundhöhle und des Rachens erhalten vor Therapiebeginn eine Anleitung zur systematischen Mundpflege. | Patients undergoing irradiation of the oral cavity and pharynx receive instructions on systematic oral care before starting therapy. | no | 83,30% | 94,00% | 3,2 | 3 | Patient interview<br>DGHO et al., 2024, Konsultationsfassung S3-Leitlinie Supportive Therapie bei onkologischen PatientInnen | He could hardly eat, swallow and so on. And she made detailed enquiries and gave us advice on what we should do to make things better. No, like sage tea. [...] And I thought it was very good that the doctor or the person sitting there mentioned it again and. No, and that, well, I thought that was fine.<br>Standardized oral care for the prophylaxis of oral mucositis should be provided in all age groups and for all types of treatment with a risk of oral mucositis. This consists of: 1. Care by the patient [...] 2. Preventive measures by the dentist 3. Fluoridation to protect the teeth 4. Close clinical monitoring and advice during ongoing therapy. |

**Appendix 7: Full list of patient safety indicators with Delphi ratings and source**
Article: Development and content validity of the Patient Safety in Radiation Oncology questionnaire (PaSaRO): A multi-method study
Authors: Maximilian Grohmann, Eva Christalle, Felicitas Schwenzer, Maria Jäckel, Nina Michalowski, Isabelle Scholl, Andrea Baehr

| Category | Sub-Category | Patient Safety Indicator (German) | Patient Safety Indicator | Change in wording after round 1 | Acceptance round 1 | Acceptance round 2 | Implementation feasibility rating (mean) | Ease of data collection rating (mean) | Reference | Excerpt |
|---|---|---|---|---|---|---|---|---|---|---|
| 4. patient-specific processes | Prophylaxis of adverse effects | Es existiert ein Standard zur prophylaktischen und therapeutischen Hautpflege während der Therapie. | There is a standard for prophylactic and therapeutic skin care during therapy. | no | 83,30% | 94,10% | 3,3 | 3,3 | Focus group | The head and neck patient, that's the patient clientele that you get when you can no longer do it on an outpatient basis. And then the skin is already broken, so to speak. And there is somehow the possibility of working prophylactically, good skin care. |
| | | | | | | | | | Focus group | But I think that the doctor often has the problem that each of us has different ideas or preferences. Or a clue or no clue. |
| | | | | | | | | | Patient interview | [...] what comes to my mind is the question of the extent to which I can care for the areas of skin that have been irradiated in between. In retrospect, I missed that. |
| 4. patient-specific processes | Prophylaxis of adverse effects | Patient:innen mit Opioid-Therapie erhalten eine protokoll-basierte Prophylaxe von medikamentös-bedingter Obstipation. | Patients on opioid therapy receive protocol-based prophylaxis for drug-induced constipation. | no | 83,30% | 94,40% | 3,2 | 3,2 | Andresen et al., 2022, Aktualisierte S2k-Leitlinie chronische Obstipation der Deutschen Gesellschaft für Gastroenterologie, Verdauungs- und Stoffwechselkrankheiten (DGVS) und der Deutschen Gesellschaft für Neurogastroenterologie & Motilität (DGNM) | Constipation prophylaxis should be given at the start of opioid therapy. |
| 4. patient-specific processes | Spatial and temporal organization of patient pathways | Die Dokumentation von Vorbefunden, Therapien, Verläufen und weiteren Dokumenten erfolgt in einer digitalen Akte, die von allen Berufsgruppen innerhalb einer Abteilung genutzt wird. | Preliminary findings, therapies, courses of treatment and other documents are documented in a digital file that is used by all professional groups within a department. | no | 90,90% | 90,90% | 2,8 | 3,3 | Focus group | So I really praise the digital files, I would perhaps actually take that into account for the patient somehow, that it goes in that direction. Because I believe that a patient benefits from being able to communicate with each other via such a digital file. |
| | | | | | | | | | Focus group | I also think it's really valuable when it's digital. If only because the information is there everywhere. Because you don't create redundant information and you never know which one is the current one. |
| | | | | | | | | | Focus group | Exactly, [...], it would be nice if the whole thing could be used so that it works independently of the person in question. |
| 4. patient-specific processes | Spatial and temporal organization of patient pathways | Für ärztliche Gespräche mit Patient:innen wird ausreichend Zeit eingeplant, sodass alle relevanten Informationen besprochen werden können. | Sufficient time is scheduled for medical consultations with patients so that all relevant information can be discussed. | no | 90,90% | 90,90% | 2,7 | 2,8 | Patient interview | [...] sometimes it was a bit too quick. Can you understand what I mean by that? [...] So it was a bit like, I'm going through the cubicles now and asking people, because we have to do rounds now. Are you okay, are you all right? |
| | | | | | | | | | Patient interview | The doctors I spoke to about the side effects were, as I said, all very empathetic, asked questions, they always had questions, can we do something else? That was also the amount of time they took, I thought that was appropriate, but yes |
| 4. patient-specific processes | Spatial and temporal organization of patient pathways | Bei Notfällen kann der Bestrahlungsplanungsprozess so gestrafft werden, dass die Bestrahlung innerhalb von 24 Stunden beginnt. | In emergency cases, the radiotherapy planning process can be streamlined so that radiotherapy begins within 24 hours. | no | 100% | 100% | 3 | 3,3 | Chowdhry et al., 2021, Tech. Innov. Patient Support Radiat. Oncol. 19: 37–40, doi:10.1016/j.tipsro.2021.07.003 | For urgent situations, we found that our center can accommodate treatment within 24 h. For routine plans using 3D conformal radiation, an approximately 1-week turnaround time is needed. For patients being treated with IMRT/VMAT an approximately 2-week turnaround time is needed. |
| 4. patient-specific processes | Spatial and temporal organization of patient pathways | Für alle Prozessschritte wurden angemessene und realistische Zeitaufwände definiert. | Appropriate and realistic time requirements were defined for all process steps. | no | 100% | 100% | 2,7 | 2,8 | American Society for Radiation Oncology, 2019, Safety is no accident A framework for quality radiation oncology care | To avoid safety problems or quality lapses caused by rushing to meet unrealistic scheduling expectations, each practice should determine the appropriate time allocated for each step in the process of care. |
| | | | | | | | | | Focus group | But I often find that the time between the consultation and the CT scan is sometimes a bit short. For example, in the case of radiotherapy to the oral cavity, the dental splint is somehow not yet ready. Or then it's decided again that another tooth needs to be extracted. Sometimes it's a bit [...] difficult that they somehow turn up at the CT scan unprepared. |
| 4. patient-specific processes | Spatial and temporal organization of patient pathways | Für die Terminierung der Neueinstellungen und weiteren Bestrahlungen existiert ein Standard zur Abschätzung der Zeitdauer anhand von Bestrahlungsfeldern, Komorbiditäten und Allgemeinzustand. | There is a standard for scheduling new treatments and further radiotherapy to estimate the duration based on radiation fields, comorbidities and general condition. | no | 88,90% | 94,40% | 2,9 | 3,1 | Focus group | It's the same, what time factor do you use for the patients? For example, I know that the previous employer allowed ten minutes per patient. Now it's 15 minutes. But we sometimes have two patients in those 15 minutes. And sometimes with several target volumes, which of course are added on top of that. Then he may be lying in bed or is an anxious patient […] |
| | | | | | | | | | Focus group | But then you would have to differentiate between all that. Is it a pedestrian, is he bedridden? |
| 4. patient-specific processes | Spatial and temporal organization of patient pathways | Für Patient:innen ohne ausreichende Deutschkenntnisse wird ein sicherer Informationsfluss mit Dolmetscher:innen oder übersetzenden Angehörigen gewährleistet. | For patients without sufficient knowledge of German, a secure flow of information is ensured with interpreters or translating relatives. | no | 100% | 100% | 2,3 | 2,7 | Focus group | What's also difficult now with the number of foreign patients who don't speak German is that there's often no interpreter present during our planning CT. And especially when you have prostate patients, to whom you might have to say something about the bladder and bowel or other patients and you can't talk to them, then you're also quite limited. |
| | | | | | | | | | Focus group | On the negative side, I have also seen a lack of language skills, because this can often lead to misunderstandings on the part of both patients and practitioners of all kinds, which in turn can lead to errors. |

#### Appendix 7: Full list of patient safety indicators with Delphi ratings and source

Article: Development and content validity of the Patient Safety in Radiation Oncology questionnaire (PaSaRO): A multi-method study

Authors: Maximilian Grohmann, Eva Christalle, Felicitas Schwenzer, Maria Jäckel, Nina Michalowski, Isabelle Scholl, Andrea Baehr

| Category | Sub-Category | Patient Safety Indicator (German) | Patient Safety Indicator | Change in wording after round 1 | Acceptance round 1 | Acceptance round 2 | Implementation feasibility rating (mean) | Ease of data collection rating (mean) | Reference | Excerpt |
|---|---|---|---|---|---|---|---|---|---|---|
| 4. patient-specific processes | Spatial and temporal organization of patient pathways | Es existiert ein standardisiertes Vorgehen, das die Vorbereitung der stationären Patient:innen vor der Bestrahlung sicherstellt (z.B. Medikation, Blasenfüllung) | There is a standardized procedure that ensures the preparation of inpatients before radiation (e.g. medication, bladder filling) | no | 100% | 100% | 2,9 | 3 | Focus group | And maybe empty urine bags again with the preparations [...] But it's also about quality, and that's not a derogatory term, but those who are trained in this area naturally have a completely different view of the patients. |
| 4. patient-specific processes | Interfaces between practitioners | Es existiert ein standardisiertes Vorgehen, um externe Befunde und Bilder in einer adäquaten Zeit zur Therapieentscheidung und -planung verfügbar zu machen. | There is a standardized procedure for making external findings and images available in a timely manner for treatment decisions and planning. | no | 100% | 100% | 2,7 | 2,9 | Focus group | For me, this is a basic prerequisite for a good flow of information, namely digitalization. If this is lacking or incomplete, then we definitely have a loss of information and this is a problem between clinics in particular. If, for example, a patient brings in documents from outside, they end up in some file, the patient is then in another clinic, nobody has access to these documents [...]. |
| | | | | | | | | | Focus group | Well, that if it's external, that they really do contact us or the doctor and say, [...] that's why the chemo wasn't given today. [...]. It has to,[...] it has to come from the outpatient clinic or something, so that they can deal with us. And unfortunately that never happens. |
| | | | | | | | | | Focus group | [...] until I have the signed declaration, other institutes don't hand over any pictures, information or anything else that I might actually need urgently at that moment and until it's faxed like in the old days, because emails are forbidden or something else. So I think it's a huge disaster and, to be honest, it's also a source of error. |
| 4. patient-specific processes | Interfaces between practitioners | Es existiert ein zuverlässiger Standard zur Dokumentation einer durch eine:n Mitbehandelnde:n applizierten systemischen Therapie und Überwachung der Nebenwirkungen (z.B. Blutbildkontrolle). | There is a reliable standard for the documentation of systemic therapy applied by a co-treater and monitoring of side effects (e.g. blood count monitoring). | no | 100% | 100% | 2,8 | 3 | Neuss et al., 2017, Oncol. Nurs. Forum 44: 31–43, doi:10.1188/17.ONF.31-43 | The health care setting policy outlines the procedure to monitor an initial assessment of patients' adherence to chemotherapy that is administered outside of the health care setting. |
| | | | | | | | | | Focus group | So chemo is sometimes a problem where you only find out WHEN the CT is already running or the radiation is already going on. Damn, he should definitely have this one special treatment first or then he shouldn't have radiation. Then we have to clarify this again and we have already taken patients off the table. |
| | | | | | | | | | Focus group | You also need fixed times, for example, at which certain information must be available in the radiotherapy department or in the practice, which are checked regularly. For example, the weekly blood count check, or you agree that you always do it at one of the two locations |
| | | | | | | | | | Focus group | With the inpatient, it's really the case that when we call them from the ward, we ask what the chemo is like . |
| 4. patient-specific processes | Interfaces between practitioners | Bei Patient:innen mit vorgesehener sequentieller Chemotherapie/Erhaltungstherapie existiert ein standardisiertes Vorgehen, um deren Durchführung zu gewährleisten. | For patients with planned sequential chemotherapy/maintenance therapy, a standardized procedure exists to ensure its implementation. | no | 100% | 100% | 3,1 | 3 | DGP et al., 2024, S3-Leitlinie Prävention, Diagnostik, Therapie und Nachsorge des Lungenkarzinoms | Quality objective: Maintenance therapy with PD-L1 antibody durvalumab as frequently as possible after definitive radiochemotherapy without progression and PD-L1 expression ≥1% on tumor cells |
| 4. patient-specific processes | Interfaces between practitioners | Die Übergabe zwischen Schichten erfolgt nach einem Standard. | The transfer between shifts takes place according to a standard. | no | 88,90% | 98,00% | 3,2 | 2,9 | Hinkel, 2008, J. Natl. Compr. Cancer Netw. JNCCN 6: 528–35; quiz 534–535, doi:10.6004/jnccn.2008.0041 | Northwestern is now using the SBAR tool (situation background, assessment, and recommendation) to organize hand-offs of patients from one clinician or setting to another. |
| 4. patient-specific processes | Interfaces between practitioners | Die Übergabe von stationären Patient:innen zwischen Abteilungen erfolgt nach einem Standard. | The transfer of inpatients between departments takes place according to a standard. | no | 90,00% | 97,40% | 2,7 | 2,7 | Hinkel, 2008, J. Natl. Compr. Cancer Netw. JNCCN 6: 528–35; quiz 534–535, doi:10.6004/jnccn.2008.0041 | Northwestern is now using the SBAR tool (situation background, assessment, and recommendation) to organize hand-offs of patients from one clinician or setting to another. |
| 4. patient-specific processes | Interfaces between practitioners | Tagesaktuelle Ereignisse und mögliche Probleme werden täglich in strukturierter Form kommuniziert (z.B. in einer täglichen Besprechung). | Current events and potential problems are communicated daily in a structured form (e.g. in a daily meeting). | yes | 90,90% | 90,90% | 2,9 | 3,1 | American Society for Radiation Oncology, 2019, Safety is no accident A framework for quality radiation oncology care | Daily Morning Huddles Having all clinical staff meet daily to review the upcoming clinical activities can be a useful exercise to preempt potential problems and promote team work and safety culture. |
| | | | | | | | | | Focus group | In addition, it is reported when a patient has had diarrhea overnight, when someone has died, when someone has jumped out of the window. [...] In this respect, we really do have the whole team covered. It's important for the doctors, for the nurses it's rather annoying, as I said, but I think it's good that there's this meeting. |
| 4. patient-specific processes | Interfaces between practitioners | Für die gleichzeitige Nutzung mehrerer klinischer Informationssysteme existiert ein Standard für die sichere, kontinuierliche Informationsweitergabe (abteilungsintern und -extern) (z.B. automatische Übertragung von Abschlussberichten). | For the simultaneous use of several clinical information systems, a standard exists for the secure, continuous transfer of information (within and outside the department) (e.g. automatic transfer of final reports). | no | 90,90% | 90,90% | 2,4 | 2,7 | Focus group | So I say someone is isolated [for an infecitous disease] or something. For us, it's bright red marked. Then they say, why don't you have it? Yes, but we didn't look. Exactly. |
| | | | | | | | | | Focus group | So you might have to see if you can somehow link systems together. |
| | | | | | | | | | Focus group | And actually there should be some kind of networking. Because at the end of the day, it's still the same patient we're looking after. |

**Appendix 7: Full list of patient safety indicators with Delphi ratings and source**
Article: Development and content validity of the Patient Safety in Radiation Oncology questionnaire (PaSaRO): A multi-method study
Authors: Maximilian Grohmann, Eva Christalle, Felicitas Schwenzer, Maria Jäckel, Nina Michalowski, Isabelle Scholl, Andrea Baehr

| Category | Sub-Category | Patient Safety Indicator (German) | Patient Safety Indicator | Change in wording after round 1 | Acceptance round 1 | Acceptance round 2 | Implementation feasibility rating (mean) | Ease of data collection rating (mean) | Reference | Excerpt |
|---|---|---|---|---|---|---|---|---|---|---|
| 4. patient-specific processes | Symptom management | Es existiert ein Standard zur Bestimmung und Bewertung von Blutwerten bei ambulanten Patient:innen. | There is a standard for determining and evaluating blood values in outpatients. | no | 80,00% | 97,10% | 3 | 3,1 | Patient interview | But if it were the job of radiotherapy to at least say, please go to your GP every two weeks for a blood test as part of the radiotherapy. Then perhaps some things would have been detected beforehand. |
| 4. patient-specific processes | Symptom management | Es gibt klare Richtlinien zur Risikoeinschätzung und zum Management von Strahlentherapie-induzierter Übelkeit und Erbrechen (RINV). | There are clear guidelines for the risk assessment and management of radiotherapy-induced nausea and vomiting (RINV). | no | 77,80% | 97,10% | 3 | 3,1 | Dennis et al., 2012, Int. J. Radiat. Oncol. Biol. Phys. 84: 49–60, doi:10.1016/j.ijrobp.2012.02.031 | Risk estimates and management strategies for RINV varied, especially for low- and moderate-risk radiation therapy cases. |
| 4. patient-specific processes | Symptom management | Patient:innen werden im Laufe der Therapie mit einem implementierten Screening-Tool auf Mangelernährung gescreent und das Gewicht überwacht. | Patients are screened for malnutrition with an implemented screening tool during the course of therapy and their weight is monitored. | no | 81,80% | 91,90% | 3 | 3,2 | Patient interview | And then there was the fact that he told us he was going to lose weight but wasn't actually allowed to because otherwise the mask would no longer fit. And that's actually my biggest concern the whole time. |
| | | | | | | | | | Focus group | And at the same time they were weighed, which I have already said a few times because I miss it. Vital signs were taken as normal. And that was recorded so that you had a beautifully kept record and not what we know here, patients who lose 5 kilos and whoops, nobody noticed. |
| | | | | | | | | | Trujillo et al., 2021, J. Acad. Nutr. Diet. 121: 925–930, doi:10.1016/j.jand.2020.11.007 | The Academy of Nutrition and Dietetics supports the use of a single, validated tool, the MST, to screen adults for malnutrition. |
| | | | | | | | | | Kruizenga et al., 2005, Am. J. Clin. Nutr. 82: 1082–1089, doi:10.1093/ajcn/82.5.1082 | Early screening and treatment of malnourished patients reduced the length of hospital stay in malnourished patients with low handgrip strength (i.e., frail patients). |
| 4. patient-specific processes | Symptom management | Für die klinischen Visiten existiert ein Standard zur körperlichen Untersuchung, Erfassung von Nebenwirkungen, deren Dokumentation und Behandlung. | A standard for physical examinations, recording of side effects, their documentation and treatment exists for the clinical examinations. | no | 90,90% | 90,90% | 3,2 | 3,3 | Patient interview | So I was examined directly every time and both at this check-up appointment and at the appointment for reddening of the skin as well as at the follow-up appointment, someone always looked at it again. |
| | | | | | | | | | Patient interview | I had tension blisters from the radiation and that was also taken care of. I brought that up. And when they saw that, they kept saying, let me know or that's why you have these blisters and if anything happens, let me know. And then automatically an appointment was always made, so to speak, where everything was looked at again or a blood sample was taken. |
| 4. patient-specific processes | Symptom management | Für ambulante Patient:innen mit akuten Schmerzen ist die adäquate medikamentöse Analgesie in der strahlentherapeutischen Einrichtung unmittelbar möglich. | For outpatients with acute pain, adequate analgesia with medication can be administered immediately at the radiotherapy facility. | no | 81,80% | 94,60% | 2,7 | 3,1 | Focus group | It also takes time until the patient says she is in pain, then we know that [metamizol] is not enough for her. We need [piritramid] now, for example. Somehow she has to wait 20 minutes, 30 minutes [...] That's very unpleasant. |
| | | | | | | | | | Focus group | The medication pool can also be transferred from brachytherapy to teletherapy. So that you have your own small supply of medication. |
| 4. patient-specific processes | Symptom management | Für ambulante Patient:innen mit hoher Symtpomlast wird die Verordnung für eine Anbindung an einen spezialisierten ambulanten palliativ-Versorger (SAPV) ausgestellt und die Kontaktaufnahme koordiniert. | For outpatients with a high symptom burden, the prescription for a connection to a specialized outpatient palliative care provider (SAPV) is issued and contact is coordinated. | no | 90,00% | 92,10% | 2,9 | 3 | Patient interview | I had the palliative care team at my home for three months [...] I was feeling so bad, and that was good advice and they showed great support at home. |
| | | | | | | | | | Patient interview | But the palliative care team was very nice, it's great. |
| 4. patient-specific processes | Symptom management | Es existiert ein Standard zur regelmäßigen Fotodokumentation, z.B. von Tumoren und Hautreaktionen. | There is a standard for regular photo documentation, e.g. of tumors and skin reactions. | no | 75,00% | 93,20% | 3,2 | 3,3 | Focus group | So that would also be my wishful thinking, for example, to document the skin once a week. |
| | | | | | | | | | Focus group | […] because when the staff changes, you can see a lot more in the photo, how has it either worsened or changed for the better during aftercare. |
| | | | | | | | | | Focus group | I can really only recommend it. Which is the case with us now. We check anal rectum patients or vaginal tumors, for example. We really do a weekly skin check. If not twice, and actually look at how the skin is behaving right from the start. We document this with photos and so on. And if we notice that there are any abnormalities, a doctor is called in immediately. |
| 4. patient-specific processes | Symptom management | Patient:innen erhalten bei Bedarf in einem adäquaten Zeitfenster eine klinische Visite. | Patients receive a clinical visit within an appropriate time frame if required. | no | 100% | 100% | 3,2 | 3,2 | Patient interview | There was always someone there, someone was always there immediately, I didn't have to wait long or come back or anything, but was taken care of immediately. |
| | | | | | | | | | Patient interview | And if I mentioned something, I would speak to the doctor straight away, so to speak. I thought that was great too. |
| | | | | | | | | | Patient interview | Interviewer: In your opinion, what SHOULD the people treating you do to reliably recognize such side effects? Patient: The examinations were carried out. The eyes were examined again, etc. pp. So everything that can be done was actually done. |

#### Appendix 7: Full list of patient safety indicators with Delphi ratings and source

Article: Development and content validity of the Patient Safety in Radiation Oncology questionnaire (PaSaRO): A multi-method study

Authors: Maximilian Grohmann, Eva Christalle, Felicitas Schwenzer, Maria Jäckel, Nina Michalowski, Isabelle Scholl, Andrea Baehr

| Category | Sub-Category | Patient Safety Indicator (German) | Patient Safety Indicator | Change in wording after round 1 | Acceptance round 1 | Acceptance round 2 | Implementation feasibility rating (mean) | Ease of data collection rating (mean) | Reference | Excerpt |
|---|---|---|---|---|---|---|---|---|---|---|
| 4. patient-specific processes | Symptom management | Jede:r Patient:in mit maligner Erkrankung erhält mindestens einmal wöchentlich eine klinische Visite. | Every patient with malignant disease receives a clinical examination at least once a week. | yes | 75,00% | 94,10% | 3 | 3,3 | Gabriele et al., 2016, Crit. Rev. Oncol. Hematol. 108: 52–61, doi:10.1016/j.critrevonc.2016.10.013 | Numerator Number of clinical examinations carried out for each patient Denominator Total number of RT sessions Standard 1 clinical examination every 5 RT sessions (1 per week) = 100% |
| | | | | | | | | | Focus group | I also used to work in a surgical practice. I can recognize a lot of inflammation or something like that and I can feel it. If they say they have a hardening somewhere. But not everyone can do that today. We're not trained to do that and I wouldn't know how we could train them internally. So a different scheme would have to take effect somehow. |
| | | | | | | | | | Patient interview | So I remember at least every ten days or so, I think, or once a week, once a week, I think, I came into the cubicle where I was waiting to be irradiated, in any case a doctor came by. [...] He asked me how I was doing, how my skin looked. And so on. And I could, for example, if I somehow had the feeling, oh, it's getting red or something, I could always show that. |
| | | | | | | | | | Patient interview | Perhaps I would have liked you to take a look at it in between. So because I can't judge whether everything was good here. I always said it was all good. Because I didn't see anything bad. It was more the case that I was asked if I had a problem, if something seemed strange, if there was anything I should look at. But I usually said, no, everything's fine and then he or she moved on. |
| 4. patient-specific processes | Symptom management | Für die Behandlung aktuer Symptome (z.B. Übelkeit, Exsikkose) stehen Medikamente unmittelbar in der strahlentherapeutischen Einrichtung zur Verfügung. | For the treatment of current symptoms (e.g. nausea, exsiccosis), medication is available directly in the radiotherapy facility. | no | 80,00% | 94,40% | 3 | 3,2 | Focus group | The medication pool can also be transferred from brachytherapy to teletherapy. So that you have your own small supply of medication. |
| | | | | | | | | | Focus group | We have a doctor's room where all the medication is kept. On the ward, I don't know, I don't know anything about that. That's why I've been holding back. But there's a big cupboard downstairs by the equipment with all the medication. And then in the other doctors' rooms there are the most important medications. |
| | | | | | | | | | Focus group | I'm mainly at KH11, which is a private institution. And I just said I'd put a pool of medication on the device. |
| 4. patient-specific processes | Symptom management | Während der Therapie wird systematisch der Schmerzgrad erhoben und Schmerzen leitliniengerecht therapiert. | During therapy, the degree of pain is systematically assessed and pain is treated in accordance with the guidelines. | no | 83,30% | 94,40% | 3 | 3,1 | Vichare et al., 2013, Pract. Radiat. Oncol. 3: 37–43, doi:10.1016/j.prro.2012.09.005 | Documentation that the patient was screened for pain using a quantitative scale |
| | | | | | | | | | Albert et al., 2013, Int. J. Radiat. Oncol. Biol. Phys. 85: 904–911, doi:10.1016/j.ijrobp.2012.08.038 | [...] pain intensity quantified (on-treatment) //[...] plan of care for pain (on-treatment) |
| | | | | | | | | | Heneka et al., 2016, Palliat. Med. 30: 520–532, doi:10.1177/0269216315615002 | […] several common deviations from opioid prescribing guidelines were identified. These included no ‘as needed’ (PRN) analgesia ordered for patients with regular opioid orders (17%–29% of patients), no pre-emptive prescribing of anti-emetics and/or laxatives to treat opioid side effects (15%–24% of patients) and incorrect opioid dosing intervals (11%–81% of patients). |

**Appendix 7: Full list of patient safety indicators with Delphi ratings and source**
Article: Development and content validity of the Patient Safety in Radiation Oncology questionnaire (PaSaRO): A multi-method study
Authors: Maximilian Grohmann, Eva Christalle, Felicitas Schwenzer, Maria Jäckel, Nina Michalowski, Isabelle Scholl, Andrea Baehr

| Category | Sub-Category | Patient Safety Indicator (German) | Patient Safety Indicator | Change in wording after round 1 | Acceptance round 1 | Acceptance round 2 | Implementation feasibility rating (mean) | Ease of data collection rating (mean) | Reference | Excerpt |
|---|---|---|---|---|---|---|---|---|---|---|
| 4. patient-specific processes | Symptom management | Patient:innen werden im Zuge der Therapie auf psychische Belastung (Distress, Ängstlichkeit und/oder Depressivität) gescreent und über die Möglichkeit psychoonkologischer Unterstützung informiert. | Patients are screened for psychological stress (distress, anxiety and/or depression) in the course of treatment and informed about the possibility of psycho-oncological support. | no | 83,30% | 92,30% | 2,9 | 3,1 | Deutsche Gesellschaft für Mund-, Kiefer- und Gesichtschirurgie et al., 2021, S3-Leitlinie Diagnostik und Therapie des Mundhöhlenkarzinoms | Quality objective: Offer psychosocial care for oral cavity cancer as frequently as possible |
| | | | | | | | | | Rae et al., 2020, Value Health J. Int. Soc. Pharmacoeconomics Outcomes Res. 23: 74–88, doi:10.1016/j.jval.2019.08.004 | Proportion of programs that have psychology or psychiatry support available for AYA patients |
| | | | | | | | | | Rae et al., 2020, Value Health J. Int. Soc. Pharmacoeconomics Outcomes Res. 23: 74–88, doi:10.1016/j.jval.2019.08.004 | Proportion of AYA patients screened for distress |
| | | | | | | | | | Lennes et al., 2009, Clin. Lung Cancer 10: 341–346, doi:10.3816/CLC.2009.n.046 | Professional psychosocial care provided to patients who indicated that they needed it |
| | | | | | | | | | Holtzman et al., 2018, BMJ Open Qual. 7: 000034, doi:10.1136/bmjoq-2017-000034 | Of those screened, 21% had a positive test for depression, anxiety or both. This resulted in 5% of all patients or 26% of the screen-positive patients being referred to clinical psychology, a significant improvement compared with 0% before the intervention. |
| | | | | | | | | | Patient interview | Well, I did at the beginning, with this mask and all. But I regularly go to the oncopsychology department anyway, so I shared that with them. And I'm in good hands there with my whole illness. |
| | | | | | | | | | Patient interview | I can find out a lot of things myself on the Internet [...], but it wouldn't have been bad if you had been approached directly or told, please contact them, here is the address or here is another flyer. |
| | | | | | | | | | Patient interview | Well, I don't think they noticed because when I went into the treatment room, I pulled myself together. So I don't think they noticed at all. [...] Interviewer: And they didn't ask you, the practitioners, that you then somehow, yes, how your mental stress was? Patient: No. I knew exactly that I could get psychological help there too. |
| 4. patient-specific processes | Symptom management | Patient:innen haben Zugang zu psychoonkologischer Unterstützung. | Patients have access to psycho-oncological support. | no | 91,70% | 91,70% | 2,8 | 3,1 | Deutsche Gesellschaft für Mund-, Kiefer- und Gesichtschirurgie et al., 2021, S3-Leitlinie Diagnostik und Therapie des Mundhöhlenkarzinoms | Quality objective: Offer psychosocial care for oral cavity cancer as frequently as possible |
| | | | | | | | | | Rae et al., 2020, Value Health J. Int. Soc. Pharmacoeconomics Outcomes Res. 23: 74–88, doi:10.1016/j.jval.2019.08.004 | Proportion of programs that have psychology or psychiatry support available for AYA patients |
| | | | | | | | | | Lennes et al., 2009, Clin. Lung Cancer 10: 341–346, doi:10.3816/CLC.2009.n.046 | Professional psychosocial care provided to patients who indicated that they needed it |
| | | | | | | | | | Holtzman et al., 2018, BMJ Open Qual. 7: 000034, doi:10.1136/bmjoq-2017-000034 | Of those screened, 21% had a positive test for depression, anxiety or both. This resulted in 5% of all patients or 26% of the screen-positive patients being referred to clinical psychology, a significant improvement compared with 0% before the intervention. |
| | | | | | | | | | Patient interview | Well, I did at the beginning, with this mask and all. But I regularly go to the oncopsychology department anyway, so I shared that with them. And I'm in good hands there with my whole illness. |
| | | | | | | | | | Patient interview | Well, I don't think they noticed because when I went into the treatment room, I pulled myself together. So I don't think they noticed at all. [...] Interviewer: And they didn't ask you, the practitioners, that you then somehow, yes, how your mental stress was? Patient: No. I knew exactly that I could get psychological help there too. |

#### Appendix 7: Full list of patient safety indicators with Delphi ratings and source

Article: Development and content validity of the Patient Safety in Radiation Oncology questionnaire (PaSaRO): A multi-method study

Authors: Maximilian Grohmann, Eva Christalle, Felicitas Schwenzer, Maria Jäckel, Nina Michalowski, Isabelle Scholl, Andrea Baehr

| Category | Sub-Category | Patient Safety Indicator (German) | Patient Safety Indicator | Change in wording after round 1 | Acceptance round 1 | Acceptance round 2 | Implementation feasibility rating (mean) | Ease of data collection rating (mean) | Reference | Excerpt |
|---|---|---|---|---|---|---|---|---|---|---|
| 4. patient-specific processes | Symptom management | Patient:innen und deren Angehörige werden gezielt über den Umgang mit akut einsetzenden Symptomen oder Nebenwirkungen der Therapien inklusive Warnzeichen, die einen sofortigen Ärzt:inkontakt erfordern, unterrichtet. | Patients and their relatives are specifically instructed on how to deal with acute symptoms or side effects of therapies, including warning signs that require immediate contact with a doctor. | no | 90,90% | 90,90% | 3 | 2,8 | Greenall et al., 2015, J. Oncol. Pharm. Pract. Off. Publ. Int. Soc. Oncol. Pharm. Pract. 21: 26–35, doi:10.1177/1078155214556522 | The education of patients and family members or caregivers about potential side effects of medications and when to seek help is a vital component of ensuring safety, and one key element section of the self-assessment tool focused on patient education. |
| | | | | | | | | | Brahmer et al., 2018, J. Clin. Oncol. Off. J. Am. Soc. Clin. Oncol. 36: 1714–1768, doi:10.1200/JCO.2017.77.6385 | Patient and family caregivers should receive timely and up-to-date education about immunotherapies, their mechanism of action, and the clinical profile of possible irAEs prior to initiating therapy and throughout treatment and survivorship. |
| | | | | | | | | | Patient interview | There is also an emergency number for dermatology - I don't know about radiotherapy - but there is an emergency number for dermatology, which I also have. I've called it several times. It's really available - not 24/7, but during the times when you're present in the building, so I'm going to say from 8 am to 5 or 6 pm. |
| 4. patient-specific processes | Completion of therapy and follow-up care | Die Patient:innen erhalten zum Therapieabschluss in strukturierter Form Informationen zu weiteren Terminen, weiteren Behandlungen und Kontaktadressen. | At the end of therapy, patients receive structured information on further appointments, further treatments and contact addresses. | no | 91,70% | 91,70% | 3,4 | 3,2 | Patient interview | Exactly, that you just write it down. Your guideline, your tumor says every three months gyn for the injection and once a year this and that. So that you know, okay, I'll do all this and then it's the right therapy. |
| | | | | | | | | | Patient interview | But again, whether I should go for annual follow-up care or every two years? So maybe another one of those, what's going to happen in the long term and do I have to go to radiation aftercare for the rest of my life now, because I was irradiated once at the age of 30. Now that I'm thinking about it, I realize that I don't know exactly. |
| | | | | | | | | | Patient interview | You fall into a bit of a hole because you don't really know what to do next. [...] Simply the connection to how the therapy continues. Whether it's really enough if I go to my gynecologist every three months and also have an appointment for an MRI once a year. [...] |
| 4. patient-specific processes | Completion of therapy and follow-up care | Es wird sichergestellt, dass Patient:innen nach Therapieabschluss Folgetermine für Bildgebung und weitere Behandlungen durch Mitbehandelnde (z.B. niedergelassene Onkologie) terminieren und wahrnehmen. | It is ensured that patients schedule and attend follow-up appointments for imaging and further treatment by co-treaters (e.g. oncologists in private practice) after completion of therapy. | no | 92,30% | 92,30% | 2,6 | 2,6 | Patient interview | And it is SO difficult to get appointments, especially with specialists like mammography and radiology and things like that. In general, I don't know, many patients wouldn't even be accepted |
| | | | | | | | | | Patient interview | I'll be contacted again, similar to the appointment with you now. There's another appointment in July, which I got, but I'll be called again beforehand because it's a bit difficult to make appointments for such a long time. They wanted to contact me again to confirm it. |
| 4. patient-specific processes | Completion of therapy and follow-up care | Patient:innen nach thorakaler Bestrahlung, die eine Immuntherapie erhalten haben, werden in besonderem Maße auf die Gefahr einer Pneumonitis hingewiesen und nachgesorgt. | Patients after thoracic radiotherapy who have received immunotherapy are made particularly aware of the risk of pneumonitis and receive follow-up care. | no | 100% | 100% | 3,2 | 2,8 | Chen et al., 2021, Cancer Med. 10: 8518–8529, doi:10.1002/cam4.4363 | During follow-up, 47 patients (48.96%) developed symptomatic treatment-related pneumonitis (grade 2 n = 28, grade ≥3 n = 19) while six patients (6.25%) suffered from fatal toxicity. |
| 4. patient-specific processes | Completion of therapy and follow-up care | Hochgradige Nebenwirkungen während der Therapie werden systematisch erfasst. | Severe side effects during therapy are systematically recorded. | no | 92,30% | 92,30% | 3,2 | 3,1 | Deutsche Gesellschaft für Urologie et al., 2024, S3-Leitlinie Prostatakarzinom | Quality objective: CTCAE grade III or IV as rarely as possible after definitive radiotherapy |
| 4. patient-specific processes | Completion of therapy and follow-up care | Patient:innen erhalten vorsorglich bei Therapieabschluss Rezepte und Anweisungen zur Therapie häufiger subakut auftretender Nebenwirkungen. | As a precaution, patients receive prescriptions and instructions for the treatment of common subacute side effects at the end of therapy. | no | 92,30% | 92,30% | 3,1 | 3,1 | Patient interview | Interviewer: What support would you have liked, [...], what support would you have liked when dealing with side effects?<br>Patient: That they would have given us a prescription right away. For a burn ointment. I would have liked that. [...] that you get a doggy bag like that over the weekend. |
| | | | | | | | | | Patient interview | Well, that I should be careful, dizziness, severe headaches, vomiting, painkillers were given to me, which I didn't need. And if I experienced any severe pain or vomiting, I was told to go to the emergency room at KH1 immediately, but I didn't need any of that. |
| 4. patient-specific processes | Monitoring and adjustment of non-tumor-effective drug therapies | Vor der Applikation von knochenwirksamen Substanzen wird das Risiko von Kiefernekrosen eruiert. | The risk of jaw necrosis is determined before the application of bone-active substances. | no | 88,90% | 96,60% | 3,2 | 3,2 | Cardoso et al., 2023, Eur. J. Cancer Oxf. Engl. 1990 187: 105–113, doi:10.1016/j.ejca.2023.03.028 | The potential benefits must be counterweighed against associated risks (such as osteonecrosis of the jaw and atypical fractures). |
| 4. patient-specific processes | Monitoring and adjustment of non-tumor-effective drug therapies | Bei der Verordnung von Retard-Medikation wird das Vorhandensein eines künstlichen Darmausgangs und einer PEG Sonde erfragt und die Medikation ggf. angepasst. | When prescribing retard medication, the presence of an artificial bowel outlet and a PEG tube is queried and the medication is adjusted if necessary. | no | 87,50% | 96,40% | 3,2 | 3 | Hasait et al., 2015, Krankenhauspharmazie | [...] it can be postulated that no absorption problems should occur for 63% of the drugs investigated with a tmax between 0.5 and 2 hours. Alternatives [...] could be identified for 91% of all drugs investigated in order to achieve faster and thus improved absorption. |
